# A Systematic Review of Epidemiological Studies into Daylight-Saving Time & Health Identifying Beneficial & Adverse Effects

**DOI:** 10.1101/2025.03.17.25324086

**Authors:** Aiste Steponenaite, Jonas P. Wallraff, Ursula Wild, Lorna Brown, Ben Bullock, Gurprit S. Lall, Sally Ferguson, Russell G. Foster, Jennifer Walsh, Greg Murray, Thomas C. Erren, Philip Lewis

**Affiliations:** Medway School of Pharmacy, University of Kent and University of Greenwich, UK; Institute and Policlinic for Occupational Medicine, Environmental Medicine and Prevention Research, Faculty of Medicine and University Hospital of Cologne, University of Cologne, Germany; Centre for Mental Health, Swinburne University of Technology, Melbourne, Australia; Appleton Institute, Central Queensland University, Wayville, SA, Australia; Sir Jules Thorn Sleep and Circadian Neuroscience Institute (SCNi), Nuffield Department of Clinical Neurosciences, University of Oxford, Dorothy Crowfoot Hodgkin Building, South Parks Road, Oxford, OX1 3QU, UK.; Centre for Sleep Science, School of Human Sciences, The University of Western Australia, Crawley, Western Australia, Australia

**Keywords:** daylight saving, DST, time change, heart attack, myocardial infarction, mortality, mental health, psychiatry, suicide, traffic, accidents, circadian, sleep

## Abstract

**OBJECTIVE:** To systematically review and narratively synthesize the epidemiological evidence regarding health effects of Daylight-Saving Time (DST) practices.

**METHODS:** *Eligibility Criteria:* Peer-reviewed, primary research of humans published in English or German and that assessed either (i) health effects of transitions (we allowed up to one month as the index period), or (ii) health differences between living with DST vs Standard Time at a given time of year, or (iii) or health effects of living at the same latitude but different longitude within the same time zone were included. Eligible health effects were broadly categorised as all-cause mortality, accident (traffic/workplace/home/substance-related)-, cardiovascular-, gastrointestinal, immunologic-, psychiatric-, neurologic-, circadian and sleep-, and cognitive-related outcomes, as well as healthcare appointments/admissions. Adverse and beneficial effects were included.

*Information Sources:* PubMed, Web of Science Core Collection, Scopus, PsychINFO, and EconLit were systematically searched in March 2023 and then updated to June 2025.

*Synthesis Organisation:* Studies were organised by outcomes into the following categories: (1) cardiovascular, (2) psychiatric, (3) traffic accidents, (4) non-traffic accidents (e.g., workplace), (5) sleep and circadian, (6) cognitive, and (7) neurological and gastrointestinal and all-cause mortality, and narratively synthesised by at least two authors. Some studies were included under multiple categories. Synthesis conclusions were based on quality and quantity of studies. Next, a synthesis of the syntheses was written and confidence in validity of outcomes was illustrated with agreement from all authors. The authorship team included expertise in sleep and chronobiology with additional expertise in physiology, psychiatry, neuroscience, and epidemiology.

*Risk of Bias Assessment:* Critical appraisal was independently conducted by at least two authors using the Joanna Briggs Institute Critical Appraisal Checklist for Quasi-Experimental Studies, with studies categorized as low, medium, or high quality. Narrative syntheses of studies identified additional strengths and limitations.

**RESULTS:** *Included Results:* N=157 studies of diverse designs and varying quality from 36 different countries were identified. Exact participant numbers cannot be determined as many studies utilised nationwide databases/registries or included DST-associated assessments as either secondary objectives or as one part of a main objective (e.g., sleep assessment across a year). Nonetheless, the participant pool can be considered substantial.

*Synthesis:* DST-Onset transitions appear associated with increased acute myocardial infarction (17 studies, 6 high quality) and fatal traffic accidents (16 studies, 4 high quality) but decreased crimes involving physical harm (5 studies, 3 high quality). DST-Offset transitions appear associated with decreased all-cause mortality (3 studies, 1 high quality) and workplace accidents (8 studies, 2 high quality), but increased crimes involving physical harm. Living with DST (as opposed to Standard Time) is associated with decreased all-cause mortality and traffic accidents in summer, the latter only tested during initial months (2 high quality studies), and possibly sleep duration during winter (2 low quality studies). No studies identify clear effects on psychiatric outcomes (15 studies, 1 high quality). Apart from DST-Offset associations with increased sleep duration (22 studies, 2 high quality), limited studies prevent clear conclusions being drawn regarding sleep duration for other periods or regarding the timing of sleep onset and offset or regarding circadian rhythm disruptions (6 studies, 0 high quality).

**CONCLUSIONS:** What this review shows (including outcome-specific detrimental effects, null effects, and beneficial effects of transitions and of living with DST compared to Standard Time during summer months) is that the evidence does not support the messaging of transitions and DST during summer months being uniformly detrimental. The potential benefits of DST during summer on mortality and traffic accidents and of DST-Offset transitions on sleep, mortality, and work place accidents present an argument for maintaining transitions (from a health perspective) and for identifying mitigation strategies against adverse effects, at least for now, until further research suggests otherwise.

**REGISTRATION:** https://doi.org/10.17605/OSF.IO/R4W6M

## INTRODUCTION

Beginning in the 1840’s for railway timetables and prominent since the late 1800’s, Standard Time is used to facilitate coordination of human activities across regions and with respect to the sun. Standard Time can be defined as a local clock time of 12:00 coinciding with the sun reaching its zenith at a specific meridian line (a specific line of longitude) for a given time zone. However, time zone boundaries are set by political and geographical considerations. In many regions, there are also biannual time transitions that involve switching between daylight-saving time (DST) and Standard Time. The intention of DST during summer months is to increase available daylight in the evenings when (for many Western lifestyles) people are unconstrained by, for example, work or study commitments. This is possible because of seasonal differences in daylight duration in the mid-latitudes. Thus, DST also serves as a standardisation of clocks to coordinate activities across regions and with respect to the sun.

Transition from Standard Time to DST involves setting local or social clocks to a later time (typically by 1-hour).^1^ Transition back involves setting the clocks back. The terms “Spring Forward” to DST and “Fall Back” for returning to Standard Time are often used as a mnemonic to remember how to adjust clocks. As of 2025, transitioning to and from DST is practiced in 71 countries/autonomous territories, including in most European countries, Greenland, most states and provinces in the USA and Canada, several states in Mexico, Chile, Paraguay, several countries in the Caribbean, Egypt, a few Australian states, New Zealand, Israel, Palestine, and Lebanon. Morocco and Western Sahara set their clocks later for Ramadan. The transition dates (i.e., DST-Onset and DST-Offset) vary by location but are typically in spring and autumn, respectively.

Biannual time changes have become the subject of intense debate because of potential health and economic implications.^2–10^ The debated issues are, firstly, whether to keep or abolish DST-Onset and DST-Offset transitions. If transitions are abolished, the debate becomes whether to implement perennial DST or perennial Standard Time. DST and Standard Time have also been widely discussed in the field of sleep and chronobiology. Living or working against the circadian timing system (the internal body clocks that align physiology with daily cycles of light and dark) can be detrimental to health, as evidenced in studies of night and shiftworkers. ^11^ Changes in the social clock and subsequent timing of behaviour and timing of light exposures – as may occur with transitions – may affect circadian timing systems and subsequent physiology, performance, and health.^4^ ^12^

Many sleep and chronobiology academic societies have offered position statements in favour of abolishing DST and implementing perennial Standard Time (incl. American Academy of Sleep Medicine [AASM], Sleep Research Society, European Biological Rhythms Society, European Sleep Research Society, and Society for Research on Biological Rhythms). ^13–15^ These statements were a response to the USA’s Sunshine Protection Act to implement perennial DST and the European Union’s proposal to abolish transitions.^2^ ^3 16^ However, the evidence bases used to support the position statements or the political proposals are not comprehensive with regard to health. For example, one can consider the following statement by the AASM (Rishi et al. 2024, pg. 122): “The 1-hour time shift in the spring results in the loss of 1 hour of sleep opportunity, due to the presence of continuing social or occupational demands in early morning hours. This sleep loss accrues daily, resulting in ongoing sleep debt.”^14^ The study being referenced – nine healthy adults, aged 20-40 years, living in Finland, and tracked across one DST-Onset and one DST-Offset transition for one week – is not sufficient to support the statement.^17^ A comprehensive review of the epidemiological literature of effects on health of transitions and living with DST or Standard Time is warranted.

Whether and how several under-appreciated challenges to causal inference have been considered in studies is also important. Assessment of transition effects or effects of living with DST or Standard Time and application of chronobiological theory are further complicated in terms of defining exposures (Fig. 1), differences in individual responsiveness, and assessment of population level outcomes, and that the timing of sunrise and sunset is also significantly influenced by longitude within a time zone (the sun rises and sets at later clock times further west within a time zone) and latitude (the duration of time from sunrise to sunset is longer in summer and shorter in winter as latitude increases, thereby also affecting the timing of sun rise and sunset). At the exposure level, setting clocks forward or back can result in a shift of social activity timing and daylight exposures, the latter also depending on latitude and longitude. How long the potential effects of transitions might persist is an open question.^18^ Individuals can be expected to differ in their responses to transitions according to several factors; e.g., timing of more susceptible periods on the phase response curve to light,^19^ susceptibility to potential sleep disruptions (e.g., circadian linked, social responsibilities, or age),^20^ times of work and social commitments (or perhaps other behaviours such as timing of medication), susceptibility to other stress such as nocebo effects, and, of course and importantly, by health status. Population level health outcomes may also vary by the populations that are assessed (e.g., proportions of population at higher or lower risk of heart attack following a time transition). In the studies of DST effects on health, comparative risk analyses using years, times-of-year, and/or places wherein DST is not practiced would seem pertinent to account for residual confounding and chance, but such placebo tests appear to currently be the exception rather than the rule.^18^

**Fig 1.**
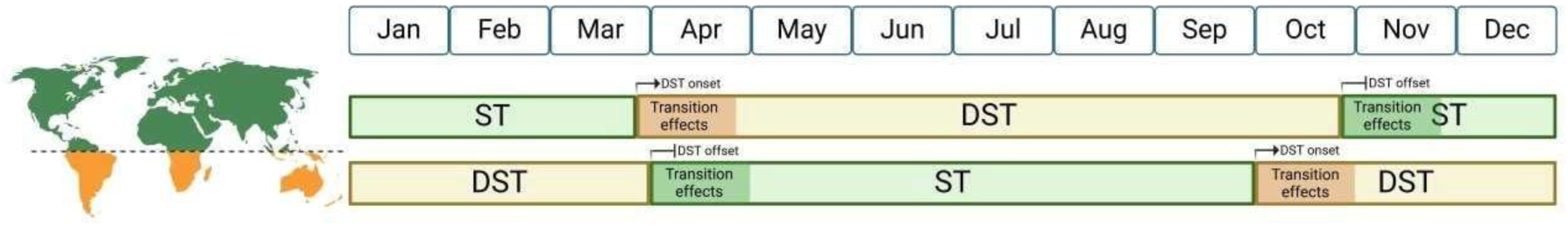
Schematic of potential exposures and time windows of interest. The DST period is marked by DST-Onset in spring and DST-Offset in autumn (approximate dates by hemisphere shown). While DST is not universally practiced, its implementation aims to extend evening daylight during summer. We ’allow’ index periods of up to one month post-transition to be considered, acknowledging that effects are harder to attribute further from the transition date. We also note that some studies include index and reference periods that both fall within one month post transitions. Additionally, studies comparing health outcomes during the DST period versus the Standard Time period (e.g., across summer months) are also relevant. Created in BioRender. Steponenaite, A. (2025) https://BioRender.com/x24e890

Overall, what is lacking in the current debate, and to inform policy, is a systematic review of the extensive epidemiological literature on DST and health. Our objective is to address this gap by providing a comprehensive synthesis of the available evidence.

## METHODS

### Study Design

This systematic literature review was conducted in accordance with the Preferred Reporting Items for Systematic Reviews and Meta-Analyses (PRISMA) guidelines^21^. The methods – including the rationale, search strategy, critical appraisal, and planned analysis – was detailed in a predefined protocol registered 22 June, 2023 on the Open Science Framework: https://doi.org/10.17605/OSF.IO/R4W6M.

### PICOS Framework

There were no population restrictions beyond “human”. Studies of effects of (i) transitions, (ii) living with DST vs Standard Time at a given time of year, or of (iii) living at different longitudes but similar latitude in the same time zone were considered relevant natural interventions. Effects of (ii) have also been inferred from (iii) in some instances. Comparators were based on time periods (e.g., pre-exposure vs post-exposure or corresponding periods with vs. without DST [i.e., index periods vs reference periods]) or location (e.g., population exposed vs. neighbouring population unexposed, or different longitudes but within the same time zone and latitude). We define transition index periods as up to 1-month post transition (i.e., shorter index periods are also included). Studies wherein reference periods include a pre-transition time period and post-transition period that falls within the first month post-transition are also accepted. Such studies attempt to account for potential seasonal trends. Relevant outcomes were broadly categorised as all-cause mortality, accident(traffic/workplace/home/substance-related)-, cardiovascular-, gastrointestinal, immunologic-, psychiatric-, neurologic-, circadian and sleep-, and cognitive-related outcomes, as well as healthcare appointments/admissions. Adverse and beneficial effects were included. Regarding circadian-related outcomes, changes or differences in circadian rhythm-related parameters were considered relevant. We also considered studies of cosinar activity parameters, social jet lag, and chronotype relevant. Measures of differences in incidence, prevalence, timing, risk, complications, deteriorations, and deaths for outcomes within the broader health outcome categories were considered relevant. There were no restrictions by study design.

### Eligibility Criteria

The eligibility criteria were defined in accordance with our PICOS framework and are presented in Table 1. In addition, only peer reviewed primary studies of humans with English and German as language restrictions were included. There were no restrictions by publication or study year. Studies that did not meet all of the eligibility criteria were excluded.

**Table 1:** Search strings & eligibility criteria

|  |  |
| --- | --- |
| **Medline**<br>**PsychINFO**<br>**EconLit** | All fields: "daylight savin*" OR "time change" OR "standard time" OR "wintertime" OR "summertime" OR "winter time" OR "summertime" OR "longitude" OR "time zone" OR "timezone" |
| **WOS**<br>**Scopus\*<sup>1</sup>** | All fields: "daylight savin*" OR "time change" OR "standard time" |
| **Eligibility Criteria** | 1) Peer reviewed primary studies of humans with English and German as language restrictions (no time restrictions)<br>2) Studies must consider either (i) the acute effects of transition in and/out of DST or (ii) DST vs standard time at a given time of year as exposure and control.<br>(i) *Acute setting*: Index periods as post-DST-related time change and reference periods as pre-DST-related time change (or also including periods of post-DST-related time change that occur after the index period) must be included in the studies of acute time change effects. The addition of a post-DST-related time change are sometimes included to determine acute effects different from seasonal trends. Time-series or regression discontinuity analyses will also be included.<br>(ii) *Non-acute setting*: These studies must involve an exposure vs control format as DST vs standard time at a given time of year such in adjacent regions that practice and do not practice DST, respectively. Alternatively, studies must involve regions at similar latitude and within the same time zone that differ in their sun time-official clock time relationship because of longitude.<br>3) Outcomes of interest are measures of incidence, prevalence, risk, complications, and/or deteriorations of accident-, cardiovascular-, metabolic-, gastrointestinal-, immunologic-, psychiatric-, neurologic-, circadian-, sleep-, and cognitive- related health outcomes, all-cause mortality and healthcare appointments/admissions. To add specificity on accidents, we intend any accidents related to traffic, workplace, home, substance-related, or other social setting. To add specificity to cognition, we intend any examination of conscious intellectual activity. To add specificity to healthcare appointments/admissions, we do not exclude any healthcare setting. To add specificity to cardiovascular-, metabolic-, immunologic-, psychiatric-, and sleep-related health outcomes, we do not require specification of ICD or DSM codes. |

<sup>1</sup>For the search with Scopus, additional filters included:
(a) Excluded subject areas: Engineering, Physics and Astronomy, Mathematics, Chemistry, Computer Science, Chemical Engineering
(b) Document type: Article
(c) Language: English and German

### Information Sources

We systematically searched for relevant literature across five major electronic databases: PubMed, Web of Science Core Collection, Scopus, PsychINFO, and EconLit. The initial search was conducted in March 2023, and updated to June 19, 2025. Further studies were identified through screening the reference lists of included studies and in feedback by peers.

### Search Strategy

The search strings (Table 1) were developed to capture a broad range of literature related to DST and its health impacts. Thus, search terms related to populations or specific outcomes were not used. Key terms across all databases included “daylight saving*”, “time change”, “standard time” and other pertinent terms for exposure. To optimize search results within specific databases, certain refinements were applied. For Web of Science and Scopus, the search strings were restricted to “daylight savin*”, “time change”, or “standard time”. Searches of Scopus were limited by excluding subject areas such as Engineering, Physics and Astronomy, Mathematics, Chemistry, Computer Science, and Chemical Engineering.

### Selection Process

All returned records were exported into the Endnote Reference software. Non-English or non-German studies were excluded according to the indicated Endnote language. If no language was indicated, the studies remained in screening. Duplicates were excluded using the Endnote de-duplication tool specified for digital object identifiers (DOIs). Duplicates without a DOI were identified and excluded manually. Remaining records were screened against the eligibility criteria presented in Table 1 using the Rayyan web tool and/or Endnote, initially by titles and abstracts and then by full texts. Screening steps were performed independently by at least two authors (AS, JPW, PL). Disagreements were resolved between the screening authors.

### Data Extraction, Critical Appraisal, and Syntheses

After the full text screening, AS and PL assigned the articles into one or more of the seven following categories: (1) cardiovascular, (2) psychiatric, (3) traffic accidents, (4) non-traffic accidents (e.g., workplace), (5) sleep and circadian, (6) cognitive, and (7) neurological and gastrointestinal and all-cause mortality. Two authors were assigned to extract data from-, critically appraise-, and write a narrative synthesis regarding the studies for each outcome category. The categories and responsible authors are presented in Table 2. Extracted data includes study identifiers, PICO information, study findings, and statistical methods. All study sizes (i.e., number of participants) and effect measures reported were considered relevant for synthesis.

**Table 2:**
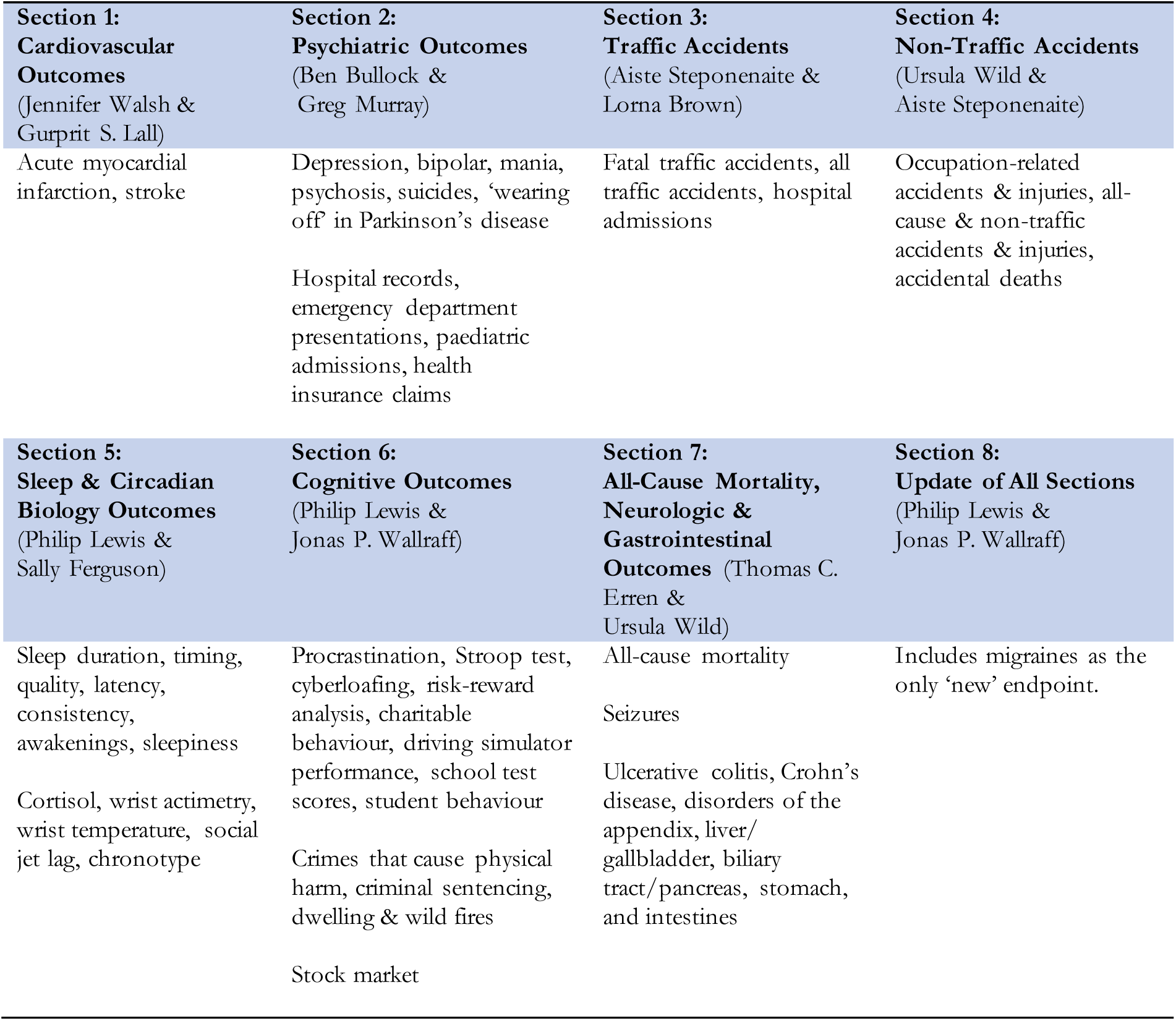
Broader Appendix Outcome Categories & Sections.

Given that most studies in this field are quasi-experimental in design, the Joanna Briggs Institute (JBI) Critical Appraisal Checklist for Quasi-Experimental Studies was used in this process.^22^ Based on the JBI checklist and independent appraisals, the quality of each study was categorized as low, medium, or high. Detailed rationales for these categorizations were provided, noting specific strengths and limitations of each study (e.g., see tables in the appendix for quality ratings and notes).

In-depth narrative syntheses for each category alongside tabulated overviews of extracted data and critical appraisals are presented in the Appendix. Descriptive, narrative syntheses were employed to present the findings due to the heterogeneity in study designs, populations, outcome measures, and statistical approaches among the included studies, which preclude formal meta-analyses. The detailed syntheses involved compiling and organizing reports of more specific outcomes within each category. Across studies, findings were summarized and consistency (or inconsistency) across studies was noted. The syntheses aimed to provide a comprehensive overview of the evidence, without including the individual study authors’ interpretations of their findings. A synthesis of the Appendix syntheses is presented in the Results section. The volume of studies and variety of outcomes necessitates such organisation. Furthermore, this approach is a clearer, more comprehensive, and systematic version of approaches used in the various opinion articles or position statements regarding this topic.

### Assessment of Confidence in the Validity of the Summarised Results

We addressed confidence in the validity of our summarised findings as follows: higher-quality rated studies were more heavily weighed, consistency in associations reported across higher-quality studies were more heavily weighted, and strengths of associations were also considered in narrative synthesis. While confidence was concluded from narrative syntheses, agreement from all authors concerning confidence was necessary (the authorship team included expertise in sleep and chronobiology with additional expertise in physiology, psychiatry, neuroscience, epidemiology, and occupational and environmental medicine).

## RESULTS

### Brief Overview

We identified 157 studies that adopted various designs (from registry analyses to small sample pre- and post-test repeated measure designs), investigated various populations (from everyone in registry catchment areas to adolescents in a school), and were conducted in various settings from schools to countries to continents (n=36 different countries). The flow of studies through the screening process is presented in Fig. 2. The use of robustness/falsification/placebo tests contributed to higher quality study ratings. Registry studies were also generally of higher quality given more comparable populations during index and reference periods of study.

**Fig 2.**
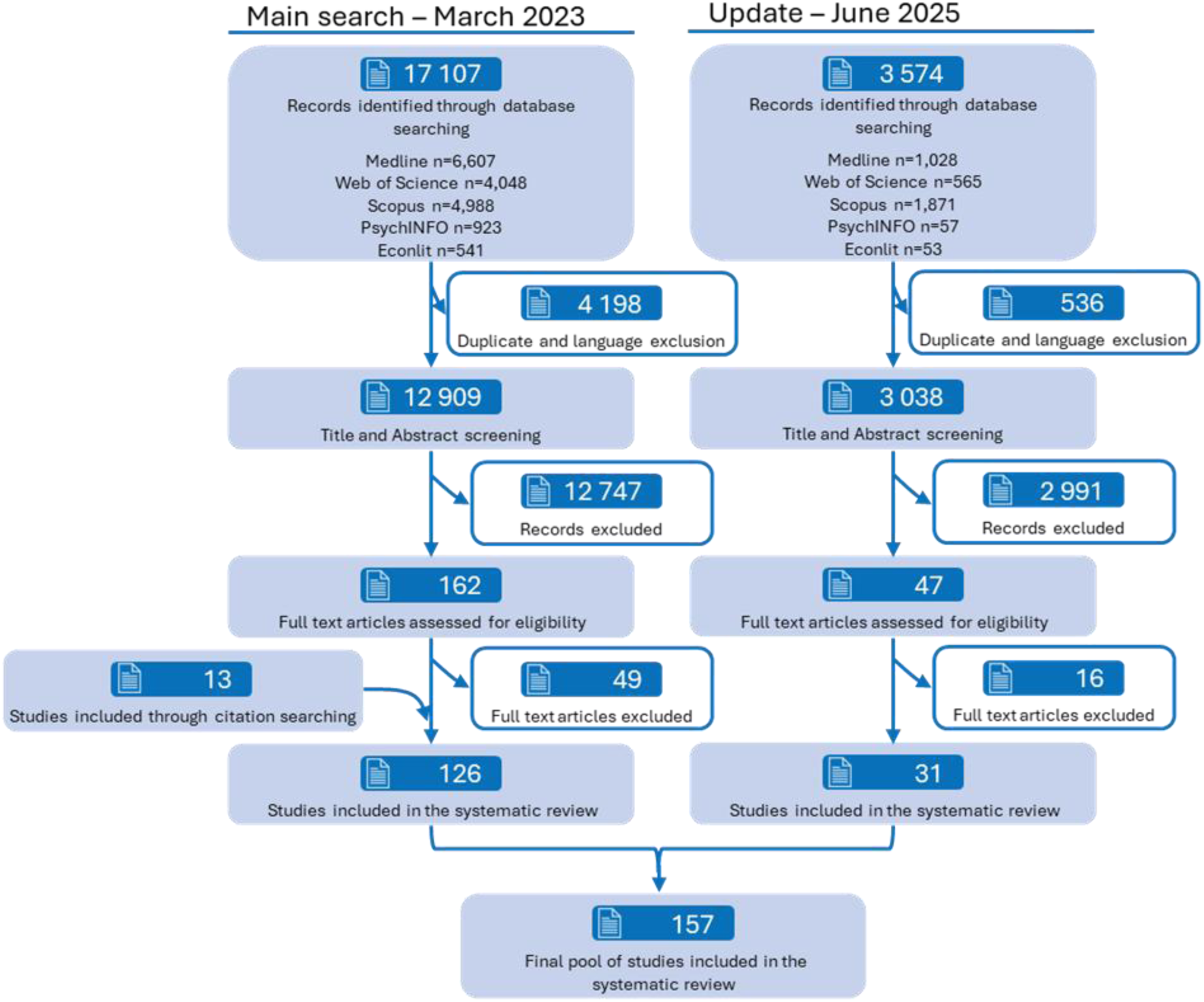
Study selection PRISMA flow chart.

An overview of identified outcomes within the seven broader categories is presented in Table 2. The categories were based on the identified outcomes and the volume of studies that would be assigned to the categories (as such, they differ slightly from the outcomes listed in our a-priori research question). An ‘Update’ category was added that includes all relevant studies identified up to June 2025.

Exact participant numbers cannot be determined as many studies utilised nationwide databases/registries or included DST-associated assessments as either secondary objectives or as one part of a main objective (e.g., sleep assessment across a year). Nonetheless, the participant pool can be considered substantial.

The vast majority of studies focus on transition effects. The vast majority of these define index periods between the day of transition to 2-weeks post transitions. Very few studies consider DST vs ST at a given time of year, which is not unexpected given the difficulty involved and rarity of opportunity.

Tabulated overviews of the extracted data and JBI checklist results for quality are also included in the Appendix. A synthesis of these syntheses is provided in the following paragraphs. The reader is referred to the sections of the Appendix (Table 2) for the syntheses of individual studies. It should be noted that not all studies assessing given outcomes agree – but the findings below are based on syntheses of all studies (including the volume and quality of studies reporting effects on outcomes and the effect sizes).

### Synthesis of Syntheses

#### DST-Onset and DST-Offset Transitions

The totality of the evidence from the identified studies suggests DST-Onset is associated with a short-term increased risk of cardiovascular events (especially myocardial infarction), and traffic accidents (especially fatal traffic accidents in the USA) – based on several high and medium quality studies with consistent findings. There may be individuals with particular susceptibility to cardiovascular events following DST-Onset.^23^ ^24^ One nationwide USA study reported no changes in all-cause mortality following DST-Onset.^25^ Changes in other endpoints following DST-Onset, including sleep duration and circadian biology-related outcomes (e.g., rhythm phase), are less clear with null or inconsistent non-null (i.e., different directions of change) associations reported in different studies of mostly medium-to-low quality. There are very few studies of circadian biology-related outcomes. If sleep duration is perturbed, it does not appear to be shorter than on many other days of the year (at the population level at least), although some individuals may be at higher risk of shorter sleep durations such as adolescents with early school start times.^26^ If circadian rhythms are perturbed, the phenomenon may be limited to more susceptible individuals as defined by, for instance, differences in typical health behaviours. ^27^ The timing of sleep onset or offset is likely to shift (albeit by much less than the 1 hour shift in clock time) for many people.^28^ ^29^ There are no apparent effects on psychiatric outcomes or work accidents following DST-Onset. A possible beneficial effect of DST-Onset is that crimes involving physical harm may decrease in the short term.^30^ ^31^ One medium quality study identified increased frequency of migraines among patients who suffer regularly after DST-Onset, but there was a two week delay before this effect manifested.^32^

The totality of the evidence from the identified studies suggests that DST-Offset may be beneficial in terms of increasing sleep duration^29^ ^33–36^ but detrimental in terms of increased incidence of crimes that cause physical harm.^30,37 38^ Workplace accidents may also be decreased following DST-Offset according to one high quality study.^39^ Migraine frequency may decrease after a two-week delay period according to one medium quality study.^32^ One high quality study reported decreased all-cause mortality following DST-Offset.^25^ Overall, the volume and quality of studies investigating DST-Offset effects are similar to DST-Onset, but no clear associations with endpoints other than those mentioned above are observed.

The lengths of investigated index and reference periods and the durations of transition-associated effects are different across studies and outcomes – no clear and conclusive findings can be described in this regard with the exception of increased risk of myocardial infarction in the first week following DST-Onset. No clear and conclusive findings can be determined with respect to geographical locations. Chronotype was investigated as a potential effect modifier in very few instances and no clear effects were observed.

#### Living with DST or Standard Time between Transition Dates

Far fewer studies assessed living with DST or Standard Time between transition dates compared to the effects of transitions. The limited quantity of evidence suggests that DST may be more beneficial than Standard Time across summer months as reflected in the mortality data in the high quality study by Cook (2022) in the USA and in the traffic accident data by Gillmore (2025) in Chile (at least in the initial months).^40^ ^41^ By contrast, DST across winter months may be detrimental based on studies of sleep, although these studies use less robust methods and are primarily based in one country (Russia).^42^ ^43^ DST may be preferable to Standard Time in the initial months of the time period wherein DST-Offset would occur in terms of decreased traffic accidents.^41^

### Reporting Bias

Given the overall volume (n=157) of studies and that many of these present null- or near-null effects, we conclude that publication bias is not strongly influencing our synthesis. There may be small-study-effects, but we give higher weight to studies of registries/databases that can also conduct placebo tests in their respective populations.

### Excluded Studies

Five studies that were excluded warrant brief mention as they might appear to meet the eligibility criteria at first glance. The study by Reis et al. (2023) compares Standard Time during winter months to DST during summer months, which is more akin to winter vs summer than to DST vs Standard Time.^44^ Similar to the study by Reis et al (2023), Zerbini et al. (2021) mention assessments under DST and under Standard Time, but during summer and winter months, respectively.^45^ This study and the related commentary articles are notable for highlighting difficulties when translating phase shifts in rhythms from local times to solar times and vice versa. ^45^ ^46^ The study by Giuntella et al. (2019) compares living in different time zones but close to the time zone border, which is not the same as living at a different longitude within the same time zone (and at the same latitude) because the neighbouring communities in the study by Giuntella et al. (2019) are working at different local times.^47^ Similary, Burns et al. (2025) assessed longitudinal differences in cognitive and academic performance parameters in the USA (not primary objective) but not within the same time zone.^48^ The study of sleep by Angelino et al. (2024) is included in the synthesis (rated low quality) but the authors also studied glucose control in type I diabetes patients without additional diabetes complications and with continuous glucose monitoring systems. The glucose time in range metric was 3-4% lower in the two weeks after transitions compared to the two weeks before transitions (which corresponds to a change from ∼9.5 days of time in range to ∼9 days of time in range). As the monitoring was continuous, this aspect of the study would receive a medium quality rating. Lastly, Dickinson et al. (2024) identified time-of-day (specifically, early morning) differences in GitHub activity in the two weeks post DST-Onset compared to pre-DST-Onset that might be taken to indicate decreased productivity, which might be considered a marker of decreased cognitive performance.^49^ The latter point is debatable because it would not be peak performance that is assessed. Of course, the observation may be due to the phase angle difference between performance phase and work time phase because work time will have shifted with DST-Onset.

### Certainty of Evidence

A tabulated summary of our synthesis of findings alongside our confidence in their validity is presented in Table 3.

**Table 3:** Summary Findings

| Outcome | Exposure | Effect | Confidence in validity* | Study Count & Quality* | References |
| --- | --- | --- | --- | --- | --- |
| **Transitions** |  |  |  |  |  |
| Myocardial infarction | DST-Onset:<br>DST-Offset: | ↑<br>↔ | High<br>Medium | 17 studies:<br>6 high & 10 medium quality | H: 23 68-72<br>M: 24 30 33 50 73-78<br>L: 79 |
| All-cause mortality | DST-Onset:<br>DST-Offset: | ↔<br>↓ | Medium<br>Medium | 3 studies:<br>1 high & 2 medium quality | H: 25<br>M: 80 81 |
| Fatal traffic accidents | DST-Onset:<br>DST-Offset: | ↑<br>↔ | Medium<br>Medium | 16 studies:<br>4 high & 10 medium quality | H: 41 65 67 82<br>M: 30 54-57 83-87<br>L: 88 89 |
| Crimes that cause physical harm | DST-Onset:<br>DST-Offset: | ↓<br>↑ | High<br>High | 5 studies:<br>3 high & 1 medium quality | H: 30 31 37<br>M: 38<br>L: 77 |
| Workplace accidents | DST-Onset:<br>DST-Offset: | ↔<br>↓ | Medium<br>Medium | 8 studies:<br>2 high & 4 medium quality | H: 39 90<br>M: 91-94<br>L: 95 96 |
| Psychiatric outcomes | DST-Onset:<br>DST-Offset: | ↔<br>↔ | Medium<br>Medium | 15 studies:<br>1 high & 8 medium quality | H: 97<br>M: 43 77 98-103<br>L: 24 104-108 |
| Circadian | DST-Onset:<br>DST-Offset: | ↔<br>↔ | Low<br>Low | 6 studies<br>1 medium quality | M: 28<br>L: 17 35 109-111 |
| Sleep Duration | DST-Onset:<br>DST-Offset: | ↔<br>↑ | Low<br>Low | 22 studies:<br>2 high & 2 medium quality | H: 26 29<br>M: 28 112<br>L: 33-36 93 104 109 113-123 |
| **Living with DST** |  |  |  |  |  |
| All-cause mortality | DST vs ST during summer | ↓ | Medium | 1 study:<br>1 high quality | H: 64 |
| Sleep Duration | Perennial DST vs biannual changes | ↓ | Low | 2 studies | L: 43 124 |
| Traffic Accidents | DST vs ST during late spring/early summer<br>DST vs ST during late autumn/early winter | ↓<br>↓ | Medium | 1 study:<br>1 high quality | H: 41 |

**Other Studies Synthesised but insufficient quantity, quality, consistency or too much heterogeneity between studies to provide confidence in validity:** <sup>27 32 63 125-175</sup> (stock market studies identified from citation searching are not included in this list – see Appendix Section 6)
↑, ↔, ↓ = increase, null/no clear effect, decrease
\*author confidence in true effect based on size of - and consistency in- effects observed, numbers of studies, and study quality;

## DISCUSSION

### Summary

In summary, this review identified n=157 studies concerning DST and health, ∼one in five were rated high quality, and quality varied across the outcome categories developed for synthesis. DST-Onset transitions appear (in the first week at least) to increase risk of acute myocardial infarction (RR ∼ 4% from a recent meta-analysis)^18^ but not all-cause mortality (both of these summary results include evidence from medium and high quality studies). DST in summer months between transitions may reduce all-cause mortality (RR ∼ -4.5%, based on one high quality study).^40^ DST-Offset transitions may reduce all-cause mortality (RR ∼ -2.5%, based on one high quality study).^25^ Following DST-Onset, there may also be a reduction in crimes that cause physical harm and a shift in the timing of sleep. The totality of the evidence suggests no or limited effects of DST-Onset on non-traffic accidents, psychiatric outcomes, cognitive measures, sleep parameters, or circadian rhythms, albeit studies on circadian rhythms are few. Following DST-Onset, increases in traffic accidents are observed in some places (in particular fatal accidents in the USA) but not everywhere. The volume and quality of studies from the USA suggest this finding is valid for the USA. DST-Offset transitions – apart from possible increases in crimes that cause physical harm – may be beneficial in terms of short-term increased sleep duration and decreased work place accidents in addition to decreased all-cause mortality.

### Mechanisms

In terms of mechanisms for the observed changes in cardiovascular outcomes, perturbation of sleep and circadian rhythms following transitions are often posited as causal. ^23^ ^50 51^ While the present review cannot exclude the possibility, the findings of no or small effects on sleep and circadian rhythms *per se* raise doubts about a simple causal relationship. Similarly, the evidence suggesting an absence of effect of DST-Onset on psychiatric or work accident outcomes is consistent with no or small effects on sleep and circadian rhythms given the known associations between them.^52^ ^53^ For acute myocardial infarction, individual baseline health could be important in terms of susceptibility to some brief and minor sleep loss. However, rather than an effect on sleep and circadian rhythms *per se*, the clock time change could result in changes in the timing of other exposures (relative to circadian rhythm phases) with health consequences, such as medication timing in susceptible individuals, but this is also speculative. Indeed, nocebo effects can also be offered as explanation, potentially fuelled by public statements about negative health effects caused by transitions and/or DST.^7^ Indeed, a combination of mechanisms seems likely to be involved. The null effects on all-cause mortality observed following DST-Onset in one high quality study (in the USA)^25^ may seem counterintuitive given the increased rates of acute myocardial infarction (as a high proportion of mortality can be attributed to myocardial infarction). However, while no significant difference for all-cause mortality was observed, the point estimate was compatible with increased mortality. Regarding the observed lower mortality during summer months of DST compared to Standard Time,^40^ mechanisms can only be speculated upon, such as individuals being more active and getting more sun exposure outside in the evening time with DST or reduced traffic fatalities.^41^ There is likely to be more than one factor driving differences in all-cause mortality. Reduced mortality following DST-Offset is compatible with reduced work accidents and increased sleep duration,^25^ ^29 30 39^ but any mechanistic attribution is speculative. The findings on traffic accidents and crimes that cause physical harm also warrant further mechanistic discussion. The short-term increase in traffic accidents following DST-Onset in some locations may be due to more difficult driving conditions at busier times of day (e.g., lower light levels during the morning rush with silver and white cars in particular being more difficult to see; more light in the evening increasing the number of people on the road). The role of time-of-day of increases in accidents reported by some studies implicates daylight availability for visibility at particular times as a candidate causal factor.^41^ ^54–57^ Not identified in our search (neither DST nor longitude mentioned in title or abstract), but relevant to discussion here, is the study by Gentry et al. (2022).^58^ The authors find increased traffic deaths in the USA in counties further west of time zone meridians but within the same time zone, including after stratifications by time zone and by area census designations of metropolitan or rural (albeit not for micropolitan). Further west means later sunrise and sunset times, which can be taken to suggest an effect of ambient light at specific times of day. The observed reduction in crimes that cause physical harm following DST-Onset may also be due to more visible light in the evenings and the converse following DST-Offset.^30^ ^37^ ‘Crimes that cause physical harm’ is used here as an umbrella term for robberies and assaults – such confrontations include knowledge of event timings. In contrast, a time range within which the event occurred may only be known for crimes that do not include confrontation, e.g., burglary. This is important for studies around a sharp timing cut off point such as DST-Onset or DST-Offset.

### Chronobiology and Sleep

Regarding chronobiology and sleep, adverse health consequences from small changes in the timing of sleep (i.e., onset and/or offset) following transitions cannot be ruled out. The count of studies on sleep is relatively high compared to other health outcomes, but they are often limited by low sample sizes or lack of repeated measures. Furthermore, changes that are observed in some studies are not necessarily different from what is observed on other days of the year. In addition, future studies would benefit from reporting and assessing data in terms of local clock time, sun time, and a standardised time wherein DST times are converted to their corresponding Standard Time times as this has been shown to aid interpretations.^45^ ^46^ Only a few studies consider circadian rhythms or chronotype. Furthermore, we were lenient in terms of what we included as markers of circadian rhythm; e.g., rhythmic parameters of 24-hour physical activity. Measures and comparisons of intra-individual rhythmic parameters of cortisol, melatonin, and core body temperature would be preferred. Regarding living with DST during summer months, it is worth noting that shifts in the timing of sleep appear to happen with progression into and out of summer months irrespective of transitions to and from DST;^2829^ thus, alignment of circadian rhythm phases and sun timing in summer months may not be significantly impacted by the presence or absence of DST.

### Policy and Positions

Regarding policy and position statements, the adverse effects observed following DST-Onset (most notably acute myocardial infarction) supports abolishing DST practice. Furthermore, the limited evidence on sleep, together with the chronobiological theory favouring Standard Time over DST during winter months (i.e., to facilitate earlier daylight exposures that prevent circadian rhythm phase delays)^4^, supports adopting perennial Standard Time if transitions were to be abolished.^59^ However, it is only after an appropriate balance of the adverse and beneficial effects that informed decisions can be made. What this review shows (including outcome-specific detrimental effects, null effects, and beneficial effects of transitions and of living with DST compared to Standard Time during summer months) is that the evidence does not support the messaging of transitions and DST during summer months being uniformly detrimental (Table 3). As a conceivable remedy to the differences in proposals and positions, and considering the different patterns in health outcomes, keeping DST and transitions that allow health benefits whilst developing and implementing strategies to mitigate negative patterns of health effects is worth consideration. Transition dates are known in advance and pre-emptive actions can be taken. To exemplify preventative candidate measures, myocardial infarction risk reduction in susceptible individuals with gradual phase shifting of behaviours and exposures (possibly including, for instance, medication timing, meals, and sleep) before transitions may be worthwhile exploring. Promoting such strategies may generally be worthwhile without the need for agreement over whether to maintain or abolish DST or even before drawing strong conclusions about the causal role of the meta-exposure of DST and transitions.^60^ Regarding traffic and crime, increasing a safety presence around transitions and at particular times of days (e.g., extra police presence, road safety officers, traffic speed cameras) could mitigate potential increased health risks.

### Who & Where

Our research question included health implications “for whom” and “where”. Some differences within and between studies are observed in this regard but within study findings are not conclusive and differences between studies cannot be concluded as attributable to differences in transition dates (e.g., different between the USA and the EU), latitudes, and longitudes. Although we cannot infer conclusions from the included studies, these factors warrant mentioning. Each of these factors can affect the clock timing of daylight availability. If transition dates were to be changed, changes in trade-offs between risks and benefits following transitions and living with DST or Standard Time are conceivable. Regarding latitude, transitions might be most beneficial in the mid-latitudes when differences in photoperiod across seasons are significant but not extreme and 1-hour changes in the clock timing of sunrise and sunset may have more impact on health. Closer to the equator and/or the poles, individual daylight exposures are unlikely to be different whether DST or Standard Time is in use. Regarding longitudes within a time zone that may affect morning light exposures, health inferences from the included studies are too limited. Cancer differences by longitude have been reported (increased in the west with later sunset timing), ^61^ which could be viewed as conflicting with reduced cancer-related deaths in older adults exposed to DST compared to Standard Time during summer months (with later sunset times). ^40^ However, the former could be driven by later sunrise and sunset times during winter months. It is important to note that findings from studies of differences by longitude in the same time zone should not be considered in debates of transitions, rather only in debates of perennial DST vs perennial Standard Time.^9^ Caution is also warranted when interpreting studies of longitude, as health impacts may not be monotonic, and social rhythms may differ across longitudes.^62^ Difficulties may also arise for countries that span a range of latitudes and/or longitudes within a time zone such as the USA, Australia, and Chile. From a health perspective, state differences may make most sense in the USA and Australia. Chile already considers regional differences. Chile is remarkable from the perspective of a very high population density around Santiago at ∼33.4 °S and much lower population densities into the Atacama Desert in the North (∼18-29°S) and the Patagonian fjords, temperate rainforests, and ice sheets in the South (∼45-55°S). In some regions, it may be easier to reschedule social behaviours and social commitments rather than shifting clock times. This is currently the case in the south of the country.^9^ Indeed, there is some evidence that rescheduling occurs in regions in close proximity to time zone borders elsewhere.^63^

### Perspectives

In terms of DST research, more studies are needed before we can draw strong conclusions regarding all health outcomes. The specifics of such studies should include location, determinants of individual sensitivities to changes in exposure timings relative to circadian phase (e.g., chronotype) and susceptibilities to adverse or beneficial health outcomes (e.g., individual baseline risks), and addressing the comparatively low number of high quality studies across several of the outcome domains. For instance, there is only one high quality study addressing all-cause mortality and one addressing traffic accidents with DST compared to Standard Time during summer months.^41^ ^64^ We noted in the Introduction that an intended benefit of DST during summer months is to increase available daylight in the evenings when (for many Western lifestyles) people are unconstrained by, for example, work or study commitments. It remains to be seen how well this is utilised and how it can best translate to a health perspective on DST vs. Standard Time during summer months. In terms of attaining higher quality studies, transitions can be seen as having sharp cut off points and time-based variation for which regression discontinuity designs – ideally with difference-in-differences analytical approaches and appropriate placebo tests – should be considered best practice.^30^ ^31 39 65–67^ Even if not using the difference-in-differences analytical approach, it is good practice to include placebo tests. Findings of transition effects on stock market decisions (as a proxy for cognitive performance) were falsified by use of such placebo tests (see the Cognitive Section of the Appendix). Such placebo tests can include assessing what would be expected if there were no transitions by using, for instance, a neighboring region or year wherein no transition takes place, or even a different date in the same year and place with no transition but close to the actual date of transition. Studies including such placebo tests should be the benchmark for future research on DST in order to deal with real-world complexity (of course, this can make data collection more difficult).

## Conclusion

In conclusion, outcome-specific detrimental effects, null effects, and beneficial effects of transitions and of living with DST compared to Standard Time during summer months are identified, synthesised, and assigned confidence ratings in this first systematic literature review across a range of DST-related exposures and outcomes. In addition to adverse effects, such as increased risk of myocardial infarction following DST-Onset, benefits of transitions and DST, such as decreased all-cause mortality and traffic accidents during summer months of DST compared to Standard Time and following DST-Offset, are detected. Whilst more high quality studies are called for, the current evidence regarding health impacts (which we acknowledge comes with limitations and is but one consideration when it comes to policy) can be taken as support for keeping the good and mitigating the bad that appears to be associated with DST.

## Data Availability

All extracted data is provided in the main text and appendix.

## Acknowledgements

We acknowledge the stimulus of the Sleep Health Foundation (Australia) in bringing together this team of researchers to conduct this project. However, the conduct of the study and the publication of the findings were independent of the SHF and the SHF did not review the manuscript prior to submission for publication.

## Contributions

Conceptualisation: TCE, RGF, GM; Methodology: AS, PL; Project Administration: AS; Literature search: AS, JPW, PL; Data interpretation (data extraction and writing appendix syntheses): AS, JPW, UW, LB, BB, GSL, SF, JW, GM, TCE, PL; Writing - original draft: PL; Writing - review & editing: All authors. Patients or the public were not involved in the design, or conduct, or reporting, or dissemination plans of our research.

## Ethical approval

Not required.

## Data sharing

All extracted data is provided in the main text and appendix.

## Transparency

All authors affirm that the manuscript is an honest, accurate, and transparent account of the study being reported; that no important aspects of the study have been omitted; and that any discrepancies from the study as planned (and, if relevant, registered) have been explained.

## Dissemination to participants and related patient and public communities

To ensure broad engagement, the findings of this systematic review have been, and will continue to be, presented at scientific conferences. Furthermore, we are committed to disseminating our findings widely to patient and public communities through strategic use of social media, the official websites of our respective organizations, and by informing sleep and health policy associations and societies.

