## Appendix for "A Systematic Review of Epidemiological Studies into Daylight-Saving Time & Health Identifying Beneficial & Adverse Effects"

#### Supplementary Material

### 1. Cardiovascular Section Synthesis

Jennifer Walsh & Gurprit S. Lall

##### *Introduction*

Systematic screening returned 22 articles that included outcomes relating to the cardiovascular system or treatments for cardiovascular events. The articles were further subcategorised into: (i) myocardial infarction (MI events, deaths, or interventions; 16 studies), (ii) cerebrovascular disease (events, deaths, or interventions; eight studies) and (iii) other cardiovascular outcomes (including, *inter alia*, atrial fibrillation, out of hospital cardiac arrest, acute coronary syndrome; seven studies). Five articles report multiple outcomes; thus, there is some overlap. All but one examined the effect of DST-Onset and/or DST-Offset. Most studies report outcomes over the initial one or two weeks post transition compared to reference periods, most include a substantial number of study years, and most include recent study years. The one different study examined the potential effect of longitudinal position in a time zone on stroke rates.

In this section, we first provide a summary of findings. Next, we provide more in-depth synthesis of studies concerning (i) MI with an overview of these studies in Cardiovascular-Table 1. Then we provide more in-depth synthesis of studies concerning (ii) cerebrovascular disease and (iii) other cardiovascular outcomes with an overview of these studies in Cardiovascular-Table 2. Lastly, we provide the quality assessment of studies using the Joanna Briggs Institute (JBI) Critical Appraisal Checklist for Quasi-Experimental Studies in Cardiovascular-Table 3.

##### *Summary*

Sixteen studies assessed occurrence of, treatment of, or death from MI (one study also included death from ischemic heart disease). Five of these studies are rated high quality and one is rated low quality. Increased risk of MI following DST-Onset appears more likely than not, with some individuals more susceptible than others. No effect following DST-Offset appears most likely. These findings include studies from Europe, USA, Mexico, and Brazil. This conclusion also fits with a recent meta-analysis on studies of transition effects on MI (which includes different quality ratings).[1] Six studies assessed occurrence of, or treatment of, or death from cerebrovascular disease following transitions, one study assessed the diurnal distribution of timing of stroke events following transitions, and one study assessed stroke outcomes by longitude. Only one medium quality study reported an increase in incidence of cerebrovascular disease following DST-Onset. The one high quality study reports no differences. Clearly, the strength of evidence for a possible effect of transitions on cerebrovascular disease is much lower than for MI. The study of longitude is in favour of perennial Standard Time (fewer events with more eastward position in time zone) but it is of too low quality to become part of the debate. Five of the six studies that assessed potential DST-Onset effects on other cardiovascular diseases report an increase in some outcomes for some individuals for specific days (too many to list specifics in this summary); two of these studies were rated high quality. The remaining study reported no difference. Three of the seven studies that assessed potential DST-Offset effects on other cardiovascular diseases reported a decrease in some outcomes for some individuals for specific days. The others report no differences.

Taken together, DST-Onset may be associated with an increase in some cardiovascular diseases and events in the days and/or week following transition, in particular MI, but this may not apply to cerebrovascular diseases. The majority of studies did not identify a consistent change in incidence of cardiovascular diseases associated with the DST-Offset transition.

##### *(i) Myocardial Infarction*

There are 16 studies concerning MI that can be further sub-grouped according to the outcome studied. Ten studies considered 'events' using registry data, four studies considered deaths *per se* (3 used registries and 1 used autopsy records; death from ischemic heart disease was also included as an MI outcome for one study and was also included in the synthesis here), and two studies considered percutaneous coronary intervention for treatment of MI using registry data. This subsection of the synthesis follows these groupings.

**Myocardial infarction events**

Of these ten studies, five report an increase for the whole week following DST-Onset. Four studies report no change for the first week or two weeks post transition (albeit two studies report increases in specific strata of the sample and/or on specific days). The remaining study assessed DST-Offset only.

The five studies reporting an increase for the whole week following DST-Onset include Culic (2016) in Croatia, two studies from Janszky et al. (2008, 2012) in Sweden, Jiddou et al. (2013) in the USA, and Zhang et al. (2020) who used data from the USA and Sweden. From 2,412 patients at a single University hospital in Croatia from 1990-1996, Culic (2016) report a 15% increase for the whole week and 29% for the Monday-Thursday post transition when compared to two weeks pre and the 2nd and 3rd week post transition.[2] The author accounted for transition day length and did not include the year wherein DST-Onset coincided with Easter. In two studies, Janszky et al. (2008, 2012) report a 5.1% and 3.9% increase for the week post DST-Onset.[3 4] In the 2008 study, they used data on 47,812 patients from a national Swedish register for the years 1987-2006 with the two weeks pre and the 2nd and 3rd weeks post transition as the reference period. In the 2012 study, they used data on 14,521 patients from the Swedish coronary care register for the years 1995-2007 and the week pre and the 2nd week post transition as the reference period. They also excluded study periods coincident with Easter and accounted for transition day lengths. In the second study, stratified analyses suggest a tendency for a more pronounced impact of DST-Onset in patients with lower total cholesterol, lower triglycerides, taking aspirin, and taking calcium channel blockers. In a study of 935 patients admitted to two US hospitals in 2006-2012, Jiddou et al. (2013) report a 17% increase for the week post transition compared to the two weeks pre and the 2nd and 3rd weeks post transition, with the largest increase of 71% on the transition Sunday.[5] The analyses accounted for transition day length and for Easter. The effect on non-ST-elevation MI (NSTEMI) was greater than on STEMI. In age and gender stratified analyses of a US insurance claim database for years 2003-2014 and a national inpatient register in Sweden for years 1968-2011, Zhang et al. (2020) report an ~1-2% increase for both sexes ≥61 years for the week post transition compared to one week pre and the 3rd post transition.[6] They accounted for transition day length and holidays.

Regarding DST-Offset in these five studies, Culic (2016) identified a 19% and 44% increase for the first week and for Mon-Thurs of the first week post transition, respectively.[2] Janszky et al. (2008, 2012) report no changes for the whole week in both studies and a 4.8% decrease for the Monday of the first week but only in the first study.[3 4] In the second study, patients with higher total cholesterol or taking statins or calcium channel blockers tended to have lower risk. Jiddou et al. (2013) report no change for the week but a 61% decrease for transition Sunday.[5] Zhang et al. (2020) report ~1-2% decrease in females ≥61 years and males 41-60 years for the week.[6]

The four studies reporting no change for the first week or the two weeks following DST-Onset include Kirchberger et al. (2015) in Germany, Sipila et al. (2016) in Finland, Rodriguez-Cortes et al. (2023) in Spain, and Mofidi et al. (2019) in Iran. Although Kirchberger et al. (2015) and Sipila et al. (2016) report no change for the first week, they do report differences in specific study sample strata and/or for a specific day of the week post transition. Kirchberger et al. (2015) studied data on 25,499 events in people aged 25-74 years from a German registry for years 1985-2010.[7] The analyses involved time series models using either the rest of year or months around transition and included adjustment for global time trend, temperature, humidity, barometric pressure, month, weekday and holidays. The 'taking ACE inhibitors prior to MI' and 'male' strata presented with increases of 49% and 16% in the first three days post transition compared to the reference period. A limitation is the restriction to a maximum age of 74 years. Sipila et al. (2016) examined 14,459 admissions to 22 Finnish hospitals with coronary intervention laboratories between 2001 and 2009, using the two weeks pre and the 2nd and 3rd week post transition as reference periods.[8] They report a 16% increase on the Wednesday post transition. Rodriguez-Cortes et al. (2023) report no difference when comparing the two weeks post to two weeks pre transitions in a study of 71,992 patients admitted to public hospitals in Andalusia in Spain for the years 2009-2019.[9] Data for the first vs second week post transition was not presented so it remains unclear whether a potential short-term effect was present. There was also no accounting for Easter, which limits the certainty of the findings. The study by Mofidi et al. (2019) compared two weeks post to two weeks pre transition in a study using data from five Iranian teaching hospitals from 2012.[10] The study was ranked low quality due to major discrepancies in the data and between data and text, using only a single transition, and very low sample size.

Regarding DST-Offset in these four studies, Kirchberger et al. (2015) report no differences for the first three days or whole week post transition.[7] However, an ~30% increase was observed for those who had previously suffered MI. Sipila et al. (2016) report no change for the whole week post transition but a 15% decrease on the Monday and a 15% increase on the Thursday.[8] Rodriguez-Cortes et al. (2023) report a 6% increase in the two weeks post compared to the two weeks pre transition (12% when considering NSTEMI alone).[9] No change was reported by Mofidi et al. (2019).[10]

The remaining study of the ten – by Jin et al. (2020) – reports a 0.0001% increase per day for four days (approximately 1 in 1 million people per day) following DST-Offset when compared to the two weeks pre and 2nd week post transition.[11] They studied 160 million hospital admissions (unclear how many MI) in Germany in the years 2000-2008. Their analysis included adjustment for season, day of week, public holidays, ambient temperature, and hours of sunshine.

Overall, most studies that considered DST-Onset report increases in the week post transition or in specific stratifications of their study sample and/or on specific days. Higher relative risks are also observed in some study sample stratifications. In addition to suggestion of higher risk among some individuals, there is also suggestion of a higher risk for NSTEMI compared to STEMI. The study by Jiddou et al. (2013) reports conspicuously high changes on transition days that are somewhat symmetric for DST-Onset and DST-Offset (71% increase and 61% decrease, respectively), but it is the only study to do this.[5] There may be some overlap in the studies that consider MI in Sweden given the use of national registries. While slightly different studies of similar datasets having similar results lends confidence to the results for Sweden, it may limit generalisability. Nonetheless, we may conclude that increased MI in the short-term following DST-Onset appears more likely than not. No clear picture emerges for DST-Offset with mostly null findings reported and no consistency among non-null findings.

**Myocardial infarction deaths**

Of the four studies which examined death/mortality registries, two used a regression discontinuity design framework and both identified an increase in MI deaths following DST-Onset. Goodwin et al. (2023) report a 4.5% increase in the ten days following DST-Onset in urban regions of Mexican states that utilise DST for the years 1998-2018.[12] No differences were observed in rural regions following DST-Onset and no differences were observed following DST-Offset. Toro et al. (2015) report a 7.4-8.5% increase in the days following DST-Onset in three states in Brazil that utilise DST for the years 2007-2012.[13] They did not study DST-Offset. The results in both studies held across various robustness and placebo tests.

Also using death registry data, Manfredini et al. (2019) report no differences in the week following either transition compared to the two weeks pre and the 2nd and 3rd week post transitions in the Veneto region of Italy for the years 2000-2015.[14] The outcome data included deaths from ischemic heart disease. Of note though, there was no correction for holidays and the study was likely underpowered; the authors report adequate power for detecting a 3% change in mortality for ~5,000 deaths in the index periods, which is approximately 2.5 times greater than the number of deaths identified to be attributed to ischemic heart disease during these periods.

The fourth study by Lindenberger et al. (2019) also reports no differences following transitions when analysing autopsy reports from one forensics centre in Germany for the years 2006-2015.[15] This includes deaths attributed to MI and a broader "cardiac" category. The absence of accounting for holidays and the very small sample limits the ability to draw robust conclusions from this study.

Overall, the two methodologically stronger studies indicate increased MI deaths following DST-Onset and no study reports differences following DST-Offset. Of note, the methodologically stronger studies conducted in Brazil and Mexico are compatible with the findings on MI events from studies in Europe and the USA.

**Percutaneous coronary interventions for myocardial infarction**

Sandhu et al. (2014) report 24% and 21% increases on the Monday and Tuesday following DST-Onset, respectively, but no differences for the whole week or following DST-Offset.[16] Their trend prediction model analysis included 42,060 patients undergoing intervention from all non-federal hospitals in one US state in the years 2010-2013. The findings held when models were adjusted for season and time-of-day. Derks et al. (2021) report no differences for the week post transition compared to the two weeks pre and 2nd week post transition for either DST-Onset or DST-Offset.[17] Their study included 12,751 patients undergoing intervention in the Netherlands in the years 2015-2018. They also observed no sex differences or differences between STEMI and NSTEMI. Overall, the two studies report different effects for DST-Onset and null-effects for DST-Offset.

Considering all 16 studies together, increased risk of MI following DST-Onset appears more likely than not, with some individuals more susceptible than others. No effect following DST-Offset appears most likely. This fits with a recent meta-analysis on studies of transition effects on MI.[1]

**Cardiovascular-Table 1: Overview of studies of myocardial infarction in order by quality rating and chronology**

| Author (Year) | Population/Data | Exposure/Comparator | Statistics | Results | R |
|---|---|---|---|---|---|
| Derks (2021)[17] | n=12,751 MI patients undergoing PCI, Netherlands Heart Registry | 1 week post vs 2 weeks pre & 2nd week post transitions, 2015-2018 | Multivariate Poisson regression | No differences for whole week or first 3 days for either men or women. | H |
| Kirchberger (2015)[7] | n=25,499 coronary deaths and non-fatal AMI, aged 25-74, MONICA/KORA Myocardial Infarction Registry, Germany | 1 week post vs whole year or vs months around transitions, 1985-2010 | Time series (generalized additive quasi-Poisson) models | DST-Onset: 49%↑ for those taking ACE inhibitors pre-MI & 16%↑ male strata for first 3 days. DST-Offset: 31%↑ prior MI stratum for first 3 days. | H |
| Culic (2013)[2] | n=2,412 AMI patients, one University Hospital in Croatia | 1 week post vs 2 weeks pre & 2nd & 3rd week post transitions, 1990-1996 | Multivariate regression | DST-Onset: 15%↑ for whole week, 29%↑ for Mon-Thurs, highest IR on Mon. DST-Offset: 19%↑ for whole week, 44%↑ for Mon-Thurs. | H |
| Jiddou (2013)[5] | n=935 AMI patients, aged>18yrs, 2 hospitals in Michigan (USA). Excl. pregnant women. | 1 week post vs 2 weeks pre & 2nd & 3rd weeks post transitions, 2006-2012 | IRs with 95%CIs based on $\chi^2$ and Poisson distributions | DST-Onset: 71%↑ for transition day, 17%↑ for whole week, ↑NSTEMI but ↓STEMI. DST-Offset: 61%↓ for transition day. | H |
| Janszky (2012)[4] | n=14,521 AMI patients, RIKS-HIA register, Sweden | 1 week post vs 2nd week pre & 2nd week post transitions, 1995-2007 | IRs with 95%CIs based on $\chi^2$ and Poisson distributions | DST-Onset: 3.9%↑ for whole week. DST-Offset: No differences. | H |
| Goodwin (2023)[12] | n=unknown*, fatality register, Mexico | Optimal bandwidth test, 1998-2018 | Regression discontinuity | DST-Onset: 4.5%↑ in urban areas stratum. DST-Offset: No differences. | M |
| Rodriguez-Cortes (2023)[9] | n=71,992 public hospital AMI admissions, Andalusian Minimum Basic Data Set, Spain | 1 week post vs 2 weeks pre & 2nd & 3rd weeks post transitions, 2009-2019 | Time series (with natural visibility graphs) models | DST-Onset: No differences. DST-Offset: 6%↑ (12%↑ NSTEMI) | M |
| Jin (2020)[11] | n=unknown*, MI cases in administrative hospital data from national statistics office, Germany | 1 week post vs 3 weeks pre & 2nd and 3rd post DST-Offset, 2000-2008 | Multivariate regression | DST-Offset: ↓~1 per 1 million population for first 4 days. | M |
| Zhang (2020)[6] | n= unknown* ischemic heart disease patients, USA MarketScan dataset and Swedish national inpatient register | 1 week post vs 1 week pre & 1 week post transitions, 1968-2011 (Sweden); 2003-2014 (USA) | Bayesian and frequentist relative risks with placebo tests | DST-Onset: ~1-2%↑ in both sexes ≥61 years for whole week. DST-Offset: ~1-2%↓ in females ≥61 years and males 41-60 years for whole week. | M |
| Lindenberger (2019)[15] | n=117 cardiac disease cases and n=30 MI cases from autopsy reports, Germany | 2 weeks post vs 2 weeks pre transitions, 2006-2015 | $\chi^2$ | No differences. | M |
| Manfredini (2019)[14] | n=7,207 ischemic heart disease deaths, Archive of Mortality Records, Veneto, Italy | 1 week post vs 2 weeks pre & 2nd & 3rd weeks post transitions, 2000-2015 | IRs with 95%CIs based on Poisson distribution | No differences. | M |
| Sipila (2016)[8] | n=14,459 MI admissions at 22 coronary catheterization laboratories, Finnish Care Register for Health Care | 1 week post vs 2 weeks pre & 2nd & 3rd weeks post transitions, 2001-2009 | IRs with 95%CIs based on $\chi^2$ and Poisson distributions, Cox regression | DST-Onset: 16%↑ for Wed. DST-Offset: 15%↓ for Mon, 15%↑ for Wed. | M |
| Toro (2015)[13] | n=34,596 AMI deaths, Mortality Information System, Brazil | Optimal Bandwidth test, 2007-2012 | Regression discontinuity | DST-Onset: ~8%↑. DST-Offset: No differences. | M |
| Sandhu (2014)[16] | n=42,060 AMI patients undergoing PCI, BMC2-PCI registry, Michigan USA | Trend model prediction for 1st week post transitions, 2010-2013 | Durbin-Watson test statistic | DST-Onset: 24%↑ for Mon. DST-Offset: 21%↓ for Tues. | M |
| Janszky (2008)[3] | n=47,812 AMI patients, Swedish AMI registry | 1 week post vs 2 weeks pre & 2nd & 3rd weeks post transitions, 1987-2006 | IRs with 95%CIs based on $\chi^2$ and Poisson distributions | DST-Onset: 5.1%↑ for whole week (↑for Mon-Wed). DST-Offset: No differences for whole week (4.8%↓ for Mon). | M |
| Mofidi (2019)[10] | n=142 AMI presentations 5 teaching hospital EDs in Iran | 1 week post vs 2 weeks pre & 2nd & 3rd weeks post transitions, 2012 | IRs with 95%CIs | No differences. | L |

(A)MI: (acute) myocardial infarction; PCI: percutaneous coronary intervention; (N)STEMI: (non-)ST elevation myocardial infarction; ED: emergency department; IRs: Incidence ratios; R: Quality Rating; L: low; M: medium; H: high; *actual number of cases used in specific analyses unknown

##### (ii) Cerebrovascular Disease

Eight studies assessed cerebrovascular disease outcomes, seven of which considered transitions and one of which – Xue et al. (1991) – considered longitude.[18] Of the seven studies of transitions, six examined relative risks and one – Foerch et al. (2008) – examined the diurnal distribution of symptoms and admissions.[19] Of the six that studied relative risk, five report no clear differences following DST-Onset and one study – Zhang et al. (2020) – reports a decrease.[6] None report a change in relative risk following DST-Offset. The study on diurnal distribution of timing is less relevant in terms of overall effect on health of the population compared to magnitude of differences following transitions so this will not be discussed further other than to say that advances and delays in timings are observed on Mondays and Tuesdays following DST-Onset and DST-Offset, respectively.[19]

Of the six studies that considered relative risk differences following transitions, four have been described already under (i) MI events. The other two studies are by Folyovich et al. (2020) and Sipila et al. (2016). Folyovich et al. (2020) report increased thrombolytic treatments on the first day post compared to the day pre and day of corresponding transitions using data from 12,630 records held by the Hungarian National Health insurance fund in the years 2006-2015.[20] The effect was not robust to excluding years wherein the index period included the last the day of the month (the authors report previously observing a striking increase in stroke on the last day of the month). Moreover, differences between Mondays, Sundays, and Saturdays may simply be day-of-week differences rather than transition effects. Sipila et al. (2016) report an 8% increase in stroke hospitalisations in Finland for the years 2004-2013 on transition Sunday and on the Monday post transitions when both biannual transitions are combined but no differences for the whole week or when DST-Onset and DST-Offset are analysed seperately.[21]

From the remaining four, Lindenberger et al. (2019) report no differences in cerebral events in their study of autopsies in Germany, Manfredini et al (2019) report no differences in cerebrovascular disease deaths in their study of mortality in the Veneto Region of Italy, and Rodriguez-Cortes et al. (2023) report no differences in stroke admissions in Andalusia in Spain.[9 14 15] Only Zhang et al. (2020) report an ~2% decrease in cerebrovascular diseases in both sexes following DST-Onset and no difference following DST-Offset in their study combining data from the USA and Sweden.[6]

Xue et al. (1991) report that with each five degree increase in longitude in China (from west to east) stroke incidence, prevalence, and mortality increased by ~22, 62, and 14 events per 100,000 inhabitants (for the year).[18] Rates were also impacted by rural/urban locations, time-of-year, and latitude.[18] The data came from a survey of approximately 5.8 million inhabitants (0.5% of total population) from 29 of 119 provinces in China on 01 January and 31 December 1986.

Overall, transition effects on cerebrovascular disease appears negligible. The single study of longitude is compatible with perennial Standard Time being preferable to perennial DST; however, as a single, low quality study, no conclusion can be drawn.

##### (iii) Other Cardiovascular Outcomes

Seven studies considered non-MI and non-cerebrovascular disease outcomes involving the cardiovascular system. Five studies are already discussed in other subsections; namely, Zhang et al. (2020) in the USA and Sweden, Jin et al. (2020) and Lindenberger et al. (2019) in Germany, Rodriguez-Cortes et al. (2023) in Spain, and Manfredini et al. (2019) in Italy. The two additional studies include Chudow et al. (2020) in the USA and Hook et al. (2021) in Australia.

Six of the seven studies assessed potential effects of the DST-Onset transition; only Lindenberger et al. (2019) reports no difference in outcome (autopsy identified cardiac disease).[15] Of the other three studies that are described in other subsections, Manfredini et al. (2019) report a 10% increase in circulatory deaths for the Tuesday post transition but no differences for the whole week.[14] Zhang et al. (2020) report a 1-2% increase in 'vasculature' and 'other heart diseases' in adults ≥ 61 years of age, and a 6% increase in 'other heart diseases' for males 11-20 years.[6] Rodriguez-Cortes et al. (2023) report no differences in 'major acute coronary events' but a 5% increase in acute coronary syndrome.[9] Of the two additional studies, Chudow et al. (2020) report a striking 25% increase in atrial fibrillation admissions at a single hospital in the USA for the Monday-Thursday post transition compared to the rest of the year for the years 2009-2016,[22] and Hook et al. (2021) report a 13% increase in cardiac arrests in adults > 18 years for the first two days post transition compared to the rest of the year in the years 2000-2020 using data from the Ambulance Cardiac Arrest Registry of one state in Australia.[23]

All seven studies considered DST-Offset. No differences were reported by Manfredini et al. (2019), Rodriguez-Cortes et al. (2023), Lindenberger et al. (2019), or Chudow et al. (2020).[9 14 15 22] The other three studies reported decreases in outcomes. Zhang et al. (2020) report 1-22% decreases in 'vasculature diseases' in adults aged ≥ 61 years and 'other heart diseases' in adults aged ≥ 40 years and Jin et al. (2020) report a 7.5% decreased admission rate for cardiac outcomes for the week post transition.[6 11] Hook et al. (2021) report a 12% decrease for the Tuesday post transition and a 30% decrease for the whole week.[23]

Overall, and taken together with the two other subsections, these findings are consistent with increased adverse cardiovascular outcomes following DST-Onset and no change following DST-Offset.

**Cardiovascular-Table 2: Cerebrovascular & other outcomes, sorted by outcome, quality, and chronology.**

| Author (Year) | Population/Data | Exposure/Comparator | Statistics | Results | R |
|---|---|---|---|---|---|
| **Cerebrovascular** | | | | | |
| Sipila (2016)[21] | n=14,834 ischemic stroke admissions, Finland | 1 week post vs 2 weeks pre & 2nd and 3rd weeks post transitions, 2004-2013 | Modified Poisson regression & Cox Regression. | No differences when transitions considered separately. | H |
| Rodriguez-Cortes (2023)[9] | n=58,327 public hospital stroke admissions, Andalusian Minimum Basic Data Set, Spain | 2 weeks post vs 2 weeks pre transitions, 2009-2019 | Time series (with natural visibility graphs) models | No differences. | M |
| Zhang (2020)[6] | n=unknown* cerebrovascular disease patients from USA MarketScan and Swedish National Inpatient Register | 1 week post vs 2nd week pre & 3rd week post transitions, 1968-2011 (Sweden); 2003-2014 (USA) | Bayesian and frequentist relative risks with placebo tests | DST-Onset: ~1-2%↓ both sexes ≥61 years and males 40-60 years. DST-Offset: no differences. | M |
| Lindenberger (2019)[15] | n=17 autopsy identified cerebral causes of death, 5 cities in Germany. | 2 weeks post vs 2 weeks pre transitions, 2006-2015 | $\chi^2$ test | No differences. | M |
| Manfredini (2019)[14] | n=4,722 deaths from 'cerebrovascular diseases', Archive of Mortality Records of the Veneto region, Italy | 1 week post vs 2 weeks pre & 2nd and 3rd weeks post transitions, 2000-2015 | IRs with 95%CIs based on Poisson distribution | No differences. | M |
| Foerch (2008)[19] | n=44,251 stroke patients with 'symptom onset time' and n=69,477 with 'admission time' from 100+ hospitals in Germany | Mon-Tue and Thur-Fri in the week post vs 1st or 1st-5th weeks pre transitions, 2000-2005 | Mann-Whitney U and Kruskal-Wallis tests | Advanced and delayed symptom-onset and hospitalisation times on Mon-Tue for DST-Onset and DST-Offset, respectively. | M |
| Folyovich (2020)[20] | n=12,630 thrombolytic treatment patients, National Health Insurance Fund, Hungary | First day, week, and month pre vs post transitions, 2006-2015 | Mann-Whitney U tests | No differences. | L |
| Xue (1991) [18] | n=~6million residents, various China provinces information on stroke. | Longitude, 1986 | Unspecified | ↑Incidence, prevalence and mortality by ~22, 61, and 14 per 100,000 inhabitants per 5° east. | L |
| **Other** | | | | | |
| Hook (2021)[23] | n=89,409 cardiac arrests, >18 years, (excl. from trauma, overdose, hanging), Ambulance Cardiac Arrest Registry, Australia. | 1 week post transitions vs rest of year, 2000-2020 | Interrupted time series multivariate regression | DST-Onset: 17%↑ for Sun-Mon. DST-Offset: 12%↓ for Tues, 30%↓ for whole week. | H |
| Jin (2020) [11] | n=unknown*, Administrative hospital data from national statistics office, Germany | 1 week post vs 3 weeks pre & 2nd and 3rd week post transitions, 2000-2008 | Multivariate regression | DST-Offset: 7.5%↓ for whole week. | H |
| Rodriguez-Cortes (2023)[9] | n=157,221 public hospital admissions, see above. | 2 weeks post vs 2 weeks pre transitions, 2009-2019 | Time series (with natural visibility graphs) models | DST-Onset: No differences in MACE. 5%↑ in ACS. DST-Offset: No differences. | M |
| Chudow (2020)[22] | n=6089 atrial fibrillation admissions, >18 years, single hospital, USA. | 1 week post transitions vs rest of the year, 2009-2016 | Wilcoxon non-parametric tests. | DST-Onset: 25%↑ for Mon-Thurs, 19%↑ for whole week. DST-Offset: No differences. | M |
| Zhang (2020)[6] | n=unknown*, see above. | 1 week post vs 1 week pre & 3rd week post transitions, 1968-2011 (Sweden); 2003-2014 (USA) | Bayesian and frequentist relative risks with placebo tests | DST-Onset: 1-2%↑ vasculature and other heart diseases ≥61 years, 6%↑ other heart diseases for males 11-20 years. DST-Offset: 1-2%↓ in vasculature diseases ≥61 years and other heart diseases ≥40 years. | M |
| Lindenberger (2019)[15] | n=117 autopsy identified cardiac disease, 5 cities in Germany | 2 weeks post vs 2 weeks pre transitions, 2006-2015 | $\chi^2$ test | No differences. | M |
| Manfredini (2019)[14] | n=20,638 'circulatory deaths', Archive of Mortality Records, Veneto, Italy | 1 week post vs 2 weeks pre & 2nd and 3rd weeks post transitions, 2000-2015 | IRs with 95%CIs based on Poisson distribution | DST-Onset: 10%↑ for Tues. DST-Offset: No differences. | M |

MACE: major adverse cardiovascular event; ACS: acute coronary syndrome; R: Quality rating; L: low; M: medium; H: high; *actual number of cases involved in specific analyses unknown; IRs: Incidence ratios

**Cardiovascular-Table 3: JBI Quality Indicators** (Questions and possible answers are shown in detail in Table footnote)

| Author (Year) | Q1 | Q2 | Q3 | Q4 | Q5 | Q6 | R | Notes |
|---|---|---|---|---|---|---|---|---|
| Chudow (2020)[22] | Y | U# | U& | Y | Y | N | M | #Supporting data for no differences in characteristics including medical conditions/treatments between exposure and control populations is not presented. &Known seasonal variation in prevalence of atrial fibrillation is acknowledged but is not controlled for in analyses. |
| Culic (2013)[2] | Y | Y | Y | Y | Y | N | H | |
| Derks (2021)[17] | Y | Y | U# | Y | Y | N | H | #Information about pre-existing medical treatments is not provided. |
| Foerch (2008)[19] | Y | U# | U& | Y | Y | N | M | #Population characteristic data not included. & Information about pre-existing medical conditions/treatments is not provided |
| Folyovich (2020)[20] | Y | U# | U& | Y | U* | Y | L | #Population characteristic data, &information about pre-existing medical conditions/treatments, and *mean data variance not provided. *Differences in 'mean daily number' of treatments for 1 day (e.g., 0.9), 1 week (e.g., 1.86) and 1 month (e.g., 3.39) values not explained. |
| Goodwin (2023)[12] | Y | U# | U& | Y | Y | N | M | #Population characteristic data is not included. &Information about pre-existing medical conditions/treatments is not provided. |
| Hook (2021)[23] | Y | Y# | U& | Y | Y | N | M | #Limited population characteristic data is provided. &Information about pre-existing medical conditions/treatments is not provided. |
| Janszky (2012)[4] | Y | Y | Y | Y | Y | N | H | |
| Janszky (2008)[3] | Y | U# | U& | Y | Y | N | M | #Population characteristic data is not provided. &Information about pre-existing medical conditions/treatments is not provided. |
| Jiddou (2013)[5] | Y | Y | Y | Y | Y | N | H | |
| Jin (2020)[11] | Y | U# | U& | Y | Y | N | M | #Population characteristic data is not included. & Information about pre-existing medical conditions/treatments is not provided. |
| Kirchberger (2015)[7] | Y | Y# | Y* | Y | Y | N | H | #Some differences in population characteristics but were evaluated in subgroup analyses. *Some population differences in CV treatments but were evaluated in subgroup analyses. |
| Lindenberger (2019)[15] | Y | U# | U& | Y | Y | N | M | #Population characteristic data is not included. &Information about pre-existing medical conditions/treatments is not provided. |
| Manfredini (2019)[14] | Y | U# | U& | Y | Y | N | M | #Population characteristic data is not included. &Information about pre-existing medical conditions/treatments is not provided. |
| Mofidi (2019)[10] | Y | U# | U& | Y | U* | Y | L | #Limited population characteristic data and no statistical comparisons presented. &Limited information about pre-existing medical conditions/treatments and no statistical comparisons presented. *Methods seem appropriate but results are not (extremely tight CIs for small sample size). *Inconsistency between sample size data in the table (from calculation) and text is present. |
| Rodriguez-Cortes (2023)[9] | Y | U# | U& | Y | Y | N | M | #Population characteristic data is not included. &Information about pre-existing medical conditions/treatments is not provided. |
| Sandhu (2014)[16] | Y | U# | U& | Y | Y | N | M | #Population characteristic data is not included. &Information about other medical conditions/treatments is not provided. |
| Sipila (2016a)[8] | Y | U# | U& | Y | Y | N | M | #There were differences in exposure and comparator groups (e.g., diabetes and renal failure (DST-Onset only)), which were not controlled for. &Information about pre-existing medical conditions/treatments is not provided. |
| Sipila (2016b)[21] | Y | Y | U& | Y | Y | N | H | &Information about pre-existing medical conditions/treatments is not provided. |
| Toro (2015)[13] | Y | U# | U& | Y | Y | N | M | #Population characteristic data is not included. &Information about pre-existing medical conditions/treatments is not provided. |
| Xue (1991)[18] | X | U# | U& | Y | U* | N | L | #Population characteristic data and &information on pre-existing medical conditions/treatments not provided. *Statistics not provided. |
| Zhang (2020)[6] | Y | U# | U& | Y | Y | N | M | #Population characteristic data is not included. &Information about pre-existing medical conditions/treatments is not provided. |

R=Quality Rating

**JBI Critical Appraisal Checklist for Quasi-Experimental Studies (Adapted)**

Answers can be either Y=Yes, N=No, U=Unsure, or N/A.

1. Is it evident which factor is considered the 'cause' and which one is the 'effect'? Is there no ambiguity regarding the sequence of variables under investigation?
2. Were the characteristics of the study participants involved in comparisons adequately matched?
3. Were the exposures/treatments (beyond that of interest in our study) of the study participants involved in comparisons adequately matched?
4. Were the methods used to measure the outcomes the same for participants who were part of different comparisons?
5. Were appropriate statistical methods employed to analyse the data?
6. Are there any irregularities in the data reported?

#### 2. Psychiatry Section Synthesis

Ben Bullock & Greg Murray

##### *Introduction*

Twelve studies investigated psychiatric symptoms and disorders. The studies represent a range of populations (patients, students, adolescents, adults), locations (North America, Europe, Russia), and psychiatric symptoms and disorders (mania, depression, dementia, Parkinson's disease, seasonal affective disorder, suicide). Most studies reported on acute effects of both DST-Onset and DST-Offset transitions. An overview of included studies is presented in Psychiatry-Table 1 and JBI quality indicators in Psychiatry-Table 2.

Overall, the evidence does not support a robust association between transitions and psychiatric outcomes. Five studies report no significant associations. The remaining seven studies report weak and inconsistent associations that are limited to specific subpopulations. In the following paragraphs, we describe (i) the five studies that find no associations, then (ii) the seven studies that report weak and inconsistent associations.

##### *(i) Studies reporting null associations*

Fetter et al. (2014) found no differences in depressive symptoms, psychosis, or severity of "wearing off" symptoms following transitions among 83 patients with Parkinson's Disease.[24] The pre-test – post-test design emphasises the matching of samples and is a strength of this French study. However, it is also limited by a small sample recruited from a single clinic and its reliance on patients' self-reported symptoms as outcome measures.

Three hospital records-based studies found no evidence of a change in psychiatric status across transitions. Only one of these considers relevant control variables like age and sex. In such registry studies, the distributions of these variables are unlikely to shift significantly in the catchment area population pre and post transition, but including these control variables will enhance confidence in any observed effects. Lahti et al. (2008) compared hospital discharge numbers following hospitalisation for mania in the two weeks pre to the two weeks post both DST-Onset and DST-Offset.[25] This study assessed transitions at over many years (1987-2003) and across all hospitals in Finland (private, public, general, mental, military, prison). These design features, along with the use of formal ICD-9 and ICD-10 diagnoses to identify relevant cases and the employment of relevant control variables (e.g., age and sex), constitute a strong test of transition-related effects on psychiatric status.

The second records-based study, by Heboyan et al. (2019), assessed 6,932 emergency department presentations at two weeks pre and post both DST-Onset and DST-Offset for a range of mental health conditions (e.g., mania, depression, dementia) at a hospital in south-eastern USA.[26] The large number of cases for analysis and the use of formal ICD-9 diagnoses for case identification are significant strengths of this study, but a lack of statistical control over co-occurring and/or pre-existing conditions and treatments is a limitation. Importantly, gender, race, age, and health insurance were employed as control variables in the analysis. The location – latitude 33°, close to the equator and therefore subject to less dramatic changes in daylight throughout the year – means the findings may not be generalisable to locations at higher latitudes.

The third study, by Nixon et al. (2021), assessed 2,498 inpatient psychiatric admissions at a paediatric hospital in Canada.[27] Admission rates in the week pre transition were compared with rates in the first week post transition and with rates in the second week post transition for both DST-Onset and DST-Offset. The young sample (mean age of 16.32 years) is a unique feature of this study, and the use of a clinician-rated measure of symptoms and functioning (Childhood Acuity of Psychiatric Illness Scale) at admission constitutes a design strength. Failure to consider potentially important covariates such as age, sex, and pre-existing psychiatric diagnoses are limitations. The schooling situation of the young sample also introduces a potential confound (acknowledged by the study authors) in that DST-Onset occurred simultaneously with a break in schooling. The break may have compensated for any transition effect on psychiatric disorder presentations. In other words, assuming school increases the number of presentations (e.g., a mechanistic hypothesis being school times reduce sleep duration, which is a transdiagnostic feature for psychiatric episode onset), presentations would be expected to decrease with school break if the same index and reference periods were compared but without the presence of a transition. If there is no effect of school on admissions, then transition *per se* has no effect on psychiatric admissions in the study population.

Lastly, Shapiro et al. (1990) assessed Scottish government records of completed and attempted suicides and found no significant difference in the mean number of cases between the week pre transition and the first week post transition for both DST-Onset and DST-Offset.[28] Like many of the other records-based studies included in this section, the analyses failed to statistically control for potentially important covariates such as pre-existing conditions and/or treatments. Unlike many of the other studies in this section, Shapiro et al. did not statistically control for important demographic features such as age and sex.

##### *(ii) Studies reporting weak and/or inconsistent associations*

A study from Australia by Berk et al. (2008) showed a small but statistically significant increase in male suicides within 2-4 weeks of DST-Offset compared to the rest of autumn.[29] No change in suicide numbers was found within 2-4 weeks of DST-Onset compared to the rest of spring. Similarly, no change in suicide numbers was found for females at either DST-Onset or DST-Offset, although this may be due to the low number of cases for analysis in this study. This study includes a large number of documented and confirmed cases of suicide across a significant period of time (1971-2001), however the effects of transitions are small, and occur in subgroups only (i.e., the significant difference for males at 2-4 weeks post DST-Offset was only found post-1986 with no reason given for this timeline).

An American study by Osborne-Christenson et al. (2022) showed a 6.25% increase in suicides on the day of DST-Onset compared to other days of the year.[30] No meaningful change was observed in the number of suicides (0.53%) on the day of DST-Offset. The effect size at DST-Onset was reported to be stronger in white males aged 45-54 years, but no other subgroup analyses were reported. Also, the 10-year period analysed for this study (1979-1988) was selected from a potential pool of 70 years of data with no justification provided for this selection. Nevertheless, this was a seemingly robust analysis of a large number of cases of suicide and one which controlled for several evidence-based covariates (e.g., wealth, income, fluctuations in the business cycle, weather).

Investigating a small local register of 63 autopsies with a determination of suicide in Germany, Lindenberger et al. (2019) reported a non-significant ($p = 0.07$) increase in the number of suicides in the two weeks post DST-Onset compared to other times of the year.[15] No significant changes in suicide numbers were reported following DST-Offset. The small sample of records precluded analysis of potentially important covariates (e.g., age, sex, pre-existing psychiatric diagnosis/treatment) and limits the strength of conclusions that can be drawn from the results of this study.

Zhang et al. (2020), using two huge databases of health insurance claims for mental/behavioural disorders, one from the USA (150 million patients) and one from Sweden (9 million patients), showed a statistically significant increase in one specific condition (psychoactive substance use, and only in men aged 20+ years) at both DST-Onset and DST-Offset.[6] The observed effects were small and, due to the data source, apply only to the insured population. Considering these small effects, and the fact that co-morbidity of other physical and mental disorder diagnoses was not controlled in statistical modelling, the results of the analyses must be considered tentative at this stage.

Using data from a national register of 277,599 hospital presentations for unipolar depression and bipolar disorder in Danish hospitals, Hansen et al. (2017) showed an 11% increase in the observed number of unipolar depression diagnoses at DST-Offset compared to expected diagnosis numbers based on long-term averages.[31] No change at DST-Onset, nor in bipolar disorder diagnoses at either transition was found in this study. The large number of observations for analysis is a strength of the study. That the data were collected over a significant period of time (1995-2012) may be considered both a strength and a limitation of the study, especially as diagnoses were based on principles of "everyday clinical practice", and such practice may be subject to variations in diagnostic trends over time (acknowledging that the inpatient and emergency room setting precludes strict adherence to research-based diagnostic criteria.). The lack of statistical control over co-occurring conditions, pre-existing treatments, and other non-DST-related factors is also a limitation.

Two final studies reported weak associations between depression and transitions, each employing a distinctive design to investigate hypothesised effects. Borisenkov et al. (2017) capitalised on the unique circumstance of multi-year transitions between seasonal DST, perennial DST, and perennial non-DST across an 8-year period (2009-2016) in northern Russia.[32] Self-reported winter depression was observed at higher rates under perennial DST conditions than under non-DST conditions. Jankowski et al. (2014) on the other hand, used two locations in Europe in the same time zone, but in which sunrise and sunset occurred one hour earlier in one location (Warsaw, Poland) than the other (Heidelberg, Germany), which could be considered as a proxy measure of Standard Time vs. DST, respectively.[33] Higher self-reported depression scores were found in the more eastern location of Warsaw compared to Heidelberg. Limitations of self-report data aside, the reported effect sizes were small in both studies. Also noticeable were the low average scores for depression (<10) on the CES-D measure at both sites in the Polish/German study.

##### *Conclusion*

A range of study designs were used, most of which relied on retrospective analysis of data. The most commonly used sources of data were government/hospital records (suicide, formal psychiatric diagnoses). Investigations typically compared case numbers in the period following DST-Onset and DST-Offset with case numbers at other times of the year. Stronger in design were those studies that compared rates of psychiatric diagnoses and/or suicide pre transition with post transition (e.g., Lahti et al. 2008[25]) because they provide a more direct assessment of the transition 'effect'. Analyses based on government/hospital records benefit from large numbers of cases collected over extended periods of time and a higher standard of psychiatric diagnosis compared to studies using self-reports. Such large numbers permit more complex statistical analyses and more powerful investigations of comparatively low-frequency events, such as suicide. Of course, a corollary of more powerful statistical analysis is potential identification of statistically significant effects that are of small magnitude. Small effect sizes were a feature of many studies included in the review.

It is noteworthy that analyses based on general and mental health hospital registers are limited to investigating those cases that present to hospital departments, usually in crisis. Cases with relatively milder presentations are probably more likely to attend their local doctor or other privately employed health practitioner. These milder, yet still distressing experiences, are not captured in reported data. It may indeed be the case that milder presentations are more common following transitions than full-blown psychiatric episodes.

In conclusion, robust evidence for an effect of transitions on psychiatric status is not apparent in the reviewed literature. This overall finding does not preclude the possibility of a meaningful effect. Moreover, if there is a meaningful effect, the current literature would suggest it is likely to be of very small effect size and limited to specific conditions (unipolar depression, psychoactive substance use) and suicide among middle-aged men.

**Psychiatry-Table 1: Overview of studies, in order by text subcategorization, quality rating, and chronology**

| Author (Year) | Population/Data | Exposure/Comparator | Statistics | Results | Rating |
|---|---|---|---|---|---|
| ***Studies reporting null associations*** | | | | | |
| Nixon (2021)[27] | Inpatient psychiatric admissions, Canada; n=2,498 (69.7% female), 16.32 ± 1.66 years. | 1 week pre vs 1st & 2nd weeks post transitions, 2012-2017 | One-Way Repeated measures ANOVA | No difference in number of psychiatric admissions. | M |
| Heboyan (2019)[26] | Database of emergency department hospital presentations, USA, n=6,932 (54% male), age range 18-65 years | 2 weeks pre vs 2 weeks post transition vs Standard Time, 2013-2015 | $\chi^2$/Fisher's Exact test; Multinomial logistic regression | No differences in hospital presentations. | M |
| Fetter (2014)[24] | Patients attending Parkinson's Disease clinic, France, n=83 (52 male) | 3 days pre vs 3 days post transitions, 2011-2012 | Student t-test | No difference in "wearing off" time, depressive symptoms, or psychosis. | M |
| Lahti (2008)[25] | Hospital discharge register, Finland | 2 weeks pre vs post transitions, 1987-2003 | Poisson regression | No effect on manic episodes. | M |
| Shapiro (1990)[28] | (i) Scottish register of parasuicides, n=1170, (ii) Scottish hospital records of psychiatric admissions, n=4722, (iii) Scottish government record of completed suicides, n=4734 | 1 week pre vs 1 week post transitions, (i) 1962-1987, (ii) 1970-1987, (iii) 1974-1983 | $\chi^2$; ANOVA | No difference in case numbers for any cohort. | L |
| ***Studies reporting weak and/or inconsistent associations*** | | | | | |
| Osborne-Christenson (2022)[30] | National Center for Health Statistics (death by suicide or drug/alcohol overdose), n=87 deaths per day, USA | 1 day post transitions vs other days in the year, 1979-1988 | Regression discontinuity analysis | ↑Suicides on DST-Onset day only (6.25%). | M |
| Lindenberger (2019)[15] | Autopsy Register, Institute of Medicine, Goethe University, Frankfurt/Main-Germany; n=63 | 2 weeks post transitions vs other times of year, 2006-2015 | $\chi^{2}$* | No significant differences in suicides. | M |
| Borisenkov (2017)[32] | Students enrolled in schools/universities, Russia, n=7,968 | Seasonal DST (April-October, 1/1/2009-26/3/2011) vs. perennial DST (whole year, 27/3/2011-26/10/2014) vs. perennial non-DST (whole year, 27/10/2014-31/12/2016) | One-way ANCOVA; $\chi^2$ | ↑Seasonal Affective Disorder (Winter depression) in perennial DST vs. non-DST periods. | M |
| Hansen (2017)[31] | National register of unipolar depression (n=185,419) and bipolar disorder (n=92,180) primary diagnoses reported in hospital inpatient and emergency room settings, Denmark | 1 week pre vs 1 week post transitions, 1995-2012 | Time series transfer function comparing effect of transitions to expected effect based on "natural" or "unperturbed" process | ↑Unipolar depression diagnosis post DST-Offset only (11%). | M |
| Zhang (2020)[6] | USA MarketScan (n>150 million), Swedish national inpatient register (n>9 million) | 1 week post vs 2 weeks pre and 2nd week post transitions, 2003-14 (USA) & 1980-2011 (Sweden) | Bayesian and frequentist relative risks, including use of "sham"/negative controls | ↑Relative risk of psychoactive substance use at both DST-Onset and -Offset (in males aged 20+ only). | L |
| Jankowski (2014)[33] | University students from Poland, n=291 (82.8% female) and Germany, n=279 (77.8% women), age range 18-28 years | Longitudinal positions in time zone (56-minute sun phase difference), 2012 | ANCOVA | ↑Depression in Poland sample (advanced sun time). | L |
| Berk (2008)[29] | Government register of completed suicides, Australia, n=61,598 (76.6% male) | 2-4 weeks pre vs 2-4 weeks post transitions, 1971-2001 | Two-Way ANOVA | ↑Suicides following DST-Offset only (men only). | L |

L=low, M=medium, H=high. *comparing with expected number of deaths based on mean observed throughout the rest of the year.

**Psychiatry-Table 2: JBI Quality Indicators**
(Questions and possible answers are shown in detail in Table footnote)

| Author (Year) | Q1 | Q2 | Q3 | Q4 | Q5 | Q6 | Rating | Notes |
|---|---|---|---|---|---|---|---|---|
| Berk (2008)[29] | N | N | N | Y | U | Y | L | |
| Borisenkov (2017)[32] | Y | N | N | Y | Y | N | M | |
| Fetter (2014)[24] | Y | Y | Y | Y | U | N | M | |
| Hansen (2017)[31] | Y | N | Y | U* | Y | N | M | *ICD-10 used, but unsure whether criteria were applied similarly across all cases. |
| Heboyan (2019)[26] | Y | N | Y | Y | Y | Y* | M | *The reported number of mental and behavioural health visits in Table 1 does not match number reported in-text. |
| Jankowski (2014)[33] | Y | N | U | Y | Y | N | L | |
| Lahti (2008)[25] | Y | N | N | Y | Y | N | M | |
| Lindenberger (2019)[15] | Y | N | N | Y | Y | N | M | |
| Nixon (2021)[27] | Y | N | N | Y | Y | N | M | |
| Osborne-Christenson (2022)[30] | Y | N | N | Y | Y | N | M | |
| Shapiro (1990) | Y | N | N | N | Y | N | L | |
| Zhang (2020)[6] | N | N | N | Y | Y | N | L | |

**JBI Critical Appraisal Checklist for Quasi-Experimental Studies (Adapted)**
Answers can be either Y=Yes, N=No, U=Unsure, or N/A.

1. Is it evident which factor is considered the 'cause' and which one is the 'effect'? Is there no ambiguity regarding the sequence of variables under investigation?
2. Were the characteristics of the study participants involved in comparisons adequately matched?
3. Were the exposures/treatments (beyond that of interest in our study) of the study participants involved in comparisons adequately matched?
4. Were the methods used to measure the outcomes the same for participants who were part of different comparisons?
5. Were appropriate statistical methods employed to analyse the data?
6. Are there any irregularities in the data reported?

### 3. Traffic Accidents Section Synthesis

Aiste Steponenaite & Lorna Brown

***Introduction***

This section is subdivided into (i) fatal traffic accidents, (ii) all traffic accidents (incl. fatal accidents), and (iii) traffic accident-linked hospital admissions. These are further subdivided into USA-based and non-USA based studies to break up the synthesis for clarity and ease of reading, comparison, and assimilation. There are 23 studies included in total and some are considered in multiple subdivisions. A tabulated overview of included studies is presented in Traffic-Tables 1-3 (corresponding to subdivisions (i) to (iii); some studies included in more than one table as applicable). JBI quality indicators are presented in Traffic-Table 4. Individual study effect sizes are presented in tables and mentioned in the conclusion paragraph at the end of the section. In the all traffic accidents sub-division, there are some studies that separate e.g., those involving motor vehicles from those involving pedestrians; in such cases, only those involving motor accidents are considered. A limitation of the all traffic accidents subdivision is that not all accidents may have been severe enough to be reported to police or to registers. This should be kept in mind when reading and evaluating this subdivision synthesis.

***(i) Fatal Traffic Accidents***

Fatal accidents include 13 studies, of whom eight are conducted in the USA, two in the UK, one in Spain, and one in Mexico. From the USA, one is rated high quality,[34] one as low quality,[35] and six as medium quality.[36-41] The one study from Australia is rated high quality,[42] the one study from Spain as low quality,[43] and the two UK-based studies and one Mexico-based study as medium quality.[44 45]

***USA based***

The USA-based high-quality study from Fritz et al. (2020) reports increased fatal accidents on the Sunday, Monday, and Monday to Friday (pooled) following DST-Onset compared to all other corresponding days of the year for the period 1996-2017 and across all of the USA.[34] They report no differences following DST-Offset.

Of the six medium quality studies from the USA, three use data from across the USA[38 39 41] and the other three from Florida,[37] Texas,[40] and Minnesota,[36] respectively. Across the USA, Smith et al. (2016) find increased fatal accidents for the period 2002-2011 following DST-Onset but not DST-Offset (using regression discontinuity design and placebo tests assuming different transition days).[38] There is a clear discontinuity on the regression day but it is unclear how quickly the fatal accidents return to following the seasonal trend (i.e., increasing). Sood et al. (2007) find decreased fatal accidents for the period 1987-2003 following DST-Onset but did not study DST-Offset. Placebo tests using non-DST years (1976-1986) suggest the DST-Onset finding could be a false positive. Lastly, their empirical strategy involved standardising log weekly data in their DST years to non-DST years.[39] Varughese et al. (2001) find increased fatal accidents on the Monday but not on the Sunday (compared to corresponding days in the weeks pre and post) for the period 1975-1995 following DST-Onset and vice versa for DST-Offset.[41] In Florida, Molina et al. (2022) observe increased fatal accidents on

Thursday post DST-Onset compared to the corresponding day in the week pre DST-Onset and no differences on other days for the period 1983-2019 using a Bayesian analysis.[37] Decreased fatal accidents were observed for the Sunday, Monday, and Tuesday following DST-Offset. In Texas, Stevens et al. (2006) report no change in fatal accidents post DST-Onset and an increase following DST-Offset for the period 1998-2000.[40] In Minnesota, Huang et al. (2010) report no change in fatal accidents on the Sunday or Monday following DST-Onset or DST-Offset compared to corresponding days in the reference weeks pre and post transitions for the period 2001-2007.[36]

The one low quality study by Hicks et al. (1998) reports increased fatal accidents associated with alcohol for the period 1989-1992 in New Mexico for the combined first week post transitions compared to the combined weeks pre transitions.[35]

Overall, increased fatal accidents in the initial days post DST-Onset are observed in four studies that include the most recently collected data,[34 37 38 41] three of which are from nationwide datasets,[34 38 41] and these also include the only high quality rated study.[34] The findings are not completely similar, with the days on which significant effects are observed differing across studies. Nonetheless, it would be remiss to conclude no effect of DST-Onset on increasing traffic accidents on at least some of the initial days post DST-Onset. Regarding DST-Offset, these same four studies report no change,[34 38] decreased,[37] and increased fatal accidents (the latter on the transition Sunday only);[41] the study with decreased accidents being from Florida only. The other two studies that assess DST-Offset report no change and an increase in fatal accidents in the initial days post transition.[36 40] Thus, both transitions may increase fatal accidents, at least on some of the initial days post transition, but this appears more likely for DST-Onset.

***Non-USA based***

The high-quality rated study from Australia by James et al. (2023) for the period 1989-2015 reports no differences in the two weeks post DST-Onset or DST-Offset when comparing states that implement DST to those that do not.[42] Also, no difference is observed when comparing 40 days pre and post transitions. In the UK-based medium quality studies, Singh et al. (2022) find no change in fatal accidents following DST-Onset or DST-Offset overall for the period 2005-2018, and decreases in few spatial- and temporal- specific models using a regression discontinuity design.[45] Bunnings et al. (2021) also report no change in fatal accidents following DST-Onset and DST-Offset for the period 1996-2017 using the regression discontinuity design.[44] In the Mexico-based medium quality study by Goodwin et al. (2024), increased fatal accidents are observed following DST-Onset (for rural areas in particular) and DST-Offset (for urban areas in particular) for the period 1998-2018 using a regression discontinuity design.[12] The low quality rated study from Spain by Prats-Uribe et al. (2018) finds increased fatal accidents on DST-Onset and DST-Offset transition days (less so for DST-Offset) compared to all other days for the period 1990-2014.[43] They also report a potential harvesting effect (i.e., a short period of excess outcome followed by a compensating deficit, possibly indicating that the events might have occurred shortly after index periods anyway).

Overall, the findings from studies in Spain and Mexico appear more similar to those from the USA. From the studies in Australia and the UK, little-to-no effect of transitions or living with DST compared to Standard Time during summer months on fatal traffic accidents are apparent. Thus, the conclusion that both transitions may increase fatal accidents, at least on some of the initial days post transition and in some places, remains (i.e., there may be effect modification present in some places and at some times).

**Traffic-Table 1: Overview of n=13 studies of fatal traffic accidents, in order by text subcategorization, quality rating, and chronology**

| Author (Year) | Population/Data | Exposure/Comparator | Statistics | Results | Rating |
|---|---|---|---|---|---|
| ***USA based*** | | | | | |
| Fritz (2020)[34] | USA, n=732,835 fatal motor vehicle accidents. | Monday-Friday pre vs post transitions, 1996-2017 | Poisson regression Benjamini-Hochberg procedure to correct multiple comparisons P values | DST-Onset: 6% ↑ fatal accidents.<br>DST-Offset: no change. | H |
| Molina (2023)[37] | USA (Florida), Crash analysis reporting system, n=unknown | 1 week pre vs 1 week post transitions, 1983-2019 | Paired Wilcoxon rank test implemented using a Bayesian approach. | DST-Onset: ↑ fatal accidents.<br>DST-Offset: ↓ fatal accidents. | M |
| Smith (2016)[38] | USA, FARS, n=unknown. | Optimal bandwidth selector pre and post transitions, 2022-2011 | Regression discontinuity design with local linear regression specification. Day-of-year Fixed effects model. | DST-Onset: ↑ 5-6.5% in fatal accidents.<br>DST-Offset: no change. | M |
| Huang (2010)[36] | USA (Minnesota), Minnesota Office of Traffic Safety, n=unknown. | 8 weeks pre vs 8 weeks post, first Sunday and first Monday post transitions vs all others over 16 weeks, 2001-2007 | Deviance and Pearson $\chi^2$ | DST-Onset: no change.<br>DST-Offset: no change. | M |
| Sood (2007)[39] | USA, FARS, n=unknown. | 13 weeks pre vs 9 weeks post transition, transition years (1987-2003) including control, non-DST years (1976-1986) | Empirical model to transform data and conventional difference-of-difference estimator framework | DST-Onset: 9-10% ↓ fatal accidents.<br>Monday Data: 13% ↑ during DST years and 10% ↑ during control years compared to other Mondays in the month. | M |
| Stevens (2006)[40] | USA (Texas), Fatal (K), incapacitating (A), and non-incapacitating (B) accidents (KAB), n=157,290 | 5 workdays pre vs 5 workdays post transition, 1998-2000 | Generalised linear models with an additional naïve before-and-after study. | DST-Onset: no change.<br>DST-Offset: 17% ↑ fatal accidents. | M |
| Varughese (2001)[41] | USA, The US National Highway Transportation Safety Administration, n=unknown. | Saturday, Sunday, Monday of 1 week pre and 2 weeks post vs 1 week post, 1975-1995 | One-tailed & two-tailed paired t-tests | DST-Onset: ↑ on Monday.<br>DST-Offset: ↑ on Sunday. | M |
| Hicks (1998)[35] | USA (New Mexico), All fatal traffic reports n=204. | 1 week pre vs 1 week post combined transitions, 1989-1992 | t-tests | ↑ in fatal (alcohol-related) accidents | L |
| ***Non-USA based*** | | | | | |
| James (2023)[42] | Australia, n=41,000, Australian Road Deaths database. | 2 weeks and 40 days pre vs post transitions, 1989-2015 | Regression Discontinuity in Time and day-of-year fixed effects model. | DST-Onset: no change.<br>DST-Offset: no change. | H |
| Goodwin (2023)[12] | Mexico, n=41,495 automobile fatalities | Optimal bandwidth selector pre and post transitions, 1998-2018 | Regression discontinuity design | DST-Onset: ↑ 14.4% fatal accidents.<br>DST-Offset: ↑ 12.6% fatal accidents. | M |
| Singh (2022)[45] | Great Britain, UK Department for Transport, n=5,429 fatalities. | Optimal local bandwidth selector pre and post transitions, 2005-2018 | Regression discontinuity design analysis. | DST-Onset: no change.<br>DST-Offset: no change. | M |
| Bunnings (2021)[44] | UK Public Road Accidents, n>4.1 million | Mean squared error optimal bandwidth pre and post transitions, 1996-2017 | Regression discontinuity design, considering weather variables, time of day effects | DST-Onset: 8.9-9.9% ↑ fatal accidents.<br>DST-Offset: 1.3-5.1% ↑ fatal accidents. | M |
| Prats-Uribe (2018)[43] | Spain, Spanish National Institute of Statistics., n=unknown. | 30 days post transitions, 1990-2014 | Ecologic timeseries design, Quasi-Poisson regression, distributed lag nonlinear model. | DST-Onset: ↑ 30% fatal accidents.<br>DST-Offset: ↑ 16% fatal accidents. | L |

FARS=Fatality Analysis Reporting System, L=low, M=medium, H=high, *too many comparisons to allow coherent tabulated overview; see text for specifics.

##### *(ii) All Traffic Accidents*

Non-specific traffic accidents are reported in 12 studies, of whom five are conducted in the USA[36 37 40 45-47] and one Canada,[48] three in the UK,[44 45 49] and three in Finland,[50] Sweden,[51] and New Zealand.[52] The studies from Sweden and New Zealand are rated high quality.[51 52] The study from Finland[50] and one of the USA based studies are rated as low quality,[46] and the rest as middle quality.

***USA based (and Canada)***

The USA based studies include the studies by Huang et al. (2010) based in Minnesota,[36] Molina et al. (2023) based in Florida,[37] Stevens et al. (2006) based in Texas,[40] another study by Hicks et al. (1983) based in California,[47] and a study by Zhou et al. (2022) that is conducted in selected states in different time zones.[46] The study in Canada by Coren et al. (1996) is conducted in DST observing states only.

The study by Zhou et al. (2022) is rated low quality and reports decreased accidents for some days in some weeks (too many sporadic differences to reiterate in text here) following DST-Onset compared to what would be expected based on an extrapolation using 1-5 weeks data pre DST-Onset and data from weeks 14 and 15 post DST-Onset for the period 2014-2016. They also find an increase in accidents for some days and weeks following DST-Offset compared to what would be expected using a similar extrapolation method.

Of the medium quality studies, some decreases in accidents post DST-Onset are observed at particular times of day in Minnesota by Huang et al. (2010) on the Sunday and Monday and no change following DST-Offset compared to corresponding days in the reference weeks pre and post transitions for the period 2001-2007 (slightly different from fatal accidents wherein there was no DST-Onset difference).[36] In Florida, Molina et al. (2023) report no difference following DST-Onset for each day of the week and increases on Sunday and Monday only following DST-Offset compared to the previous week in the period 1983-2019 (very different to what is reported for fatal accidents in this study).[37] In Texas, Stevens et al. (2006) report no change in accidents (not including those that only damage property) following DST-Onset and a decrease following DST-Offset comparing the 5-day work week pre and post transition for the period 1998-2000 (slightly different from fatal accidents wherein there was no difference following DST-Offset).[40] In California, Hicks et al. (1983) only report numbers of crashes by day for the week pre and post transitions for the period 1976-1978, but we can analyse these numbers ourselves using paired t-tests.[47] We find (using their reported data) increased ($p<0.05$) accidents on the Sunday and Monday post DST-Onset and decreased ($p<0.05$) accidents on the Sunday only post DST-Offset and no differences when comparing the seven days combined. The study in Canada by Coren et al. (1996) reports increased and decreased accidents on the Monday post DST-Onset and post DST-Offset, respectively, compared to their corresponding preceding and following Monday for the period 1991-1992.[48]

Overall, the findings are quite variable, mostly limited to the Sunday and Monday following transitions, and not very similar to the corresponding study analyses of fatal accidents. Two of the six North American studies report increased accidents following DST-Onset; however, these were the oldest studies using data from 1991-92 and 1976-78.[47 48] More recent data suggests no change or decreased accidents. Following DST-Offset, two studies suggest increased accidents,[37 46] one of which is rated low quality.[46] Three other studies indicate decreased accidents and one indicates no change following DST-Offset.

**Non-USA (or Canada) based**

The studies by Lambe et al. (2000) in Sweden for the period 1984-1995 and by Robb et al. (2018) in New Zealand for the period 2005-2016 were rated as high quality.[51 52] In Sweden, Lambe et al. (2000) report no differences on the Monday post either transition compared to other Mondays.[51] In New Zealand, Robb et al. (2018) reported an increase in accidents on the Sunday and Monday post DST-Onset compared to corresponding days in the previous week and no change for other days.[52] For DST-Offset, they report decreased accidents on the Thursday but no change on other days.

The three medium quality studies from the UK also report mixed results. Bunnings et al. (2021) find no change following DST-Onset and increased accidents following DST-Offset for the period 1996-2017 (compared to decreases across both transitions for fatal accidents).[44] Singh et al. (2022) report decreased accidents following DST-Onset (for particular times-of-day and locations) and no changes following DST-Offset for the period 2005-2018 (compared to no changes when studying fatal accidents only).[45] Both studies by Bunnings et al. (2021) and Singh et al. (2022) used regression discontinuity design. Whittaker et al. (1996) report decreased accidents following DST-Onset and no changes following DST-Offset comparing the week pre and the week post transitions for the period 1983-1993 in the Cheshire region of the UK specifically.[49]

The low quality study from Finland by Lahti et al. (2010) reports no differences comparing the week pre to the week post transitions (pooled) in the period 1981-2006.[50]

Similar to the North American studies, only one of six non-North American studies report increased accidents following DST-Onset and one following DST-Offset, the rest report decreases or no change. Also, the findings here are quite variable and not consistent with the corresponding study analyses of fatal accidents. Thus, although increases in accidents have been detected following both transitions in some studies, increases appear less likely.

**Traffic-Table 2: Overview of n=12 studies of non-specific traffic accidents, in order by text subcategorization, quality rating, and chronology**

| Author (Year) | Population/Data | Exposure/Comparator | Statistics | Results | Rating |
|---|---|---|---|---|---|
| ***USA (and Canada) based*** | | | | | |
| Molina (2023)[37] | USA (Florida), Crash analysis reporting system, n=unknown | 1 week pre vs 1 week post transitions, 1983-2019 | Paired Wilcoxon rank test implemented using a Bayesian approach. | DST-Onset: no change.<br>DST-Offset: ↑ accidents on Sunday, Monday. | M |
| Huang (2010)[36] | USA (Minnesota), Minnesota Office of Traffic Safety, n=unknown. | 8 weeks pre vs post, first Sunday and first Monday post transitions vs all others over 16 weeks, 2001-2007 | Deviance and Pearson $\chi^2$ | DST-Onset: ↓ accidents at particular times of day on Sunday and Monday.<br>DST-Offset: no change. | M |
| Stevens (2006)[40] | USA (Texas), Fatal (K), incapacitating (A), and non-incapacitating (B) accidents (KAB), n=157,290 | 5 workdays pre vs 5 workdays post transition, 1998-2000 | Generalised linear models with an additional naïve before-and-after study. | DST-Onset: no change.<br>DST-Offset: ↓ accidents. | M |
| Coren (1996)[48] | Canada, Ministry of Transport. DST-Onset n=9593<br>DST-Offset n=12,010 | Monday post vs Mondays 1 week pre and 2 weeks post transitions, 1991-1992 | $\chi^2$ | DST-Onset: ↑ 8.6% accidents.<br>DST-Offset: ↓ 6.3% accidents. | M |
| Hicks (1983)[47] | USA (California), California Highway Patrol, n=100,111 | 1 week pre vs 1 week post transitions, 1976-1978 | Two-way ANOVA | DST-Onset: ↑ accidents on Sunday, Monday.<br>DST-Offset: ↓ accidents on Sunday | M |
| Zhou (2018)[46] | US data from Washington (pacific time), Idaho (mountain time), Illinois and South Dakota (central time), Virginia and Indiana (eastern time). | DST-Onset: 5 weeks pre vs 10 post transition. DST-Offset: 8 weeks pre vs 6 weeks post transition. Sunday and Monday 1 week pre vs post transitions, 2014-2016. | Regression analysis Modelling effects of time change vs perennial DST. | DST-Onset: ↓ accidents on Sunday (26.3%) and Monday (23.4%).*<br>DST-Offset: ↑ accidents on Sunday (22.1%) and Monday (8%). | L |
| ***Non-USA based*** | | | | | |
| Lambe (2000)[51] | Sweden, Swedish National Road Administration, n=6,844 DST-Onset: n=2650<br>DST-Offset: n=4,194 | First Monday pre and second Monday post vs first Mondays post transition, 1984-1995 | Incidence rate ratio from negative binomial regression. | DST-Onset: no change.<br>DST-Offset: no change. | H |
| Robb (2018)[52] | New Zealand, Accident Compensation Corporation, n=3795. | 1 week pre vs 1 week post transitions, 2005-2016 | Log-linear regression model. | DST-Onset: ↑ accidents on Sunday (16%) and Monday (12%).<br>DST-Offset: ↓ 9% on Tuesday. | H |
| Singh (2022)[45] | Great Britain, UK Department for Transport, n=311,766 | Optimal local bandwidth selector pre and post transitions, 2005-2018 | Regression discontinuity design analysis. | DST-Onset: ↓ accidents.<br>DST-Offset: no change. | M |
| Bunnings (2021)[44] | UK public road accidents, n>4.1 million | Mean squared error optimal bandwidth pre and post transitions, 1996-2017 | Regression discontinuity (RD) design, considering weather variables, time of day effects | DST-Onset: no change.<br>DST-Offset: ↑ accidents | M |
| Whittaker (1996)[49] | UK, STATS19, n=4185 | 1 week pre & 1 week transitions, 1983-1993 | $\chi^2$ | DST-Onset: ↓ 9.6% accidents.<br>DST-Offset: no change. | M |
| Lahti (2010)[50] | Finland, Finnish Motor Insurers' Centre, n=unknown | 1 week pre vs 1 week post transitions, 1981-2006 | Poisson model with log-link function | DST-Onset: no change.<br>DST-Offset: no change. | L |

L=low, M=medium, H=high. *too many comparisons to allow coherent tabulated overview; see text for specifics.

##### *(iii) Traffic Accident-Linked Hospital Admissions*

Two studies consider traffic accident-linked hospital admissions in Germany, Switzerland, and Austria for the period 2003-2017 (Nohel et al. 2021) and in Turkey (Teke et al. 2021) for the period 2014-2016, respectively.[53 54] The study by Nohl et al. (2021) is rated high quality while the study by Teke et al. (2021) is rated low due to poor comparison of pre vs post transition periods and being a single tertiary hospital study. Both observed increased admissions when comparing seven days post- to seven days pre DST-Onset and decreased admissions for DST-Offset, albeit the results by Teke et al. (2021) were not statistically significant. The low quality study (due to low sample size) of traffic-linked deaths by autopsies (Lindenberger et al. 2019) observed no differences across either transition in the Augsburg region of Germany.[15]

##### *Conclusion*

Considering all traffic accidents, some studies detect increases across either transition but most studies find no change or decreases. When zoning in on fatal accidents, more studies report increased fatal accidents following DST-Onset, but these are mostly based in the USA, with no changes reported in studies from the UK or Australia. More traffic linked hospital admissions following DST-Onset have been observed in two European studies. That some studies find increases in accidents, and fatal accidents in the USA in particular, means we cannot rule out an impact of transitions everywhere, but any effects of transitions on traffic accidents may not be as extreme expected. Effect sizes are also variable; however, some are strong enough for traffic accidents following transitions to warrant special attention (greater effect sizes are less easily explained by biases). The role of time-of-day of increases in accidents reported by some studies links daylight availability for visibility at particular times as a candidate causal factor.

**Traffic-Table 3: Overview of studies of traffic-linked hospital admissions and autopsies, in order by quality rating and chronology**

| Author (Year) | Population/Data | Exposure/Comparator | Statistics | Results | Rating |
|---|---|---|---|---|---|
| Nohl (2021)[53] | Germany, Switzerland, and Austria. n = 14,807 trauma patients | 1 week pre vs 1 week post transitions, 2003-2017 | $\chi^2$, Mann-Whitney U test. | DST-Onset: ↑ 17.4% admissions.<br>DST-Offset: ↓ 10.9% admissions. | H |
| Teke (2021)[54] | Turkey, hospital road traffic admissions, n = 710 | 1 week pre vs 1 week post transitions (2014-2016) and compared to perennial DST (2016-2018) | Kolmogorov-Smirnov test, $\chi^2$, Mann-Whitney U test. | DST-Onset: no change.<br>DST-Offset: no change. | L |
| Lindenberger (2019)[15] | Germany, Hospital autopsy data, n=86 traffic linked. | 2 weeks pre vs 2 weeks post transitions, 2006-2015 | $\chi^2$ | DST-Onset: no change.<br>DST-Offset: no change. | L |

L=low, M=medium, H=high. *too many comparisons to allow coherent tabulated overview; see text for specifics.

**Traffic-Table 4: JBI Quality Indicators**
**(Questions and possible answers are shown in detail in Table footnote)**

| Author (Year) | Q1 | Q2 | Q3 | Q4 | Q5 | Q6 | Rating | Notes |
|---|---|---|---|---|---|---|---|---|
| Bunnings (2021)[44] | Y | Y | N/A | N/A | Y | N | M | |
| Coren (1996)[48] | Y | Y | N/A | N/A | Y | N | M | |
| Fritz (2020)[34] | Y | Y | N/A | N/A | Y | N | H | |
| Goodwin (2023)[12] | Y | Y | N/A | N/A | U* | N | M | *Mentioned robustness checks, but referred appendix tables do not exist. |
| Hicks (1983)[47] | Y | Y | N/A | N/A | Y | N | M | |
| Hicks (1998)[35] | Y | Y | N/A | N/A | Y | N | L | |
| Huang (2010)[36] | Y | Y | N/A | N/A | Y | N | M | |
| James (2023)[42] | Y | Y | N/A | N/A | Y | N | H | |
| Lahti (2010)[50] | N~ | Y | N/A | N/A | U* | N | L | ~Unclear whether transitions were analysed separately or pooled. *No Stated sample size or exclusion criteria. |
| Lambe (2000)[51] | Y | Y | N/A | N/A | Y | N | H | |
| Lindenberger (2019)[15] | Y | Y | N/A | N/A | Y | N | L | |
| Molina (2023)[37] | Y | Y | N/A | N/A | Y | N | M | |
| Nohl (2021)[53] | Y | Y | N/A | N/A | Y | N | H | |
| Prats-Uribe (2018)[43] | Y | Y | N/A | N/A | Y | N | L | |
| Robb (2018)[52] | Y | N/A | N/A | N/A | Y | N | H | |
| Singh (2022)[45] | Y | Y | N/A | N/A | Y | U* | M | *No information on days used in analysis. |
| Smith (2016)[38] | Y | Y | N/A | N/A | Y | N | M | |
| Stevens (2006)[40] | Y | Y | N/A | N/A | Y | N | M | |
| Sood (2007)[39] | Y | Y | N/A | N/A | Y | U* | M | *Unclear what timeframe parameter is related to the presented data. |
| Teke (2021)[54] | N~ | Y | N/A | N/A | Y | N | L | ~Unclear whether statistical analysis is of transitions vs perennial DST or pre vs post transitions. |
| Varughese (2001)[41] | Y | Y | N/A | N/A | Y | N | M | |
| Whittaker (1996)[49] | Y | U* | N/A | N/A | Y | Y | M | *Inconsistency regarding age group casualty numbers for DST-Offset and some results in text do not align with their Table 2. |
| Zhou (2018)[46] | Y | U* | N/A | N/A | Y | N | L | *No standardised time frames for analyses. |

**JBI Critical Appraisal Checklist for Quasi-Experimental Studies (Adapted)**
Answers can be either Y=Yes, N=No, U=Unsure, or N/A.

1. Is it evident which factor is considered the 'cause' and which one is the 'effect'? Is there no ambiguity regarding the sequence of variables under investigation?
2. Were the characteristics of the study participants involved in comparisons adequately matched?
3. Were the exposures/treatments (beyond that of interest in our study) of the study participants involved in comparisons adequately matched?
4. Were the methods used to measure the outcomes the same for participants who were part of different comparisons?
5. Were appropriate statistical methods employed to analyse the data?
6. Are there any irregularities in the data reported?

### 4. Non-traffic Accidents, Injuries, & All-Cause Hospital Admissions Section Synthesis

Ursula Wild & Aiste Steponenaite

***Introduction***

In this section, 15 articles are subdivided into (i) occupation-related accidents and injuries (six studies), (ii) all cause or other non-traffic/occupational accidents/traumas (five studies, one overlaps with (i)), (iii) accidental deaths (three studies), and (iv) all-cause hospital admissions/visits (two studies). Of course, the latter subdivision can include more than accidents and injuries, but accidents and injuries are likely to strongly contribute to admission rates on a given day. All studies are registry-based, including, for instance, hospital registers, occupational accident registers, claims registers. Eight studies were conducted in the USA and Canada,[6 11 55-60] eight in Europe,[6 11 15 25 61-64] and one in New Zealand.[52] Several studies used data from multiple countries. All studies focus on the acute effects of transitions and one study considers potential DST effects across several weeks.

For study quality ratings, we defined high quality studies as those that included some form of robustness test, considered seasonality effects, controlled for holidays (where appropriate), and allowed answers indicative of higher quality to the JBI critical appraisal checklist questions more generally. Many medium quality-rated studies ticked most of the critical appraisal quality checklist boxes; however, they also included ambiguous comparisons, focused on self-reported data, or lacked demographic or accident-type specific information. Low quality-rated studies ticked fewer quality boxes or had more obvious and severe limitations. Specifics for each study are described in the following subsections.

Our synthesis includes the four subsections described above, a Table 1 (Accident-Table 1) in which we provide an overview of studies by subsections, and a Table 2 (Accident-Table 2) of the JBI quality indicators. The section closes with an overall conclusion.

***(i) Occupation-related accidents and injuries***

Four studies indicate no transition effects on workplace accidents;[52 57 58 63] two studies indicate an effect of DST-Onset, but in opposite directions.[56 59] All studies were rated medium quality except for the studies by Robb (2018) and Kolla et al. (2021) which were rated as high and low quality, respectively.[58] Kolla et al. (2021) indicate no change in safety-related incidents concerning patients that were reported by healthcare staff at a large healthcare organisation across the USA (2010-2017) post DST-Onset and DST-Offset. Of course, safety-related incidents are not necessarily specific to factual accidents that caused injury or harm, rather they also include those that could cause harm. They can include, for instance, near misses or something that could lead to an accident at a later date (e.g., the identification of labels that look similar for different medications being placed close to each other).

Regarding the other three studies reporting no effect of transitions, Lahti et al. (2011) compared occupational accidents in the week pre and post transitions (note, DST-Onset and DST-Offset were not analysed separately).[63] The analysis was performed on all compensated accidents recorded in the Federation of Accident Insurance Institutions' registry for Finland in the years 2003-2006. Comparing the occupational accidents one week post transitions with one week pre (a total of 14,151 accidents), there were no statistically significant associations. Holland et al. (2000) report no significant difference between the first week pre and post transitions using 1990-1996 data from the construction accident database of the State of Washington.[57] There were also no differences between the first week pre transition and the second week post transition.

In the one high quality study, Robb et al. (2018) also found no significant difference in work-related accidents comparing one week pre and post DST-Onset or DST-Offset using data from an accident compensation corporation in New Zealand (2005-2016).[52]

Regarding the two studies reporting a transition effect, Barnes et al. (2009) report a 3.6 fold increase in the number of mining injuries on the Monday following DST-Onset compared to all other days of the year.[56] No significant effect was observed for DST-Offset. Injuries were reported to- and recorded on- the USA National Institute for Occupational Safety and Health's register for occupational accidents in the USA (1983-2006). Holidays, day of week, and week of year were included in the statistical models; however, Barnes et al. (2009) describe coding the day of transitions in their respective models as "1" and all other days as "0". Morassaei et al. (2010) analysed work injury claims registered to a Workplace Safety & Injury Board in Ontario, Canada (1993-2007) and observed a decrease on the Thursday-Saturday in the first week post DST-Onset compared to the week pre.[59] However, this decrease is possibly due to years when Good Friday occurred in the week post DST-Onset because the effect disappeared when excluding these years. They also report no difference following DST-Offset. This study was rated medium as claims would not necessarily include all workplace accidents, only those sufficiently severe to make a claim.

Overall, there is little-to-no evidence to suggest a remarkable effect of DST-Onset increasing workplace accidents.

##### *(ii) All-cause or other non-traffic/occupational accidents/traumas*

One out of five studies indicates transition effects.[6 25 52 61 64] In this study, Zhang et al. (2020), examined a multitude of health outcomes in the USA (IBM Watson Health MarketScan dataset; 2003-2014) and in Sweden (Swedish national inpatient register from 1986-2011).[6] In the USA, they found an increase in injuries post DST-Onset when comparing the day of the week post transition with the average of the corresponding days two weeks pre and two weeks post DST-Onset. No differences were observed in Sweden.

Of the four studies indicating no differences, Lahti et al. (2008) indicates no effect when investigating changes in hospital-treated accidents following DST-Onset and DST-Offset transitions (analysed separately) in Finland (Finnish Hospital Discharge Register; 1987-2003).[25] However, it is unclear whether the start point or end point of hospital treatment was utilised in statistical models; the authors indicate determining both variables and, of course, the latter might not reflect a transition related effect as the accident could have occurred pre transition. Nohl et al. (2021) observed no differences in prevalence of penetrating or blunt traumas when comparing the week pre with the week post transitions (DST-Onset and DST-Offset not analysed separately) using data from the German Trauma Surgery Register (DGU; 90% from Germany, 10% from Austria and Switzerland; 2003-2017).[64] Post transitions severity scores and days in intensive care were slightly higher than pre transitions (22.3 vs 21.8 severity score, 3 vs 2 days, respectively). There was, however, a difference between mean injury severity score from pre to post DST ($p=0.052$) and the number of ICU days increased ($p=0.035$), assessed by $\chi^2$ analyses. Counts for differences between pre and post transitions for both DST-Onset and DST-Offset are provided, but they are pooled years and without a descriptor of variability; thus, we cannot test for possible differences by DST-Onset and by DST-Offset separately. Ponkilainen et al. (2022) observed no significant differences in incidence of femur fractures in older people ($\geq$ 70 years) in Finland using data from the Finnish National Hospital Discharge Register (1997-2020).[61] They compared the Mondays post transitions to all other days of the year. Robb et al. (2018) found no significant change in falls or home and community accidents comparing one week pre and post DST-Onset or DST-Offset using data from an accident compensation corporation in New Zealand (2005-2016).[52]

Overall, there appear to be no clear effects of transitions on all-cause or non-traffic/occupational accidents and traumas.

##### *(iii) Accidental deaths*

This subdivision concerns three articles; two of whom find increased accidental deaths in the first week following DST-Onset compared to the previous week but no difference around DST-Offset.[15 55] Specifically, Coren (1996) finds 6.5% more deaths in the first four days of the week post DST-Onset compared to both the corresponding days in the previous week and the following week in the USA National Centre for Health Statistics registry (1986-1988).[55] This includes traffic and occupation-related accidents. Lindenberger et al. (2019) find more "non-natural" deaths compared to the previous two weeks and the following week using a $\chi^2$ analysis of forensic autopsy data from Frankfurt/Main, Germany (2006 to 2015).[15] This category includes traffic-, drug-, fire-, malpractice, occupation-, murder-, and other accident-related deaths. There was no change in occupation- or other accident-related deaths, but the counts of these events are very few. Lee et al. (2019) find no changes in the median number of trauma activations based on data from a trauma registry in a tertiary care hospital in Pennsylvania, USA ("over 20 years") when comparing the Mondays one week pre, first week post, and second week post DST-Onset and DST-Offset, using Mann-Whitney U tests.[60] Overall, increased accidental deaths are observed following DST-Onset, but to what extent this is driven by non-traffic accidents is unclear.

##### *(iv) All-cause hospital admissions/visits*

Two studies assess all-cause hospital admissions. Ferrazzi et al. (2018) and Jin et al. (2020) find decreased visits to the Accident & Emergency department of a hospital in Italy (2007-2016) and decreased admissions to hospitals in general in Germany (2000-2008) in the week following DST-Offset compared to the previous week(s), respectively (or at least for the first four days of week in the latter study).[11 62] Jin et al. (2020) finds little change comparing pre to post DST-Onset, but Ferrazzi et al. (2018) reports increased Accident & Emergency visits. However, the changes observed in the study by Ferrazzi et al. (2018) may not be due to transition.

More specifically, using Poisson regression, Ferrazzi et al. (2018) analysed two weeks pre to 19 weeks post DST-Onset, and two weeks pre to five weeks post DST-Offset.[62] Total accident and emergency admissions increased from ~ weeks 4-12 post DST-Onset and decreased in week 19 post DST-Onset compared to the week pre DST-Onset. The increase remains through to week 19 if only minor severity admissions are assessed. Decreased visits appeared in the two weeks pre DST-Offset and in weeks 1-4 post DST-Offset. After adjusting for photoperiod, however, few significant differences remained (from many assessed) following DST-Onset and DST-Offset. When assessing return visits (which would typically be scheduled by a doctor as follow-up), the initial differences were the same as total visits but adjustment for photoperiod did not affect estimates. Reasons why significant effects would be observed for one stratum of the total visits (i.e., return visits) but not for total visits overall is unclear. Rationale for assessing the stratum of return visits only is missing. Overall, and especially as visits appear to decrease again pre DST-

Offset, it is difficult to infer that transitions or living with DST compared to Standard Time during summer months is causal of increased visits.

Jin et al. (2020), assessing the effect of DST-Offset, find decreased total admissions and decreased admissions for various causes, including, the injury admission rate.[62] Various robustness tests included estimating 14 and 21 post DST-Offset day effects instead of just 7, and accounting for weather. Following DST-Onset, they find significant increased injury admissions on the Tuesday post DST-Onset compared to the average.

Overall, it is unclear whether all-cause hospital admissions are increased and decreased following DST-Onset and DST-Offset, respectively; however, the latter, especially for injuries, seems more likely.

##### *Conclusion*

There is little-to-no evidence to suggest a remarkable effect of transitions affecting non-traffic accidents & injuries. Increased accidental deaths are observed following DST-Onset, but to what extent this is driven by non-traffic accidents is unclear. It is also unclear whether all-cause hospital admissions are increased or decreased following DST-Onset and DST-Offset, respectively; however, the latter, especially for injuries, seems more likely. From these 15 registry-based studies of (i) occupation-related accidents and injuries, (ii) all-cause or non-traffic/occupational accidents and traumas, (iii) accidental deaths, and (iv) all-cause hospital admissions, it appears unlikely that transitions increase risk of non-traffic injury.

**Accident-Table 1: Overview of studies, in order by text subcategorization, quality rating, and chronology**

| Author (Year) | Population/Data | Index Period | Reference Period | Outcome/Results | Statistics | Rating |
|---|---|---|---|---|---|---|
| ***Workplace Accidents*** | | | | | | |
| Robb (2018)[52] | New Zealand, n=12.6 million total claims (only work-related claims used) | First week post transitions, 2005-2016 | First week pre transitions, 2005-2016 | No changes. | Log-linear regression. | H |
| Lahti (2011)[63] | Finland, n=14,151 accidents | First week post transitions, 2003-2006 | First week pre transitions, 2003-2006 | No changes (transitions not analysed separately). | Poisson regression. | M |
| Morassaei (2010)[59] | Ontario-Canada, Workplace Safety & Injury DST-Onset: n=199,846 claims. DST-Offset: n=269,100 claims. | First week post transitions, 1993-2007 | First week pre and 2nd week post transitions, 1993-2007 | DST-Onset: ↓ claims on Thursday-Sunday DST-Offset: no changes. | Poisson regression. | M |
| Barnes (2009)[56] | USA, n=576,292 mining injuries, 98% male, mean age=39 years, mean work experience=6.5 years | Monday post transitions, 1983-2006 | All other days, 1983-2006 | DST-Onset: ↑ mining injuries (database) and missed work days. DST-Offset: no changes. | Hierarchical linear model | M |
| Holland (2000)[57] | Washington-USA, n=unknown | First week post transitions, 1990-1996 | First week pre and 2nd week post transitions, 1990-1996 | No changes in injuries reported to construction accident database. | Paired two-sample t-test. | M |
| Kolla (2021)[58] | "Multiple states", USA | First week post transitions, 2010-2017 | First week pre transitions, 2010-2017 | No changes in self-reported Patient Safety-Related Incidents. | Negative binominal mixed model | L |
| ***All-cause or Other non-traffic/occupational Accidents /Traumas*** | | | | | | |
| Ponkilainen (2022)[61] | Finland, National Hospital Discharge Register, n=112,658 femur fractures | Monday post transitions, 1997-2000 | All other days, 1997-2000 | No changes. | Negative binomial regression. | H |
| Zhang (2020)[6] | USA MarketScan (n>150 million), Swedish national inpatient register (n>9 million) | First weekdays post transitions, 2003-2014 | Corresponding days one week pre and 2nd week post transitions, 2003-2014 | DST-Onset: ↑ injuries in USA but not Sweden. | Bayesian and frequentist relative risks, placebo controls. | H |
| Nohl (2021)[64] | Trauma Register DGU: Germany (90%), Austria, Switzerland, n=14,807 patients, males=71%, mean age=51±22 years | First week post transitions, 2003-2017 | First week pre transitions, 2003-2017 | No changes. (transitions not analysed separately) | $\chi^2$ or Mann–Whitney U-test. | M |
| Lahti (2008)[25] | Finland, data from the Finnish Hospital Discharge Register | 2 weeks post transitions, 1987-2003 | 2 weeks pre transitions, 1987-2003 | No changes. | Poisson Regression | M |
| ***Hospital Admissions*** | | | | | | |
| Jin (2020)[11] | Germany, n=336,604, from national census; 54.28% females, age 0-2=1.8%, age 65-74 years=1.6%, >74 years=0.04% | First week post DST-Offset, 2000-2008 | 3 weeks pre and 2nd week post DST-Offset, 2000-2008 | DST-Onset: no changes. DST-Offset: ↓ admissions. | Regression model | H |
| Ferrazzi (2018)[62] | Italy, n=366,527 A&E visits to a hospital, mean age ~ 52 years | Period 1=9 weeks post and 2 weeks pre DST-Onset. Period 2=4 weeks post and 2 weeks pre DST-Offset Period 3=5 weeks from 2nd week Jan. 2007-2016 | | DST-Onset: ↑ on different weeks. DST-Onset: ↓ on different weeks. Possible seasonal effect. | Poisson models | M |
| ***Accidental Deaths*** | | | | | | |
| Lindenberger (2019)[15] | Frankfurt/Main, Germany, n = 690 autopsy cases (460 males, 230 = females) | 2 weeks post transitions | 2 weeks pre transitions | DST-Onset: ↑ autopsy cases. DST-Offset: no changes. | $\chi^2$ | M |
| Coren (1996)[55] | USA, DST-Onset: n=8,429, DST-Offset: n=8,771 | First week post transitions, 1986-1988 | First week pre and 2nd week post transition, 1986-1988 | DST-Onset: ↑ accidental deaths. DST-Offset: no changes. | $\chi^2$ | M |
| Lee (2019)[60] | Pennsylvania, USA, n = 2-3 | Monday post transitions, 20 years | Monday pre transitions | No changes (transitions not analysed separately) | Mann-Whitney U-test | L |

L=low, M=medium, H=high.

**Accident-Table 2: JBI quality indicators**
(Questions and possible answers are shown in detail in Table footnote)

| Author (Year) | Q1 | Q2 | Q3 | Q4 | Q5 | Q6 | Rating | Notes |
|---|---|---|---|---|---|---|---|---|
| Barnes (2009)[56] | Y-NA | Y | U-N/A | Y-N/A | Y | N | M | -Discrepancies in text regarding what day they analysed, only self-reported accidents were recorded, years with a certain job title is not the same as years of experience *per se*. |
| Coren (1996)[55] | Y | Y | N-N/A | U-N/A | Y | N | M | -Methods part is brief and give no detailed information. |
| Ferrazzi (2018)[62] | Y | Y | U-N/A | Y-N/A | Y | N | M | -Limited data on distribution of visits across assessment periods. |
| Holland (2000)[57] | Y | Y | U-N/A | Y-N/A | Y | Y-N | L | -Limited in location (Washington) and outcome (construction accidents), weather was not considered, only reported accidents considered, no description of demographical data, no information about kind or severity of the injuries. |
| Jin (2020)[11] | Y | Y | U-N/A | Y-N/A | Y | Y-N | H | -Few inconsistencies between text and tables. |
| Kolla (2021)[58] | Y | Y | U-N/A | Y-N/A | Y | N | M | -Only self-reported incidents were considered, study population across the various states of the USA without mentioning possible influence of longitudes. |
| Lahti (2011)[63] | Y | Y | U-N/A | U-N/A | Y | N | M | -Unclear whether hospital admission and/or discharge data were used, transitions analysed together. |
| Lahti (2008)[25] | Y | Y | U-N/A | U-N/A | Y-U | N | M | -Minor discrepancies in the text. |
| Lindenberger (2019)[15] | Y | Y | U-N/A | U-N/A | Y | N | M | -Low sample size. |
| Lee (2019) [60] | Y | Y | N/A | Y | U | N | L | -Low sample size. |
| Morassaei (2010)[59] | Y | Y | U-N/A | U-N/A | Y | N | M | -Only severe injuries likely to be included. |
| Nohl (2021) [64] | Y | Y | U-N/A | U-N/A | Y | N | M | -Minor discrepancies in the text, transitions analysed together. |
| Ponkilainen (2022) [61] | Y | Y | U-N/A | U-N/A | Y | N | H | |
| Robb (2018)[52] | Y | Y | U-N/A | U-N/A | Y | N | H | |
| Zhang (2020)[6] | Y | U-Y | U-N/A | Y-N/A | Y | N | M-H | -Risk of "driver" and "passenger" diseases, risk of coding errors, only insured individuals included. |

**JBI Critical Appraisal Checklist for Quasi-Experimental Studies (Adapted)**
Answers can be either Y=Yes, N=No, U=Unsure, or N/A.

1. Is it evident which factor is considered the 'cause' and which one is the 'effect'? Is there no ambiguity regarding the sequence of variables under investigation?
2. Were the characteristics of the study participants involved in comparisons adequately matched?
3. Were the exposures/treatments (beyond that of interest in our study) of the study participants involved in comparisons adequately matched?
4. Were the methods used to measure the outcomes the same for participants who were part of different comparisons?
5. Were appropriate statistical methods employed to analyse the data?
6. Are there any irregularities in the data reported?

#### 5. Sleep & Circadian Section Synthesis

Philip Lewis & Sally Ferguson

##### *Introduction*

Systematic screening returned 33 articles that consider measures of sleep and circadian biology. The vast majority of studies have been conducted in Europe and in the USA. Study populations vary from children and adolescents to university students, adults, and those with illnesses. Major limitations in studies of transition effects include low samples sizes in studies that use repeated measures while those with larger sample sizes often use different populations in reference and index periods. Index periods vary from the transition night only to several days post transition. The usage of actimetry devices and diaries/questionnaires to assess sleep is well mixed. Most studies assess sleep duration and some markers of sleep timing (e.g., sleep onset or offset or mid-point); fewer assess sleep quality and sleepiness.

This section is divided into the following subsections: (i) transition effects on sleep (19 studies), (ii) transition effects on circadian biology (six studies), (iii) DST vs Standard Time (and longitude) effects on sleep (studies), and (iv) DST vs Standard Time (and longitude) effects on circadian biology (six studies). Some studies are included in more than one subsection. Sleep onset or offset or mid-point is considered in the sleep-specific subsections as opposed to the circadian-specific subsections even though it is sometimes used as a marker of circadian phase. Designated circadian-related measures include cosinar activity parameters, wrist temperature rhythms, cortisol rhythms, social jet lag, and chronotype. For each of the subsections 1-4, a tabulated overview of the included studies and their final quality ratings is provided in Sleep-Tables 1-4, respectively. Given the larger volume of studies in the subsection on transition effects on sleep, a summary of conclusions pertaining to DST-Onset and to DST-Offset is provided in Box 1 and Box 2 below these syntheses, respectively. JBI quality indicators are presented in Sleep-Table 5 at the end of this section. A conclusion paragraph closes this section.

Lastly, one of the 33 articles is a reanalysis of six other studies and will only be discussed further in the concluding paragraph at the end of this section.[65] Another study suffers from severe limitations such that it does not warrant inclusion in synthesis or discussion.[66] In effect, 31 articles are synthesized in this section.

##### *(i) Acute transition effects on sleep*

First, we synthesize (a) DST-Onset associated findings by three high quality studies, then by one medium quality study, and then by nine low quality studies. Box 1 provides an overview of conclusions pertaining to DST-Onset effects on sleep. Second, we synthesize (b) DST-Offset findings by two high quality studies, by one medium quality study, and then by nine low quality studies. Box 2 provides an overview of conclusions pertaining to DST-Offset effects on sleep. An overview of studies in this subsection of acute transition effects on sleep is provided in Sleep-Table 1.

###### *(a) DST-Onset*

###### *High Quality*

Heacock et al. (2022) report a decrease in sleep duration, delayed sleep onset and minimal change in sleep offset timing, and no change in sleep consistency in 24,250 users of the *Whoop* actimetry device and online platform (specifically, those with +70% wear time across one year).[67]

Medina et al. (2015) report a decrease in sleep duration and quality, an increase in sleepiness, and no change in awakenings or latency in 35 high school students.[68] Fetter et al. (2014) report no change in sleepiness in 83 non-dementia Parkinson's patients.[24] These studies score high because of repeated measures.

Heacock et al. (2022) assessed sleep parameter differences between the transition night only and the mean of the same day of week for the 4 weeks pre and the 4 weeks post transition night, which is a weakness because acute effects (if any) may be expected to extend beyond the transition night.[67] From the absolute value plots provided in the article, sleep duration is 7-7.2 hours on DST-Onset night but many other calendar days are similar. Sleep consistency is comparatively low. While sleep onset and offset timing are not unusual *per se*, a discontinuity in trend around transition can be observed (times are given as local time). In terms of relative change compared to the reference period, sleep duration was lower by ~1% and sleep onset and offset delayed by ~15-20mins in terms of local. Of course, the delay in offset in regard to local time represents an advance of ~40-45min in regard to when the body is expecting sleep assuming the reference sleep reflects the anticipated timing. Overall, though, these do not appear extreme, especially compared to many other days of the year in the provided plots. Additional limitations to this study include the non-generalizable sample population (e.g., those with heightened health interests as users of *Whoop*) and the non-research grade activity device used to assess sleep.

Medina et al. (2015) compared diary and actigraphy ("Micro-Mini Motionlogger Actigraph") assessed sleep parameters on the Monday to Friday pre and post DST-Onset for 35 high school students whose school day started at 07:40.[68] Diary- and actigraphy-assessed sleep duration were shorter, especially between Fridays. More generally, sleep quality was lower and sleepiness was higher. No change in latency, efficiency, awakenings, or 'wake after sleep onset' between index and reference periods were observed, but fewer awakenings on the Monday post transition were observed. The authors indicate no difference when stratified by chronotype in text but results are not shown.

Fetter et al. (2014) found no change in sleepiness across transitions among non-dementia Parkinson's disease patients seen at their clinic in the UK. Sleepiness was assessed on three consecutive days in the week pre and the week post transition (specific days not mentioned).

***Medium Quality***

The study by Kantermann et al. (2007) was rated as medium quality because there is no descriptive comparison of the individuals assessed pre and post transition and the study concerned only one of each DST-Onset and DST-Offset transition.[69] In the first part of their study, and albeit difficult to comment on acute effects of transition because the data presented are half-monthly averages, the authors observe an apparent beginning of a plateau in mid-sleep and sleep offset from DST-Onset based on single time-point data from ~55,000 participants from central Europe in the MCTQ database. Important to note is that this a plateau is described in terms of solar time and not local clock time. Thus, sleep offset after DST-Onset would be delayed based on local clock time. The significant cosine fit across seasonal sleep duration would suggest no direct impact of DST-Onset on sleep duration (albeit the forced fit would not show a discontinuity) but the half monthly average change across DST-Onset could indicate an increase in sleep duration (the latter is speculative given the resolution of the data). There is no apparent plateau line in sleep duration at transition as it appears to continue decreasing until mid-summer. In part 2 of their study, the authors identify advanced mid-sleep timing post DST-Onset in 50 volunteers assessed over 4 weeks. There are no differences among pre or post transition weeks, only between pre and post transition weeks, which suggests either sleep phase does not track sun time or mid-sleep timing at least remains the same because of changes in sleep duration in this population. At a glance, this may be contradictory to what is indicated in the first part of their study, but this could be an effect of time resolution of measurements in the first part of their study.

***Low Quality***

Nine studies were rated as low quality because of very low sample sizes, non-repeated measures, not strictly targeting DST-Onset as the intervention, or irregularities in the data. For brevity, first author and year are not mentioned in text (unlike above). Seven of these studies consider sleep duration. In three studies there is no evidence of a change in sleep duration following DST-Onset, including in eight healthy adults in Spain 2017-2018,[70] in 19,772 respondents in the US Behavioural Risk Factor Surveillance System (BRFSS) from 2001-10,[71] or in 136 hospitalised patients with a variety of medical conditions in Italy in 2013.[72] Decreased sleep duration was observed in three studies, including in a study of 554 medical interns in the USA (but the decrease was small [~12mins] and only in those with a polygenetic risk score that indicated being a later chronotype),[73] in a study of low sample size (n=10) healthy persons in Finland (up to 60mins),[74] and in the American Time Use Survey (ATUS) but only on transition night for 45mins.[11] Only one study observed increased sleep duration but in a low sample size (n=14 ) of university students in Italy.[75] In the studies that indicate a decrease in sleep duration, the finding is not generalisable – affecting only 1/3 of a health-orientated population in the USA by ~12mins and affecting ten individuals in Finland for whom we have no descriptive data. Overall, a change in sleep duration cannot be concluded from these studies.

Regarding timing, naturally there is (or, at least, would be) a change indicated for those in whom sleep duration was observed to change.[11 73-75] No timing differences were observed in three studies but these studies may be influenced by timing of Easter holidays and hospital schedules for patients and workers.[70 72 73] Advanced sleep onset and offset was reported in one further study and advanced sleep offset in two further studies.[75-77] However, in the study with advances in both sleep onset and offset, the sleep offset time occurs after the time of attending university morning classes, which was one of the sample selection criteria. Nonetheless, it appears more likely that sleep onset and/or offset will change following DST-Onset, at least for some people.

No differences were observed in sleep quality, daytime sleepiness, latency, or awakenings in the study of hospital patitents.[72] Sleepiness was higher at 08:00 but not at other times of day in the study of university students attending morning classes.[77] Increased awakenings and decreased sleep efficiency (sleep duration/time in bed) was observed in another study of university students, likely linked to the reported increased time in bed.[75] Daytime sleepiness was higher post DST-Onset in a study of adolescent school pupils in Germany but different populations were assessed pre and post DST-Onset.[78] Lastly, in a study of nine Finnish participants, no change in sleep efficiency but increased sleep movement and fragmentation post DST-Onset is reported, but there is no mention of statistics tests used. The authors also state that they collected more sleep data, but do not provide data or any indication that these are analysed.[79] There is insufficient evidence to draw conclusions about sleep quality or other sleep parameters from these studies.

**Box 1: Summary Conclusions**
**DST-Onset transition effects on sleep**

**High quality studies**[24 67 68]

- Decrease in sleep duration on transition night (but not lower than many other days), possibly prolonged (i.e., beyond transition night) decreased duration in adolescent school students whose school day started at 07:40.
- Advanced sleep onset and offset (relative to reference sleep phase; albeit appearing as delayed on local clock times) seems likely, at least for a few days.
- Possible decreased sleep quality and increased sleepiness for adolescent school students, at least for a few days.

**Medium quality study**[69]

- No apparent impact on sleep duration.
- Mixed findings on mid-sleep timing and no clear effect on wake timing.

**Low quality studies**[56 70-79]

- A change in sleep duration cannot be concluded.
- It appears more likely that sleep timing will advance following DST-Onset, at least for some people (advanced = relative to reference sleep phase; albeit appearing as delayed on local clock times).
- Insufficient evidence to draw conclusions about sleep quality or other sleep parameters.

**Overall**

- Sleep duration is likely impaired for adolescent school students with an early school start time in the week following DST-Onset. This is based on a study of 35 students. There is no strong evidence to suggest that sleep duration is generally impaired in the week post DST-Onset.
- Generally, sleep onset and offset is likely to be advanced (relative to reference sleep phase; albeit appearing as delayed on local clock times).
- There is insufficient evidence to draw conclusions about sleep quality or other sleep parameters.

##### *(b) DST-Offset*

###### *High Quality*

Two studies already described above are included here. Heacock et al. (2022) report increased duration (~25mins) and consistency and delayed sleep onset (~15mins) and offset (~40mins) on DST-Offset night only in their study of *Whoop* device users.[67] Fetter et al. (2014) report no change in sleepiness in their study of Parkinson's patients.[24]

###### *Medium Quality*

Also described above, the study by Kantermann et al. (2007) indicates mid-sleep and wake times appear to delay (according to solar time) and there is no apparent impact on sleep duration in part 1 of their study.[69] In part 2 of their study, mid-sleep time appears to delay around transition but no clear effects are observed thereafter.

###### *Low Quality*

In the low quality-rated studies, three studies report increased sleep duration.[11 70 80] Of these, the study by Harrison et al. (2013) suggests a possible role for short and long habitual sleeping durations with the former sleeping longer and the latter sleeping shorter and no change in the habitual medium duration sleepers in the few days post DST-Offset when assessed by diary.[80] However, no duration difference was observed in a subset also assessed by actigraphy. Moreover, in addition to no repeated measures used in the analysis, the plots of mean difference in sleep duration add to ambiguity as they show differences for Sunday, while only one Sunday night was described as assessed (i.e., no comparator or reference Sunday). Regression to the mean also presents as an issue for the differences observed by habitual sleepers. The study by Jin et al. (2020) using US BRFSS data from 2013-16 that finds increased sleep duration post DST-Offset is in contrast to their null finding using data from 2001-10 published in their working paper.[11 71] Another study reports no change, another reports a decrease, and another reports individual differences (but no pre vs post group comparisons in the latter study of four participants) in sleep duration.[56 75 81] Taken together, a change in sleep duration cannot be concluded, but an increase in duration appears more likely than a decrease. Delayed sleep offset time is reported by five studies.[70 75 76 80 82] Individual differences in the study of four participants are reported.[81] No change in sleep onset is reported in two studies.[70 75] A delay in sleep offset appears likely, which would support increased sleep duration. No change in latency or fragmentations is reported in one study.[75]

**Box 2: Summary Conclusions**
**DST-Offset transition effects on sleep**

**High quality studies**[24 67]

- Increase in sleep duration on transition night.
- Delayed sleep onset and/or offset.

**Medium quality study**[69]

- No apparent impact on sleep duration.
- Delayed mid-sleep timing.

**Low quality studies**[56 70-79]

- A change in sleep duration cannot be concluded.
- It appears more likely than not that sleep offset will delay following DST-Offset, at least for some people.
- Insufficient evidence to draw conclusions about sleep quality or other sleep parameters.

**Overall**

- Sleep duration is likely increased in the week post DST-Offset.
- Sleep offset is likely to be delayed in the week post DST-Offset.
- Insufficient evidence to draw conclusions about sleep quality or other sleep parameters.

**Sleep-Table 1: Acute transition effects on sleep, in order by quality and chronology**

| Author (Year) | Population/Data | Index Period/ Intervention | Reference Period/ Comparator | Outcome ("↑"=increase/delay; "↓"=decrease/advance, "–" =no change) DST-Offset | DST-Onset | Statistics | Rating |
|---|---|---|---|---|---|---|---|
| Heacock (2022)[67] | N=24,250 (25.5% female), 18-65 years, Whoop platform and device users reporting ≥70% days of wear time across one year, USA | Transition nights, 2020-21 | Mean of same nights on 4 weeks pre and post transitions, 2020-21 | ↑ duration, sleep onset, sleep offset, consistency | ↓ duration, ↓ sleep onset, sleep offset, - consistency | Mean differences with 95% confidence intervals. | H |
| Medina (2015)[68] | N=35 high school students (20 female), 15-18 years, school day start at 07:40, USA | Mon-Fri post onset, presume 2015 | Mon-Fri pre onset, presume 2015 | n/a | ↓ duration (especially between Fridays), quality, ↓ awakenings (between Mondays) ↑ sleepiness - latency, efficiency | Paired t-test with Holm-sequential-Bonferroni adjustment | H |
| Fetter (2014)[24] | N=83 non-dementia Parkinson's Disease patients (31 female), 67±7.7(SD) years, UK | 3 days in week post transitions, 2011-12 (1 onset & 2 offset periods) | 3 days in week pre transitions, 2011-12 (1 onset & 2 offset periods) | - sleepiness | - sleepiness | Student's t test | H |
| Kantermann (2007)[69] | MCTQ database, central Europe & N=50 (34 female), 18-59 years, central Europe | 4 weeks post transitions, 2006-07 | 4 weeks pre transitions, 2006-07 | ↑ mid-sleep times, regardless of chronotype. Pre weeks significantly different from post weeks but not among each other either side of transition. | ↓ mid-sleep times, regardless of chronotype. Pre weeks significantly different from post weeks but not among each other either side of transition. | RM-ANOVA | M |
| Arguelles-Prieto (2022)[70] | N=8 (4 female), 33±11(SD) years, Spain | 1 week post transitions, 2017-18 | 4 days pre transitions, 2017-18 | ↑ duration, sleep offset - sleep onset | - duration, sleep onset, sleep offset | Student's t test & RM-ANOVA | L |
| Oskardottir (2022)[81] | N=4, 52-86 years, Denmark | 15, 30, and 60 days post offset, 2017 | 15, 30, and 60 days pre offset, 2017 | Individual differences in duration and timing. | n/a | T-test of individual means/counts | L |
| Tyler (2021)[73] | N=554 medical interns, USA | 1 week post onset, 2019 | 1 week pre onset, 2019 | n/a | ↓ duration | T-test | L |
| Jin (2020)[11] | N=174,503 survey respondents (BRFSS), 18+ years, USA | 1 week post offset, 2013-16 | 3 weeks pre and 2nd and 3rd week post offset, 2013-16 | ↑ duration | n/a | Multivariable linear regression. | L |
| Jin (2015)[71] | N=19,772 survey respondents (BRFSS), 18+ years, USA | 1 week post transitions, 2001-10 | 3 weeks pre and 2nd and 3rd week post transitions, 2001-10 | - duration ↑ "unintentionally following asleep in last 30 days" post offset. | -duration | Multivariable linear regression. | L |
| Bano (2014)[72] | N=136 hospitalised patients (81 female), 77±11(SD) years, Italy | 01-21 April, 2013 | 01-30 March, 2013 | n/a | - duration, sleep onset, sleep offset, latency, quality, awakenings, sleepiness | RM-ANOVA | L |

**Sleep-Table 1: Acute transition effects on sleep, in order by quality and chronology**

| Author (Year) | Population/Data | Index Period/ Intervention | Reference Period/ Comparator | Outcome ("↑"=increase/delay; "↓"=decrease/advance, "–" =no change) DST-Offset | DST-Onset | Statistics | Rating |
|---|---|---|---|---|---|---|---|
| Quintilham (2014)[77] | N=318 (281 female) university students, ~ 20 years, Brazil | Mon-Fri post onset, 2008-11 | Mon-Fri pre onset, 2008-11 | n/a | ↓ sleep onset, sleep offset<br>↑ sleepiness at 08:00 but not other times. Possible chronotype differences.<br>-duration | T-test and two-way ANOVA. | L |
| Tonetti (2013)[75] | N=14 university students (all male), 21-30 years, Italy | Mon-Fri pre transitions, 2009-10 | Mon-Fri post transitions, 2009-10 | ↓ duration, time in bed<br>- sleep onset, latency, fragmentations<br>↑ "get-up" time | ↑ duration, time in bed, efficiency, awakenings<br>- sleep onset, latency, fragmentations<br>↓ "get-up" time, quality | T-test | L |
| Harrison (2013)[80] | N=120 students (~68% female), 23.1±8(SD) years, UK | Mon-Sat post offset, 2010 | Mon-Sat pre offset, 2010 | ↑ duration & offset in short sleepers, opposite in long sleepers (pre average sleep). No differences by actigraphy. | | Two-way ANOVA, Three-way ANOVA | L |
| Barnes (2009)[56] | N=14,310 ATUS participants who were employed (50.8% female), 42.3 mean years, USA | Transition nights, 2004-06 | Non-transition nights, 2004-06 | - duration | ↓ duration (~40mins) | HLM | L |
| Schneider (2009)[78] | N=469 school students (251 female), 10-20 years, Germany | Three weeks post onset (school days), 2008 | Two weeks pre onset (school days), 2008 | n/a | ↑ daytime sleepiness | No test indicated but comparison of means described. | L |
| Lahti (2008)[79] | N=9, Finland | 4 days post transitions, 2005-06 | 4 days pre transitions, 2005-06 | ↓ efficiency<br>↑ movement, fragmentation, regardless of chronotype | - efficiency<br>↑ movement, fragmentation, regardless of chronotype | Non-parametric tests for related samples. | L |
| Lahti (2006b)[74] | N=10 (6 female), 32-70 years, Finland | 3 days post offset, 2003-04 | 3 days pre offset, 2003-04 | n/a | ↓ duration, efficiency<br>- movement, fragmentation | Paired Student's t test | L |
| Monk (1980)[76] | N=59-73 (~55% female), 18-79 years, UK | 5 days post transitions, 1976-77 | 5 days pre transitions, 1976-77 | ↑ sleep offset | ↓ sleep offset | ANOVA | L |
| Monk (1976)[82] | N=65 (55 female), UK | 6 days post offset, 1974 | 6 days pre offset, 1974 | ↑ sleep offset | | Paired Student's t test | L |

L=low, M=medium, H=high

##### *(ii) Acute transition effects on circadian biology*

There are six studies pertinent to this sub-section, all of whom report on DST-Onset effects but only four of whom report on DST-Offset effects. For a tabulated overview, see Sleep-Table 2. One study is rated as medium quality (Kantermann et al. 2007; described above) and the rest as low quality. Kantermann et al. (2007) observed an advance and a delay in the phase of wrist actimetry following DST-Onset and DST-Offset, respectively.[69]

Among the low quality studies, some phase changes in activity and wrist temperature were observed following DST-Onset but not DST-Offset.[70 83] Phase relationship changes cannot be confirmed though; Arguelles-Prieto et al. (2022) find that wrist temperature peaks and activity troughs are already changing pre transitions, which disallows a conclusion of transitions affecting internal synchrony in these eight participants.[70] Miguel et al. (2013) state using a measurement period of ten days pre and post transition and provide mean acrophase values; however, as means are used, we do not know if the individual relationships between wrist-temperature and activity rhythms are back "in synch" by day ten (assuming the phase relationship pre DST-Onset is "optimal/correct") in these five participants.[83] The study of US medical interns by Tyler et al. (2021) describes increased social jet lag following DST-Onset but only for individuals with a polygenic score indicative of being a later sleeper.[73] Lastly, studies from Finland in 19 participants by Lahti et al. (2008, 2006a) reported decreased inter-daily activity variability in short sleepers and increased inter-daily activity variability in long sleepers (presumably, "intra-"daily variability as a measure of rhythm fragmentation is intended) following DST-Onset only.[79 84] Of course, physical activity, which is highly dependent on behaviour and social responsibilities, may not be a useful marker of internal rhythmicity and circadian phase anyway.

Overall, transitions may advance or delay rhythm timing, but there are no indications of differences in phase relationships between internal rhythms caused by transitions.

**Sleep-Table 2: Acute transition effects on the circadian system, in order by quality and chronology**

| Author (Year) | Population/ Data | Index Period/ Intervention | Reference Period/ Comparator | Outcome ("↑"=increase/delay; "↓"=decrease/advance, "–" =no change) DST-Offset | DST-Onset | Statistics | Rating |
|---|---|---|---|---|---|---|---|
| Kantermann (2007)[69] | N=50 (34 female), 18-59 years, central Europe | 4 weeks post transitions (2006-07) | 4 weeks pre transitions (2006-07) | ↓ **c**entre of activity but not among each other either side of transition | ↑ **c**entre of activity but not among each other either side of transition | RM-ANOVA | M |
| Arguelles-Prieto (2022)[70] | N=8 (4 female), 33±11 years, Spain | 1 week post transitions (2017-18) | 4 days pre transitions (2017-18) | - wrist temperature phase marker, motor activity phase marker, phase relationship | ↓ wrist temperature phase marker, motor activity phase marker<br>- phase relationship | Student's t test & RM-ANOVA | L |
| Tyler (2021)[73] | N=554 medical interns, USA | 1 week post onset, 2019 | 1 week pre onset, 2019 | n/a | ↑ social jet for those with a later mid sleep polygenic score | T-test | L |
| Miguel (2012)[83] | N=5 (4 female), 20-46 years, Brazil | 10 days post transitions (2010-11) | 10 days pre transitions (2010-11) | - wrist temperature phase marker, motor activity phase marker, phase relationship | ↓ wrist temperature phase marker, motor activity phase marker,<br>change in phase relationship | Single Cosinor analysis, Wilcoxon matched test | L |
| Lahti (2008)[79] | N=9, Finland | 4 days post transitions (2005-06) | 4 days pre transitions (2005-06) | - activity relative amplitude, inter-daily variability, inter-daily stability, no apparent difference by chronotype | - activity relative amplitude, inter-daily variability, inter-daily stability, no apparent difference by chronotype | Non-parametric tests for related samples. | L |
| Lahti (2006a)[84] | N=10, Finland | 5 days post offset (2003-04) | 5 days pre offset (2003-04) | n/a | - activity relative amplitude, inter-daily variability, inter-daily stability, or tau by short vs long sleepers or by chronotype.<br>↓ inter-daily variability in short sleepers,<br>↑ inter-daily variability in long sleepers. | Paired Student's t test | L |

L=low, M=medium, H=high

##### *(iii) DST vs Standard Time (and longitude) effects on sleep*

One study from central Europe is rated high quality,[33] another study from Turkey is rated as medium quality (both study longitude differences within the same time zone),[85] and n=6 studies are rated as low quality. Of these six studies, three (ATUS, Czech Republic, Central Europe) consider longitude differences within the same time zone[86-88] and one compares years of transitions with years of perennial DST or perennial Standard Time in Russia.[32] The remaining two are somewhat different. One assessed the relationship between sleep, sun time, and time zone switches in neighbouring time zones in the USA.[89] The other study indicated comparing DST to Standard Time differences without indicating the actual measurement periods (location unknown).[90] The latter is also the smallest and oldest study (by 74 years). For a tabulated overview, see Sleep-Table 3.

The high quality rated study (high because of similarity in populations) was by Jankowski et al. (2014), who assessed sleep in two comparable student populations in Warsaw (~21.1 °E) and Heidelberg (~8.6°E); the key difference being residence at difference longitudes within the same time zone and at approximately equivalent distances from the longitudinal meridian.[33] The authors use ANCOVA with country as a factor and sleep parameters as dependent variables (measured by questionnaire of the previous seven days of 16-22 and 26-28 November 2012 in Poland and Germany, respectively; i.e., during Standard Time and ~3-6 weeks post transition). The eastern sample went to bed later (~10mins) and got up earlier (~20mins) with overall sleep duration difference of ~30mins between groups. The western sample went to bed later and got up later on free days but with no significant difference in sleep duration compared to the eastern sample. Thus, there were no differences in weekday mid-sleep but an ~25min difference in sleep debt-corrected mid sleep phase on free days (that corresponds to ~2mins delay per longitudinal degree westward; albeit, this was not measured). The work day vs free day difference may point to cultural differences. The medium quality study by Masal et al. (2015) identified a delayed mid-point of sleep by ~4.29mins per longitudinal degree westward on weekends among 15,362 elementary and secondary school students (aged 11-18 years) in Turkey (model adjusted for age, urban vs rural, school start time, and latitude).[85] The study was conducted from December 2012-March 2013; thus, during Standard Time and with the sun zenith occurring closer to noon for the more westerly population. There was no change in average sleep duration. As weaknesses, there were no descriptive statistics by longitude nor accounting for season.

Among the lower quality studies to assess longitudinal differences, Fischer et al. (2022) found mid-sleep time on weekends was delayed from east to west in three of four US time zones by ~ 2-mins per longitudinal degree using data from the ATUS (2003-2014) and univariate regression.[86] They used three different methods to capture the exposure of "average" longitude per state, all giving similar results. States spanning more than one time zone were excluded. Sladek et al. (2020) identify a significant correlation between east-west longitude difference and later sleep debt-corrected mid sleep phase on free days in the Czech Republic based on data from two surveys during 2015-2018.[88] However, the apparent difference is ~4 hours per half degree of longitude and the correlation, albeit significant, is near null (r=-0.095). Randler et al. (2009) identified a correlation between corrected mid-sleep times and longitude (r=-0.33, p<0.001) in adolescents at German speaking schools in Germany and abroad but within the same time zone.[88] The inverse correlation indicates delays with each degree west but specific sleep onset and offset times differences cannot be determined from the article.

Regarding the study in Russia, to compare years with biannual transitions (01 Jan 2009 to 26 Mar 2011) to years with perennial DST (27 Mar 2011 to 26 Oct 2014) or perennial Standard Time (27 Oct 2014 to ~Mar 2016), Borisenkov et al. (2017) compiled several different cross-sectional surveys of school and university students.[32] Survey locations spanned 11 degrees of latitude and 28 degrees of longitude (all within the same time zone), which were included in ANCOVA analyses as covariates alongside sleep and social jet lag data as dependent variables. They found sleep duration was shortest on perennial Standard Time (7.34 hours) but longest in years with transitions (7.69 hours). Free day sleep onset, latency, and rise times were latest/longest on perennial DST and work day rise time was latest in years with transitions.

In a study of sun time and time zone switches in neighbouring time zones, Hamermesh et al. (2008) find that the probability of being asleep at 07:00-07:10 and neighbouring times is associated with hour of sunset but not sunrise (adjusting for, *inter alia*, time zone and sleep hours) but the effect size is very small.[89] They use data from the ATUS (2003-2004) and only from DST observing regions. Furthermore, time zones appear to affect the relative probability of an individual being asleep at a given time of day, and more so for non-workers compared to workers and for financial/information workers compared to workers in other services in the morning hours. Using a difference-in-differences approach, they find that residents in non-DST practicing regions shift their sleep when neighbouring regions transition to DST. Specifically, they show that the probability of being sleep at 07:30-07:45 is lower and at 22:30-22:45 is higher (similar for neighbouring times) in a non-DST practicing region when neighbouring regions have transitioned (i.e., appears to be irrespective of sun times). The authors observe similar effects in Australia (Time Use Survey 1992) comparing the state of Queensland with New South Wales and Victoria; namely, when New South Wales and Victoria transition to DST, residents in Queensland shift their sleep onset and/or offset. This appears to be irrespective of sun times and television times. The non-repeated measures across transitions and that sample groups may include shiftworkers are limitations.

Lastly, Reese (1932) assessed sleep duration in 13 young children (2-5 years) at an 'institution' (presumably an orphanage) and found no apparent differences between measurements (timing based on eyes closed and few gross movements and steady breathing until wake) made during Standard Time or DST.[90] No further details, such as dates of assessment, are provided. This study neither adds or takes from any evidence base.

Overall, the 2-4mins differences and direction of shift in mid-sleep times by degree of longitude (or 30-60mins per 15° degrees which corresponds to a 1-hour difference in sun phase) that are observed in high and low quality studies fits with apparent sleep onset ad offset timing shifts and directions following the sun phase timing differences that occur with DST-Onset and DST-Offset.[33 85-88] The study designated as high quality suggests a shorter sleep duration for students (on work days only) further east of the meridian compared to a comparable distance further west of the meridian during Standard Time in central Europe while no difference in sleep duration was observed among adolescents during Standard Time in in Turkey.[33 85] As DST implements an analogous 1 hour "westward" shift in the clock times of the sun phases, we might then expect longer sleep durations during perennial DST in winter (or at least in students in central Europe and on work days, but cultural differences may play a role here). In line with the finding in central Europe, the study in Russian students suggests shortest sleep durations during years with perennial Standard Time compared to perennial DST.[32] However, longest sleep durations were identified in years with biannual transitions.[32] Lastly, there is suggestion that regions without DST that neighbour regions that change to DST may also alter their sleep onset and offset to be in line with DST anyway.[89]

In conclusion, and albeit based on limited evidence, a study of central European students implies perennial DST in winter months may be beneficial to sleep duration whereas a study of Russian students implies biannual time changes may facilitate longer sleep durations. A study in the USA suggests that sleep onset and/or offset differences (and, thus, possibly duration differences) may remain in non-DST practicing regions if neighbouring regions still employ biannual time changes.

##### *(iv) DST vs Standard Time (and longitude) effects on the circadian system*

Five studies are in this subsection, one high quality and the rest low quality. Two studies report on social jet lag, two studies on chronotype differences across longitude, and one study reports on cortisol rhythm at the population level (not individual level). All are described in more depth above apart from two low quality studies by Shawa et al. (2016) studying chronotype in South African adults and Hadlow et al. (2013) studying cortisol in Australia. For a tabulated overview, see Sleep-Table 4.

The higher quality study by Jankowski et al. (2014) does not indicate a difference in social jet lag or chronotype between more easterly and westerly longitudes.[33] The study by Borisenkov et al. (2017) describes most social jet lag in years of perennial DST and least in years of perennial Standard Time, but there was no statistical difference between years of perennial Standard Time and years with biannual transitions.[32]

The study by Randler et al. (2009) identifies a correlation, albeit weak, between morningness scores and longitude (more morningness further east; $r=0.11, p<0.001$).[87] Shawa et al. (2016) report a similar finding in South African adults.[91]

Hadlow et al. (2013) categorized single individual cortisol measurements collected at different times of day (07:00 – 17:00; one time of day per person) into two groups; namely, those collected in weeks 44-53 and 1-13 (summer months in Australia) of the year in years when Standard Time was enacted (2000-2005; 2010-2012) and in years when DST was enacted (2006-2009).[92] They fit cosinor curves to the two population level rhythms and found no differences in mesor or amplitude but a difference in acrophase. It is unclear what statistical analyses were used to compare the two mesors, amplitudes, or acrophases. Furthermore, the acrophases appear to occur within the 14 hours of the day wherein no cortisol measurements were taken.

Overall, eastern populations within the same time zone appear more likely to be more morning chronotypes, which fits with sleep onset and offset timings reported in subsection 3. Albeit weak evidence, perennial DST may be more detrimental than perennial Standard Time or biannual time changes for the circadian timing system across the year.

**Sleep-Table 3: DST vs Standard Time (and longitude) effects on sleep, in order by quality and chronology**

| Author (Year) | Population/ Data | Intervention /Comparator | Outcome ("↑"=increase/delay; "↓"=decrease/advance, "–" =no change) | Statistics | Rating |
|---|---|---|---|---|---|
| Jankowski (2014)[33] | For Warsaw and Heidelberg respectively: N=291 (82.8% female), n=279 (77.8%) female, 18-28 years, same class times, Poland and Germany | Different longitudes within a time zone | ↑ Duration from east to west. | ANCOVA | H |
| Masal (2015)[85] | N=15,362 elementary and secondary students, 11-18 years, Turkey | Different longitudes within a time zone | ↑ Mid-sleep delayed by ~4mins from east to west.<br>- Duration. | Multivariate linear regression. | M |
| Fischer (2022)[86] | N=50,753 (American Time Use Survey), USA | Different longitudes within a time zone | ↑ Mid-sleep by ~2 mins per longitude from east to west. | Univariate linear regression | L |
| Sladek (2020)[88] | N=3277 survey participants (n=1793 female), 18+ years, Czech Republic | Different longitudes within a time zone | ↑ Mid-sleep per longitude from east to west but negligible correlation. | Pearson Correlation | L |
| Borisenkov (2017)[32] | N=7968 students (~57% female), 10-24 years, European North of Russia. | Period with biannual time changes: (01 Jan 2009 to 26 Mar 2011)<br>Period with pDST: (26 Mar 2011 to 26 Oct 2014)<br>Period with pST: (27 Oct 2014 to ~ Mar 2016) | ↑ Durations on DST.<br>↓ Duration on DSTp.<br>↑ Free day sleep onset, latency, and rise times on DSTp.<br>↑ Work day rise time was latest on DST. | ANCOVA | L |
| Randler (2009)[87] | N=1211 students, ~10-23 years, Central European Time Zone | Different longitudes within a time zone | More eastern longitudes were correlated with more morningness. | GLM | L |
| Hamermesh (2008)[89] | N= 4,763, ATUS 2003-04, USA & n=2,741 Australian TUS, 11992 | Regions with Standard Time vs regions with DST | Sleep onset and offset in regions that do not practice DST also changes when neighbouring regions change to from Standard Time to DST. | Probit model marginal effects and differences in differences. | L |
| Reese (1932)[90] | N=13 (5 female) children at in institution (likely orphanage), 2-5 years, location unknown | DST vs Standard Time (Year and dates unknown) | No apparent differences in sleep duration | N/A | L |

L=low, M=medium, H=high

**Sleep-Table 4: DST vs Standard Time (and longitude) effects on the circadian system. in order by quality and chronology**

| Author (Year) | Population/ Data | Intervention /Comparator | Outcome ("↑"=increase/delay; "↓"=decrease/advance, "–" =no change) | Statistics | Rating |
|---|---|---|---|---|---|
| Jankowski (2014)[33] | For Warsaw and Heidelberg respectively: N=291 (82.8% female), n=279 (77.8%) female, 18-28 years, same class times, Poland and Germany | Different longitudes within a time zone | -Social Jet lag, chronotype | ANCOVA | H |
| Borisenkov (2017)[32] | N=7968 students (~57% female), 10-24 years, European North of Russia. | Period with biannual time changes: (01 Jan 2009 to 26 Mar 2011) Period with DSTp: (26 Mar 2011 to 26 Oct 2014) Period with nDSTp: (27 Oct 2014 to ~ Mar 2016) | ↑ Social jet lag with DSTp. ↓ Social jet lag with nDSTp albeit not different to periods with biannual time changes. | ANCOVA | L |
| Shawa (2016)[91] | N=304 (~57% female), ~mean 37 years | Different longitudes within a time zone | ↑ Morningness in the East | Mann-Whitney test | L |
| Hadlow (2014)[92] | N=27569 (65% female), 459.9 (19.1) years, Western Australia | DST (2006-2009) vs Standard Time (2000-2005, 2010-2012) during summer (weeks 44-53 & 1-13) | In population level rhythms, a difference in cortisol acrophase but not mesor or amplitude is reported, but this is unclear from the Figure. | Unclear how two population level acrophases were compared. | L |
| Randler (2009)[87] | N=1211 students, ~10-23 years, Central European Time Zone | Different longitudes within a time zone | ↑ Morningness in the East | GLM | L |

L=low, M=medium, H=high, DSTp=perennial DST, nDST,=perennial Standard Time

**Sleep-Table 5: JBI quality indicators**

(Questions and possible answers are shown in detail in Table footnote)

| Author (Year)[a] | Q1 | Q2 | Q3 | Q4 | Q5 | Q6 | R | Notes |
|---|---|---|---|---|---|---|---|---|
| Arguelles-Prieto (2022)[70] | Y | Y | N* | Y | U† | Y# | L | *Short sleep pre transition & possible bias by holidays, #very long sleep post DST-Offset, †t-test not paired although they use RM-ANOVA, no mention of children. |
| Barnes (2009)[56] | Y | U# | U# | Y | Y | N | L | #Non-repeated measures across transitions and the sample group may include e.g., shiftworkers. |
| Bano (2014)[72] | Y | N† | N* | Y | N# | N | L | †No intra-individual comparisons, *various diseases, unclear how close measurement was to DST-Onset time, compliance bias, #RM-ANOVA was used but pre and post DST-Onset were different groups of participants. |
| Borisenkov (2017)[32] | N* | U# | N# | Y | Y | N | L | *Unclear exactly when data were collected. #Similar, but not matched, and no repeated measures. |
| Fetter (2014)[24] | Y | Y | Y | Y | Y | N | H | |
| Fischer (2022)[86] | Y | U# | N* | Y | N* | N | L | #No descriptive statistics by longitude. *No accounting for season or latitude |
| Hadlow (2013)[92] | Y | U* | U* | Y | N# | Y# | L | *Matched somewhat by time of year but not by socio-dem. Some were likely ill. #Population level rhythms had to be used, and the population level acrophase appears to be interpolated to time periods wherein no data was available (between 17:00 and 07:00). It is also unclear how acrophases were compared to produce a statistically significant result. |
| Hamermesh (2008)[89] | Y | U# | U# | Y | Y | N | L | #Non-repeated measures across transitions and the sample group may include e.g., shiftworkers. |
| Harrison (2013)[80] | Y | Y | Y | Y | U* | Y* | L | *Two different analyses without reason, no repeated measures indicated in the second, regression to the mean a possible issue, figure indicates an effect on Sunday which was not measured. |
| Heacock (2022)[67] | Y | Y | Y | Y | Y | N | H | |
| Jankowski (2014)[33] | Y | Y | U# | Y | Y | N | H | #Cannot speak to light exposures but similar populations otherwise. |
| Jin (2020)[11] | N* | U† | U† | Y | Y | N | L | *The sleep duration question does include recency. †The survey was "representative", but only a selection was used and there were no intra-individual comparisons. |
| Jin (2015)[71] | N* | U† | U† | Y | Y | N | L | *The sleep duration question does include recency appropriately. †The survey was "representative", but only a selection was used and there were no intra-individual comparisons. |
| Kantermann (2007)[69] | Y | U† | Y | Y | Y | N | M* | †Unclear for database study. *In addition to "U" for Q2, part 2 of the study considered only one of each transition. |
| Lahti (2008)[79] | Y | Y | Y | Y | U† | Y# | L* | #Authors claim different to what is shown in graphs. †No actual tests mentioned. *Low sample size, then further stratified. |
| Lahti (2006a)[74] | Y | Y | Y | Y | Y | N | L* | *Low sample size, then further stratified. |
| Lahti (2006b)[84] | Y | Y | Y | Y | Y | N | L* | *Low sample size. |
| Masal (2015)[85] | Y | U# | U* | Y | U* | N | M | #No descriptive statistics by longitude. *No accounting for season. Effect modification requires testing. |
| Medina (2015)[68] | Y | Y | Y | Y | Y | N | H | |
| Miguel (2012)[83] | Y | Y | Y | Y | Y | N | L* | *Low sample size. |
| Monk (1980)[76] | Y | Y | Y | Y | Y | N | L* | *Single measurement and 1 transition period, limited population description |
| Monk (1976) | Y | Y | Y | Y | Y | N | L* | *Single measurement and 1 transition period, limited population description. |
| Oskardottir (2022)[81] | Y | Y | U* | Y | U* | Y# | L | *Analyses of four individuals independently, no data provided. |
| Quintilham (2014)[77] | Y | Y | Y | Y | U† | Y# | L | †No mention of paired analyses, #wake-time post DST-Onset did not correspond with being in a 07:30 class. |
| Randler (2009)[87] | Y | U# | U# | Y | Y | N | L | #Only age described by location and school language by longitude. No accounting for season or latitude in analyses. |
| Reese (1932)[90] | Y | Y | U* | Y | N+ | Y" | L# | *Not specified. #Low sample size. +Study was conducted pre-computer age. "No dates of study are provided. |
| Shawa (2016)[91] | Y | U* | U* | Y | N* | N | L | *There was no adjustment for age or sex or ancestry (or gene differences, which would have been possible) in the analysis. |
| Schneider (2009)[78] | Y | Y | Y | Y | U* | N | L | *They are not described. There is also no mention of repeated measures. |
| Sladek (2020)[88] | Y | U* | U* | Y | N* | N | L | *No matching, adjustment, population descriptions at different longitudes, no mention of being ill or shiftworkers. |
| Tonetti (2013)[75] | Y | Y | Y | Y | Y | N | L* | *Low sample size. |
| Tyler (2021)[73] | Y | U# | U# | Y | Y | Y" | L | #Non-repeated measures across transitions and the sample group includes shiftworkers. "The later midsleep group apparently had an earlier sleep onset on Friday nights pre transition. |

**JBI Critical Appraisal Checklist for Quasi-Experimental Studies (Adapted)**
Answers can be either Y=Yes, N=No, U=Unsure, or N/A.
R=Rating, High=High, M=Medium, L=Low

1. Is it evident which factor is considered the 'cause' and which one is the 'effect'? Is there no ambiguity regarding the sequence of variables under investigation?
2. Were the characteristics of the study participants involved in comparisons adequately matched?
3. Were the exposures/treatments (beyond that of interest in our study) of the study participants involved in comparisons adequately matched?
4. Were the methods used to measure the outcomes the same for participants who were part of different comparisons?
5. Were appropriate statistical methods employed to analyse the data?
6. Are there any irregularities in the data reported?

[a]As a re-analysis of previous studies, Putilov et al. (2020), is not included here. Due to severe limitations, Tarquini et al. (2019) is not included here.

***Conclusion***

With an opportunity for increased sleep duration at DST-Offset and possible longer sleep durations across the year more generally, biannual time changes may prove better than perennial Standard Time or perennial DST. Compatible with the above, Putilov et al. (2020) consider data from previously published studies to estimate what they define as 'actual sleep loss' under DST compared to Standard Time and concluded mild sleep loss (if any).[65] In their review, actual sleep loss was computed as a percentage: 100*([weekend rise time-weekday rise time]/[weekend rise time-weekday bed time+24]). From re-analysis of a seventh study (Borisenkov et al. 2017 – also included in synthesis above), they conclude little-to-no sleep loss during perennial DST compared to perennial Standard Time. The authors appropriately highlight the seasonal reduction in sleep and the difficulty in isolating what might actually be sleep "loss". The latter point also fits with the studies by Kantermann et al. (2007) and Heacock et al. (2022) that indicate a seasonality to sleep onset and offset and duration that either may not be apparent when only a few weeks around transitions are studied due to time resolution. Also, variability in sleep within and between individuals may also play a role.[67 69]

Overall, there appear to be more benefits to sleep and timing when a paradigm that includes DST-Offset transitions and Standard Time during winter months is in practice, but evidence is limited. Sleep onset and offset may advance at DST-Onset (relative to the reference sleep phases, not based on local clock times), but advances appear to be occurring with seasonal changes anyway. Adolescent school students and university students with early start times may be susceptible to an acute but relatively minor decrease in mean sleep duration at DST-Onset – which suggests that some populations may be more vulnerable than others but reasons are unclear (e.g., later chronotypes, early start times, already sleep deprived, etc). Low quality studies and contrasting findings disallows further conclusions.

#### 6. Cognitive Section Synthesis

Jonas P. Wallraff & Philip Lewis

##### *Introduction*

Systematic screening returned 19 articles that we categorize as 'Cognitive' because the outcomes studied therein pertain (more or less) to multi-faceted aspects of thinking and self-regulation (e.g., attention, perception, problem-solving, decision-making, impulse-control) to guide behaviour and emotion. Furthermore, the specific tests add another layer of differentiation. As such, this synthesis is necessarily longer than for other sections.

This section includes eight subsections with some studies included in more than one subsection. Concerning effects of transitions, subsections include: (i) attention (three articles), (ii) perception (three articles), (iii) task performance (three articles), (iv) criminal activity (four articles), (v) non-criminal behaviour change (three articles), and (vi) wildfire and dwelling fires (two articles). Human activity-related wildfires and dwelling fires are included here based on potential roles for attention, risk perception, decision-making, and impulsivity in their cause. We include one subcategory dealing with studies that (vii) compare regions with transitions to regions without (two articles). We also include one category on (viii) stock market returns (two articles), for which we identified a further thirteen articles after citation searching. We find that the current subsections best suit our goal of comprehensive and coherent syntheses; however, as detailed syntheses are long, we first provide a summary.

Additionally, we provide an overview of studies and their final quality rating in Cognitive-Table 1 (aligned with the subsection order in text). JBI quality indicators are presented in Cognitive-Table 2. Stock market return studies are not included in Cognitive-Tables 1 or 2 as they do not lend well to simple overview due to the volume of markets, tests, comparisons, and elaborations on limitations. Nonetheless, the strengths and limitations of stock market return studies are synthesized at length in the text.

##### *Summary Synthesis*

Of the non-stock market return studies, we identify eight low quality studies, two medium quality studies, and seven high quality studies, but none are without limitations. To be rated as high quality, we deemed some form of robustness tests, such as placebo or control tests (e.g., comparing two or more non-DST periods wherein there is no expectation that they could be influenced by transitions to and from DST) as a general pre-requisite alongside higher quality answers to other checklist questions. Medium quality rated studies included issues with reference periods and non-individual level data. Low quality studies included potential for misclassifications of exposure or outcome, very low samples sizes, and only one DST-Onset or DST-Offset transition studied.

The findings of the seven high quality studies were as follows: riskier driving simulator performance coupled to a reaction time increase of 0.37s from 1.80s to 2.17s was observed in young men following DST-Onset in Italy, but whether this translates to a higher stakes real world setting is unknown.[93] An ~7% drop in robbery incidence per million following DST-Onset was observed in a study of three years of USA National Incident-Based Reporting System records; however, the effect was significant only at the 10% level, and may be driven by sunset hour differences rather a cognitive change.[94] In absolute numbers, this crudely translates to ~27 fewer robberies per 10 million people over the 3 weeks bandwidth used in the study (assuming 66.1

robberies per 100,000 people in 2022[A]).[94] An ~16% increase in crime-related fatalities was observed in Mexico following DST-Offset but not DST-Onset in a study of database records over 20 years;[12] however a possible role of the change in sunset timing is a valid question here too.[94] In (crude) absolute terms though, this equates to ~25 more fatalities in the week following DST-Offset.[12] The study of dwelling fires in the UK reports no apparent transition effect and the study of wildfires in the USA suggests a possible DST-Onset increase but this may be due to a change in the clock time of sun phases that allow for more drying time before ignitions.[95 96] One study identified ~5% longer imprisonment sentencing lengths following DST-Onset, but this was hampered by positive placebo tests.[97] If true, a usual 10 year imprisonment sentence might be expected to be 6 months longer on the Monday following DST-Onset.[97] Yet, such longer sentences are not longer than are given on regular Fridays (when sentences also tend to be stricter);[97] so post transition Monday may be no more or less fair than a regular Friday. The final higher rated study finds ~10% fewer charitable donations made over the "DonorsChoose" online database in the week following DST-Onset but not DST-Offset;[98] however, Easter holidays may bias donation giving amounts/volume or donation giving may change to a different method.

Of the medium quality studies two, one study focusing on acute effects (~3% more and fewer assaults in the USA following DST-Offset and DST-Onset, respectively) was hampered by positive placebo tests and not using pre transition dates as reference periods.[99] Furthermore, and according to the authors, the 3% difference would equate to 330 more or less assaults per year of the ~4 million total, which is insubstantial at the population level.[99] The second study found ~16 points worse mean school SAT (scholarly aptitude test) scores in a region that practices DST compared to a region that does not; however, this is not individual level data.[100] The author extrapolates an economic effect of ~US $200 less earnings per person per year.[100]

Findings among low quality studies are mixed and, thus, overall do not add to the debate about whether transitions or living with DST compared to Standard Time affect cognitive function.

The stock market studies can be followed chronologically wherein a potential transition-associated anomaly was initially identified in the USA[101] but later ruled out by various robustness tests and tests in different markets.[102] The stronger DST-Offset anomaly may also be an artefact of a general October anomaly in mean market returns.[102] One recent study that focuses on returns from mergers rather than mean returns more generally also identified an DST-Offset effect,[103] but it remains to be seen if this is also linked to the October anomaly.

In conclusion, there is insufficient evidence to abandon DST practices based on the totality of the evidence from the studies in this section. Studies with regression discontinuity designs suggest some effects of transition may last longer than a week. Substantial effect sizes from some studies suggest more comprehensive investigation is warranted.

[A] https://www.statista.com/statistics/232564/robbery-rate-in-the-us-by-state/

**Cognitive-Table 1: Overview of studies in Cognitive Section, by subcategorizations, quality rating, and chronology**

| Author (Year) | Population/ Data | Index Period/ Intervention | Reference Period/ Comparator | Results | Statistics | Rating |
|---|---|---|---|---|---|---|
| ***Attention*** | | | | | | |
| Maier (2021)[104] | Germany, n=101 employees, 20-66 years, 67% female, 64% white collar. | First Monday post DST-Onset, 2021. | Monday pre DST-Onset, 2021. | Inverse sleep quality (PSQI-single question) association with procrastination (Tuckman) but only in later chronotypes and only post DST-Onset. | Nested hierarchical modelling with interaction terms, simple slope post hoc analyses. | L |
| Schaffner (2018)[105] | Australia, neighbouring populations in NSW (n=47) & Queensland (n=91). | DST-Onset day, 2013 | Sunday pre DST-Onset, first Sunday post DST-Onset, 2013. | No difference in online Stroop test. | T-test, multivariate OLS regression with interaction terms (differences-in-differences). | L |
| Wagner (2012)[106] | USA, 203 metropolitan areas, Google database | First Monday post DST-Onset, 2004-2009 | Monday pre DST-Onset and 2nd Monday post DST-Onset, 2004-2009 | ~3-5% increase in relative volume of searches categorised as entertainment (used as proxy for cyberloafing) | Hierarchical linear modelling with weighting by metropolitan area. | L |
| ***Perception*** | | | | | | |
| Cho (2017)[97] | USA, Sentencing Commission Data | First Monday post DST-Onset, 1992-2003. | Monday pre DST-Onset, Second Monday post DST-Onset, 1992-2003. | ~5% longer sentences | Hierarchical modelling with covariate adjustment. Includes robustness test. | M |
| Schaffner (2018)[105] | As above. | As above. | As above. | No difference in online lottery choice (risk perception). | As above. | L |
| Hicks (1980)[107] | USA, n=1 teacher, n=14 male and n=12 female students. | First Tuesday post DST-Offset, 1980. | First Tuesday pre DST-Offset, 1980. | Perception of both sexes behaviour changed but in different directions. | RM-ANOVA, T-test. | L |
| ***Task performance*** | | | | | | |
| Orsini (2022)[93] | Italy, n=45, healthy males (24.1 ±3.6 years), ≥ 1-year driving experience, full licence, ≥1000 average annual miles. | Intervention group trials: Mar 22th-25th (pre DST-Onset) and 29th-Apr 1st (post DST-Onset) 2021. | Control group trials: Mar 8th-11th and 15th-18th (both pre DST-Onset) 2021. | Impaired driving simulator performance/riskier behaviour post DST-Onset | McNemar test, Fischer Test, RM-ANOVA. | M/H |
| Herber (2017)[108] | Europe, elementary school students at similar level (~10 years old) in Sweden, Norway, Lithuania, Finland, Denmark, Norway, and Spain, (n=8,813 TIMSS, n=13,255 PIRLS). | First week post DST-Onset, 2011. | One week pre DST-Onset, 2011. | No differences in math, science, reading scores. | Hierarchical modelling with stratification by sex, country, test language spoken at home, or number of books at home. Robustness tests used. | L |
| Monk (1980)[109] | UK, n=39 university students and workers from London. | First week (Monday-Friday) post DST-Offset, 1977 | Two weeks pre DST-Offset and second week (Monday-Friday) post DST-Offset, 1977 | Faster mathematic calculation times on the Wednesday and Thursday only. | One-tailed t-test. | L |
| ***Criminal Activity*** | | | | | | |
| Goodwin (2023)[12] | Mexico, General Directorate of Health, Crime Fatalities | Optimal bandwidth selection pre and post transitions, 1998-2018 | | ~16% increased fatalities post DST-Offset but not post DST-Onset | Regression discontinuity, various robustness tests. | H |
| Doleac (2015)[94] | USA, National Incident-Based reporting System | Optimal bandwidth selection pre and post DST-Onset, 2005-2008 | | ~7% drop in robbery incidence per million following DST-Onset, significant at the | Regression discontinuity and differences-in-differences, other robustness tests also used. | H |

**Cognitive-Table 1: Overview of studies in Cognitive Section, by subcategorizations, quality rating, and chronology**

| Author (Year) | Population/ Data | Index Period/ Intervention | Reference Period/ Comparator | Results | Statistics | Rating |
|---|---|---|---|---|---|---|
| | | | | 10% level, apparently driven by sunset hour differences | | |
| Umbach (2017)[99] | USA, National Incident-Based reporting System & local databases for Chicago, Los Angeles, and New York | First Monday post transitions, 2001-2014 | Second Monday post transitions, 2001-2014 | ~3% fewer assaults following DST-Onset and ~3% more assaults following DST-Offset, but robustness tests challenged the DST-Offset finding | Poisson quasi-maximum likelihood estimator regression model, Years with public holidays at transition time were excluded, robustness tests used. | M |
| Lindenberger (2019)[15] | Germany, Hesse, Forensic autopsy reports | Two weeks post transitions, 2006-2015 | Two weeks pre transitions, 2006-2015 | No differences in deaths by drugs/intoxication, homicide, or malpractice | $\chi^2$ | L |
| ***Non Criminal Behaviour Change*** | | | | | | |
| Ben Simon (2022)[98] | USA (DST practicing states only), DonorsChoose database | First week post transitions, 2001-2016 | Four weeks pre and 2-4 weeks post transitions, 2001-2016 | ~10% fewer charitable donations in the DST-Onset week but not in the DST-Offset week | Linear regression, outliers removed and accounting for longer/shorter transition days, robustness tests used. | M/H |
| Barnes (2015)[110] | USA, Google Database | First Monday post DST-Onset, 2008-2013 | Monday pre and second Monday post DST-Onset, 2008-2013 | Lower relative internet searches using moral-associated search terms | Hierarchical linear modelling with time*search category interaction term. | L |
| Hicks (1980)[107] | As above. | As above. | As above. | Both sexes behaviour changed but in different directions. | As above. | L |
| ***Wildfires & Dwelling Fires*** | | | | | | |
| Kountouris (2021)[96] | UK, Home Office dataset | Post DST-Onset/DST-Offset, 2010-2018 | Pre DST-Onset/DST-Offset, 2010-2018 | No difference in incident dwelling fires | Regression Discontinuity, robustness tests were used. | H |
| Kountouris (2020)[95] | USA (DST observing states/years only), 4th editions of the USDA Forest Service Fire Program Analysis-Fire Occurrence Database | Post DST-Onset/DST-Offset, 1992-2018 | Pre DST-Onset/DST-Offset, 1992-2018 | ~30% jump in human-associated wildfires at DST-Onset, including in different strata of employment (~32%), leisure (~44%), and arson (~26%). Only leisure (~16%) significant for DST-Offset. | Regression discontinuity, taking seasonality and weather into account, robustness tests used. | H |
| ***DST vs No DST*** | | | | | | |
| Gaski (2011)[100] | USA, Indiana, n=~350 public high schools | DST practicing region, 1997-2006 | Non-DST practicing region, 1997-2006 | School mean SAT scores were ~16 points worse in DST practicing regions. | Least squares linear regression with weighting by school's number of test takers. | M |
| Schaffner (2018)[105] | As above. | As above. | As above. | No differences between NSW and Queensland. | As above. | L |

L=low, M=medium, H=high, TIMSS: Trends in International Maths and Science Study, PIRLS: Progress in International Reading Literacy Study, SAT: Scholarly Aptitude Test

**Extended Syntheses**

##### *(i) Attention*

Three studies concerning attention and focussing on the DST-Onset transition use three different outcome metrics and take place on three different continents. Schaffner et al. (2018) identified no effects of DST-Onset on attention in an online-based task in Australia.[105] Wagner et al. (2012) concluded that an increased relative percentage of internet searches designated as 'Entertainment' by the Google database post DST-Onset is indicative of increased "cyberloafing" at work in the USA.[106] Maier et al. (2022) found that DST-Onset might elicit/strengthen an association between sleep quality and procrastination at work in a questionnaire-based study in Germany, but only in late chronotypes.[104] Although the latter two study findings support a DST-Onset effect on attention, study limitations negate conclusion of a causal effect.

Schaffner et al. (2018) assessed neighbouring populations in Queensland and New South Wales (n=91 and n=47 study participants, respectively; 70km either side of the border, who share the same time zone until DST-Onset in New South Wales [NSW] on the first Sunday in October).[105] On the transition day, one week pre, and one week post transitions, participants completed the Stroop Interference Test (a series of colour words, with or without the same font colour as described by the word, participants must select the correct font from options). The NSW group performed better than the Queensland group on transition days ($p<0.05$, t-test), but multivariate OLS regressions find both groups improved compared to the prior non-transition day and there were no significant time*group interactions (i.e., difference-in-differences). Limitations include the potential learning effect and that the tests were not necessarily performed at the same times of day, although the authors attempted to control for the latter with a time-of-day covariate. The attrition from the first to last test was 307 to 138 participants and analyses were performed on the 138 participants only. Thus, there may be a selection bias insofar as those less affected by DST-Onset were those who participated in all three instances. However, this can be considered unlikely as the attrition percentage was similar in both territories.

Wagner et al. (2012) analysed normalised volume of internet searches classified as 'Entertainment', archived and provided by Google, for 203 US metropolitan areas, on the Monday pre DST-Onset, first Monday post DST-Onset, and second Monday post DST-Onset, for the years 2004-2009.[106] Normalised refers to being a percentage of all searches for that metropolitan area over a 3-month period (i.e., the February to April downloaded data). The data was weighted by metropolitan population estimates. Hierarchical modelling showed no differences between the non-transition week Mondays but transition week Mondays included ~3% and ~6% more 'Entertainment' searches than the pre and post Mondays, respectively. To speculate on potential sources of bias, transition leads to more 'Entertainment' related internet searches at home or during work breaks instead of socialising. Another weakness is the assumption that this excess in entertainment searches constitutes "cyberloafing" at work and that these searches are not a part of one's work.

Maier et al. (2022) assessed sleep quality (single-item PSQI, "1 question") and procrastination (Tuckman Procrastination Scale) in 101 employees from various workplaces around Germany (20-66 years, 64% white collar, n=67 female, non-shiftworkers) on the Monday pre DST-Onset and the Monday immediately post DST-Onset, immediately after each workday period (advised at least), in different chronotypes, by questionnaire.[104] Although the time covariate was not significant, a significant chronotype*time*sleep quality interaction in the best fitting model of nested hierarchical models led the authors to conduct simple slope analyses. In stratified data, they found a significant inverse association between sleep quality and procrastination in later chronotypes post DST-Onset but not pre DST-Onset. The same direction of association was observed for early chronotypes pre DST-Onset and opposite directions were observed for later chronotypes pre DST-Onset and for earlier chronotypes post DST-Onset, but these were not significant. The authors speculate that later chronotypes may be more prone to DST-Onset-induced depletion of self-regulatory resources, making them more reliant on sleep quality to avoid procrastination post DST-Onset. Concurrent measurement of subjective sleep quality and subjective procrastination remains an issue more generally, but is unlikely to explain differences between the chronotype*time strata. Mean procrastination scores appear very low for the different chronotype*time*sleep strata (the latter categorised by ± 1 standard deviations of mean sleep quality) with the highest score being 1.6 (out of a maximum of 5), which suggests procrastination may not be much of an issue in this sample more generally.

Taken together, transition effects on attention cannot be concluded.

##### *(ii) Perception*

Three studies on perception include perception of risk based on lottery choice (Schaffner et al. 2018),[105] perception of severity of criminal transgression and criminal based on legal sentencing (Cho et al. 2016),[97] and perception of children behaviour and manners in school based on the ratings of a teacher (Hicks et al. 1980).[107] More generally, no differences were observed in perception of risk following DST-Onset.[105] Potential differences in legal sentencing were observed following DST-Onset but not DST-Offset, albeit limited by a significant difference observed in a placebo test.[97] A sex*time interaction in children behaviour rating was observed, but this may reflect behaviour *per se* rather than a change in the teachers perception of behaviour, but this is also limited by potential regression to the mean.[107]

More specifically, the study by Schaffner et al. (2018) in NSW and Queensland (Australia, described above in the subcategorization 'Attention') also included a lottery choice task that was considered a measurement of perception of risk.[105] Participants had to choose one of a pair of lotteries to be played out in ten different rounds, with similar expected values but differences in odds of payoffs (i.e., greater risk might lead to greater reward; e.g., do you choose $10Aus guaranteed or do you choose a 50-50 chance of either $5Aus or $15Aus, etc.). Both groups became more risk averse over time but there were no group differences or time*group interactions. Limitations for this study are already described in (i) Attention.

Cho et al. (2017) compared the length of imprisonment sentences on the Monday post DST-Onset with the preceding and following Monday, between 1992 and 2003, in the USA (DST practicing states only).[97] Data on US citizens from the US Sentencing Commission was used in a hierarchical model with judicial district as level 2. Sentences rendered on the Monday post DST-Onset were ~5% longer compared to the preceding and following Mondays after accounting for age, gender, race, education, year, criminal history, type of trial, number of convictions, and offense level. They were also longer compared to the Wednesday and Thursday but not compared to the Tuesday or Friday in the same corresponding week. There were no differences between the Tuesday, Thursday, and Friday post DST-Onset and their corresponding days in preceding and following weeks. There were differences between the two reference Wednesdays but these were not different compared to the index Wednesday. The Monday finding was still observed after accounting for Federal holidays. Mondays immediately post DST-Onset were also different to all other Mondays in the year ($p<0.01$). There were no differences between the Mondays around DST-Offset. The main limitation is the finding of a significant difference in a placebo test (i.e., between the two reference Wednesdays).

Hicks et al. (1980) assessed a teacher's rating of behaviour and manners of students in class (n=14 males, n=12 females) on the Tuesday pre and Tuesday post DST-Offset in the USA (presumably near San Jose State University).[107] RM-ANOVA revealed no sex or time effect but there was a sex*time interaction. In stratified analyses by t-test, boys scored worse and girls better post DST-Offset ($p<0.05$). The finding is limited by possible regression to the mean (boys who started high scored lower and girls who started low scored higher), chance (one transition under study, no placebo tests), or may reflect actual changes in the behaviour of the children rather the teacher perception or be a combination of both. This study is also notably older – from 1980 – and may be less relevant today.

Taken together, transition effects on perception cannot be concluded.

##### *(iii) Task Performance*

Three studies include task performance that go beyond attention, use different tasks, are assessed in different populations, and in different decades. Monk & Alpin (1980) find greater calculation efficiency in the week following DST-Offset in the UK.[109] Herber et al. (2014) find no differences in student reading, science, and math performance scores within a European assessment program following DST-Onset.[108] Orsini et al. (2022) found that DST-Onset was likely to affect driving simulator performance in Italy in young adult males in Italy.[93]

Monk & Alpin (1980) provided a booklet covering the 4 weeks around transition with three calculations to be done each day (addition, multiplication, subtraction – all with 4-digit numbers) to 39 university students and workers from London who were also told to time themselves doing the calculations.[109] Subjects were blinded to the true purpose of the study and scores were normalised to a placebo test. Shorter than normal times were apparent in the test week and significant on the Wednesday and Thursday (one-tailed test) compared to corresponding days. The improvement is hypothesized to be linked to a lack of rhythm adjustment and that the task was performed at later "internal" times in the week following DST-Onset. However, the Wednesday and Thursday scores were also the lowest of the week in the reference weeks, and a training effect may bias findings.

Herber et al. (2017) used the Trends in International Maths and Science Study (TIMSS) and Progress in International Reading Literacy Study (PIRLS) results as a basis for their study.[108] In 2011 around the time of DST-Onset, these tests were administered to elementary school students at a similar school level (~10 years old) in Sweden, Norway, Lithuania, Finland, Denmark, Norway, and Spain (n=8,813 students for TIMSS, n=13,255 for PIRLS). Specific dates with individual schools were scheduled several months in advance by country coordinators; thus, there was some random allocation of schools and students to test dates either one week pre or one week post transition. Scores pre DST-Onset were compared with scores post DST-Onset and no significant differences were observed in pooled observations or after stratification by sex, country, test language spoken at home, or number of books at home, in placebo tests, or after adjustment for day of week. However, the TIMSS analysis was repeated for ~14-year olds in Finland with possible effects of DST on math ($p<0.1$) and on science ($p<0.05$) observed (but the effect size was not greater than was observed for ~10-year olds). A hierarchical linear regression model with maximum likelihood was used. The main limitation is that a difference-in-differences approach was not used and that the post transition participants may score higher than pre transition participants may if they had been assessed pre transition. It should be noted that these tests are not high-stakes tests for either school or student and that test times were not accounted for.

Orsini et al. (2022) assessed driving simulator performance (which involves, *inter alia*, attention and perception) in 45 young males (19-31 years) with no previous simulator experience.[93] Furthermore, they had normal or corrected vision, at least 1-year driving experience with a full licence, average annual mileage of at least 1000 miles, and had no known diseases or sleep disorders nor were taking medication known to affect sleep or reaction times. Participants were naive to the purpose of the study. The simulator involved 15 minutes with various driving tasks near and in Padova (Italy), which would be a common commute. The control group completed tasks on the same weekday and time (Mar 8th-11th and 15th-18th 2021, 08:00-10:30) as did the experimental group but the second trial in the experimental group was post DST-Onset (Mar 22th-25th and 29th-Apr 1st 2021, 08:00-10:30). On the day of the first trial, participants also completed a 5-minute training simulation. The route was identical in both trials. There were no differences between groups by age, driving experience, daytime sleepiness, night sleep quality, or chronotype. The experimental group overtaking violations increased from 3 to 9 and the control group from 1 to 2 across trials. The inter-trial difference was significant for the experimental group (McNemar test), as was the inter-group difference for trial 2 (Fischer exact test). There were no significant effects when stratified by chronotype. Repeated measures ANOVA with mid-sleep on free days as a covariate revealed a trial*group interaction with the experimental group performing worse. No interactions were observed for the following variables associated with overtaking a bicycle: ratio between distance and speed difference between driver and cyclist, rear and lateral passing distances, and mean passing speed. Regarding exiting from a freeway, interactions were observed for standard deviation of steering angle, mean lateral acceleration but not for maximum deceleration, speed, and trajectory. The control drivers appeared to perform better in trial 2 but the experimental drivers appeared to perform worse, especially in terms of risk assessment. Whether potential DST-Onset effects on simulator performance translate into the real world driving scenario remains open. Perhaps DST-Onset increases what might be considered riskier behaviour only when there is in fact no danger.

A transition effect on task performance may depend on factors such as the task *per se,* its difficulty, aspects of cognitive ability required, the population, and the time-of-day of task. In the studies above where differences were observed, the tasks were not high stakes and the study samples were small.[93 109] In the largest study, no differences were observed, but this study is also most limited by not involving intra-individual differences across the transition.[108] Taken together, transition effects on consequence-free calculation efficiency, math, science, and reading performance, and driving simulator performance cannot be concluded, but there is stronger suggestion of an effect in this subcategory than for attention or perception.

##### *(iv) Criminal activity*

Four studies to be synthesized under criminal activity include robberies[94] and assaults[99] determined from databases in the USA, crime fatalities determined from a database in Mexico,[12] and deaths due to drugs/intoxication, homicide, and malpractice determined from forensic autopsy in Germany.[15] Potential transition effects on cognition associated with criminality include impulse control and decision-making, which may result in increased crimes happening (which may be related to risk perception) and increased violence associated with crime (impulse control). An important limitation is the change in the clock time of sun phases that may affect opportunity and number of confrontations and probability of getting caught rather than cognitive processes. Conceivably, both are important in evaluation of this evidence line.

Doleac et al. (2015) use crime data from 2005-2008 obtained from the National Incident-Based Reporting System (NIBRS) in the USA to assess robberies pre and post DST-Onset using a regression discontinuity design.[94] Robberies are considered most suitable for such analyses, as they require confrontation (thus, event timing is likely known by the victim) and are financially motivated (though, of course, robberies gone wrong can be classified as rape, aggravated assault, or even murder). The authors find an approximately 7% drop in robbery incidence per million following DST-Onset, though significant at the 10% level only. According to the regression discontinuity figure provided by the authors, ~35 days were required for robbery rates to "return" to a level comparable to those observed prior to DST-Onset. Collapsing the incidents into the sunset hour and following hour (pre DST) versus all other times of day, they find a 27% decrease during sunset hour and hour immediately after specifically, significant at the 1% level. It is difficult to reconcile confounders that would systematically affect these specific hours. The authors also use a difference-in-differences approach with the same weeks in March that were non-DST in 2005 and 2006 but were DST in 2007 and 2008 (following USA policy change). They find 20% less robberies in the sunset hours compared to other hours in DST compared to non-DST, significant at the 1% level. Plotted DST vs non-DST differences over hours since sunset further point at the sunset hours as driving the difference. These analyses suggest that sunset timing is a causal factor, and that this may not be due to any cognitive–related effects. Robustness checks (different bandwidths, polynomials, restricted samples) are not all significant, possibly due to restrictions on the data used, but all estimates are within 1 standard error of the primary model. Placebo regression discontinuity tests indicate no effects at other times of year. DST-Offset analyses are considered less reliable due to Halloween and large standard errors. There is no DST interaction with an indicator for weekend when included in the model for robbery, but no effect on weekends could also be due to residual confounding from e.g., family time. Ultimately, the drop in robberies is conspicuous to the sunset hours, which may be more indicative of changes in opportunity rather than a cognitive change in offenders. Of note, Doleac et al. (2015) also find no differences in aggravated assault or murder.

Umbach et al. (2017) analysed assaults (that resulted in police involvement, including aggravated and simple assault) from the NIRBS and also from the cities Chicago, Los Angeles, and New York (as NIBRS disproportionately represents smaller jurisdictions) from 2001 to 2014.[99] They compared the Mondays post transition to the Mondays one week later (noting this provides the best control for sunlight-social schedules) with a Poisson quasi-maximum likelihood estimator regression model. Years that included holiday overlap with reference or index periods were discarded (Halloween 2005, St. Patricks Day 2008 and 2014). The authors found 3% fewer assaults on the Monday following DST-Onset, no moderating effect of age or sex, and falsification tests were null. They also found ~3% more assaults on the Monday following DST-Offset, with no moderating effect of age or sex. Robustness checks including a placebo test with subsequent Mondays, comparisons of Wednesdays and Thursdays, and using reverse coding for the 2006 dates for 2007 onward and *vice versa* were mostly null for DST-Onset but not for DST-Offset. However, the latter non-null findings may suggest a more prolonged effect of DST-Offset. Albeit with slightly different focuses and methods, the authors use similar databases and find similar results in terms of a drop in robberies and assaults respectively following DST-Onset, but Doleac et al. (2015) also find no differences in assaults.[94 99] Moreover, the robberies difference may be specific to the changed sunset hours,[94] while Umbach et al. (2017) find the drop in assaults across the whole day but use the second week post transition as the only reference period, wherein the clock time of sun phases will be the same as the index week.[99] With more nuanced studies, a transition effect on different crimes might be teased out, but it cannot be concluded from these studies.

Goodwin et al. (2023) studied criminal fatalities in Mexico (relatively stable photoperiod across year; various DST paradigms: no DST since 1998 in the region along the Arizona border, other USA border regions follow the current USA DST paradigm [i.e., transitions on the second Sunday in March and the first Sunday in November], no DST since 2015 on the Caribbean coast, the rest is the same as the older USA DST paradigm [transitions on the first Sunday in April and last Sunday in October]).[12] Individual level health and demographic data with daily resolution from 1998-2018 is recorded in the General Directorate of Health. No distinction was made on type of assault-mediated death or intent. Regression discontinuity methods reveal that crime fatalities do not appear to be affected by DST-Onset but appear to increase post DST-Offset. An increase in fatalities is observed for DST-Offset only (~16%), regardless of urban or rural locale and following a previous decline. The decline was observed in the 10 days prior to transition. It took ~10 days before crime fatality rates were again comparable to those observed prior to the decline (i.e., pre transition). However, within the documented 60-day period around transition, crime rates did not reach as low levels as observed in the week immediately preceding the transition. Using higher order polynomials and different bandwidths made little difference and placebo tests by time and place were null. These finding are in line with the DST-Offset increase in assaults observed in Umbach et al. (2017)[99]. Lindenberger et al. (2019) analysed autopsy data gathered over ten years (2006-2015) in the state of Hesse in Germany, and compared the two weeks pre transitions with the two weeks post transitions by $\chi^2$ testing, excluding the transition night.[15] Of course, some non-natural deaths may be attributable to cognitive issues, but it is not possible to be more specific. Traffic and occupational accidents and suicides are covered in other sections, but deaths due to drugs/intoxication, homicide, and malpractice may be due to cognitive change. No differences were observed for these for the time around DST-Onset or DST-Offset, but the counts of cases are low.

Overall, increased assault and assault leading to death post DST-Offset were observed in two studies from the USA and from Mexico, and placebo tests were null in both.[12 94] However, one study using similar data in the USA observed no change in assaults.[94]

##### *(v) Non-Criminal Behaviour Change*

Three studies are included here but only one at the individual level, by Hicks et al. (1980) (included above under 'Perception').[107] Their findings suggest a sex-modified effect of DST-Offset school student behaviour, but (as mentioned above) this is limited by only being a 1-time study, small in size, hampered by potential regression to the mean and potential DST effects biasing the observer. The other studies include a possible lower relative internet search volume of internet search for moral words[110] and lower charitable donations[98] following DST-Onset in the USA.

Barnes et al. (2015) compared 'moral awareness' (reflected in Google searches of moral words in the USA from 2008 to 2013, using the same method as previously described for Wagner et al. 2012,[106] but for the whole USA – for which DST is not wholly standardised) on the Monday following DST-Onset with the preceding and following Mondays.[110] The list of moral search terms used appears thorough and follows a rigorous approach to its development, including high interrater agreement on the moral content of the terms. The authors report no difference between the Monday following DST-Onset compared to the preceding and following Mondays but a significant interaction between day and search category (moral words vs Google General Categories), with moral search terms being lower (however, the search volume was already normalised, it is unclear why and how this interaction was included).

Ben Simon et al. (2022) analysed altruistic monetary donations in the USA (non-DST practicing states labelled as reference as appropriate) from 2001-2016 in the four weeks pre and four weeks post DST-Onset. However, the study did not account for the state of Indiana, which did not observe DST until 2006. Mean donation amounts in the week (specifically weekdays) pre DST-Onset were lower than in the preceding and following weeks (controlling for donation day, month, and year), assessed by multiple regression. No differences were observed in comparison of other weeks, no differences were observed in non-practising states, and no differences were observed around DST-Offset. Thus, it seems the effect is highly temporal, occurring solely as a direct result of the transition. The effects around DST-Onset remained after accounting for the number of donations (which could have been affected by the lost hour on the first Sunday of DST, i.e., reduced donating time). Donations were assessed via the 'DonorsChoose' website that helps to raise funds for school projects. Extreme outliers were removed (outside three standard deviations from the mean). Easter, which can fall close to DST-Onset and is a religious holiday that may stimulate donations and bias donation estimates toward zero if close to transition but not on index days or away from zero if on reference days, was also not accounted for.

Overall, a cognitive behaviour change resulting from transition cannot be concluded.

##### *(vi) Wildfires & Dwelling Fires*

The occurrence of dwelling fires and human-caused wildfires can also be linked to human cognition in terms of perturbed attention, risk perception, decision-making, and impulsivity. Kountouris (2021) examined incident dwelling fire data from the UK Home Office dataset (all incidents attended by Fire and Rescue Services in England from April 2010-April 2018).[96] The author also examined wildfires in the USA except pre-2006 Indiana, Arizona, Hawaii, and Alaska using data from the 4th edition of the USDA Forest Service Fire Program Analysis-Fire Occurrence Database (2020).[95] Both analyses involved regression discontinuity designs and included robustness checks. No statistically significant jump in dwelling fires were observed at DST-Onset or DST-Offset for UK data, whereas a significant jump in incidents was observed at DST-Onset (~30%) for wildfires, including in the individual strata of employment (~32%), leisure (~44%), and arson (~26%) in the US. Only the leisure stratum was significant for DST-Offset (16%). Focussing on wildfires, similar results are observed after taking seasonality and weather into account, but differential effects were observed when specific hours of the day were analysed. DST-Onset appears to both reallocate and increase incidents (reduced in morning and increased in late afternoon/evening) whereas DST-Offset appears to cause the opposite reallocation (net effect is null). DST-Offset is also more sensitive to changes in bandwidth, omitting observations, and using polynomials, but some instability is also observed in tests of states that do not observe DST. Daylight timing and hours of drying before ignitions may play a role. Overall, DST-Onset may play a role in increasing human-caused wildfires in the USA, possibly in part from both cognition and changing the timing of ignitions relative to sun time.

##### *(vii) DST vs no DST*

The previous categories all pertained to effects of transition. The following two studies include comparison of regions that practice DST paradigms with regions that do not.

In the first, Gaski et al. (2011) find that SAT scores (Scholarly Aptitude Test: combined math and reading) are ~16 points worse in DST practicing regions, including after adjustment for longitude with time zone.[100] The authors assessed mean SAT scores for ~ 350 public high schools (score per school) over a 10-year period (1997-2006), in Indiana (USA), weighted by each school's number of test-takers in a weighted least squares linear regression. The schools were based in two different time zones, with one part of one time zone practicing a DST paradigm and the other not. Approximately 20% of scores were obtained in DST practicing regions. Of note, longitude was not associated with DST scores, but Indiana spans ~5° (a time zone spans ~15°). Models included adjustment for SES and ethnicity and with/without longitude (to avoid multi-collinearity). The effects are apparently worse in lower SES groups (means of time zone and SES strata). The main limitation is the non-repeated measures or not using a difference-in-differences approach.

The second study is that by Schaffner et al. (2018), which is already described in the subcategories on attention and perception.[105] The authors find no differences in SIT (attention) scores or lottery (risk perception) scores between neighbouring NSW and Queensland populations on a given day one week pre DST-Onset on NSW (Australia). The study by Schaffner et al. (2018) is too limited (one measurement) to allow a conclusion, whereas the study by Gaski et al. (2011) is more convincing of an effect of living with a DST paradigm, but the findings need to be confirmed in another place and population.[100 105]

##### *(viii) Stock Market Returns as a Proxy for Investor Sentiment and Decision-Making*

The systematic review identified two studies that consider stock market returns (growth or declines) following DST-Onset and/or DST-Offset transitions. Returns are considered as a proxy for investor sentiment and decision-making (i.e., their outlook and assessment of risk – cognitive processes), which can cause prices to increase or decrease. After citation searching, a further 13 relevant articles were identified, seven of which comprise two threads of commentaries, replies, and rebuttals based around the Kamstra et al. (2000) article, which was the first article on the topic and identified in the systematic review.[101 111-116] That only two of 15 studies were identified in the first instance likely stems from strictly not selecting investor sentiment change as a proxy for cognitive change in our search.[102 103] All studies consider mean market returns on the first Monday (or first trading day) post DST-Onset and/or DST-Offset (which shall be referred to as index Mondays). Comparisons of index Monday mean returns are typically made with other Mondays (or first trading days after weekends) or with all other trading days. In the latter case, a Monday covariate is often included in regression analyses to allow comparison of statistics for index Monday vs all other trading days and all Mondays vs all other trading days (as there is a known "weekend effect" in stock market returns, wherein Mondays are typically more negative). This synthesis section will start with the earliest article by Kamstra et al. (2000),[101] synthesize the two threads of commentaries that follow this article, synthesize other articles published in this period (until 2010, which often include reanalyses of the Kamstra et al. (2000) study), and finish with a study published in 2019 that only considered returns based on mergers only.

Kamstra et al. (2000) analysed mean returns on the first trading day post transitions in a joint analysis using OLS regression. Larger negative returns (2 to 5 times lower than average

Monday returns, $p<0.05$) were observed on the first trading day following transition for either equal- or value-weighted indices from NYSE, AMEX, NASDAQ (all USA, 1967-97), S&P500 (USA, 1928-1997), TSE300 (Canada, 1969-98), and UK-Total Market (UK, 1969-98). According to the authors, this implies a financial loss of about 31 billion dollars in the US for every transition – of course, this includes both realised (sold) and unrealised (not sold) returns. The DAX100 index (Germany, 1973-98) was not significant at the 5% level. The results held after utilising a maximum likelihood estimate of a GARCH or a bootstrap estimation to account for autocorrelation and heteroscedasticity. Data was obtained from CRSP (Centre for Research in Security Prices) and Datastream.

In one comment thread, Pinegar (2002) assessed the same NYSE, AMEX, and S&P500 data (with one extra year).[115] The author replicated the Kamstra et al. (2000) findings[101] but then showed they attenuate (including DST-Onset and DST-Offset strata) with heteroscedasticity adjustments (White correction and Glosten correction), Bayesian sample adjustments, and omission of two outlier years (large stock market crashes) with many estimates no longer significant at the 10% level. Kamstra et al. (2002) claim that the Bayesian adjustments and omissions were inappropriate, but the results after heteroscedasticity adjustments were in line with their original findings.[116] In the other comment thread, Berument et al. (2010) studied the same American stock market indices up to 2007 (but did not include the pre-1967 years) and compared the joint index Mondays to all other trading days, but also included a Monday covariate and 15 lag returns.[112] Stock market volatility was also assessed using an exponential (E-)GARCH model. Neither differences in returns nor volatility were significant at the 10% level. Kamstra et al. (2010) respond that Berument et al. (2010) used an over-parameterized model that is prone to bias and that the lags would dilute any transition effects, but agree that transitions not have a significant influence on volatility (albeit unclear to what extent volatility reflects cognitive change).[114] They further state the model did not perform well, based on large differences in E-GARCH and OLS estimates. Kamstra et al. (2010) repeat the Berument et al. (2010) analyses using their own dataset (i.e., without the additional ten years, without the few mistakes they identified in the Berument et al. (2010) data coding, but possibly without any revisions since 1997 that might have been made by their data provider). They found estimates that are more similar across the different models and most return differences were significant at the 10% level. They also produced similar findings using Hansen's generalized method of moments with calculation of heteroscedasticity and autocorrelation consistent standard errors, which is less parameterized. In an unusual response, wherein none of the highlighted issues were addressed, Berument et al. (2011) performed the same analysis as before, but also included an index Monday return*market volatility interaction term and a Monday mean*market interaction term.[111] This inclusion appeared to change the index returns from negative to positive (not statistically significant) and the interaction itself was statistically significant, leading the authors to claim that the transitions change the returns-volatility relationship (i.e., that transitions affect the marketer's cognitive response to a more or less volatile market, even if there is no significant effect on returns). In a final response, Kamstra et al. (2013) highlight the same issues (over-parameterisation, biased estimates), additionally highlight the lack of discussion to interpret why the index returns estimate changed sign, whilst maintaining their original conclusion from Kamstra et al. (2000).[101 113]

Six related articles were published between the first and last paper by Kamstra et al. (2000, 2013). Worthington (2003) finds no consistent effects in the Australian equity market (data drawn from ASX, 1980 to 2003) after using various index dates (given the non-standardised or nonexistent DST practice across Australian territories).[117] The author used similar methods to both Kamstra et al. (2000) and Pinegar (2002), and compared index days to all other days and all other first days of trading after weekends. Lamb et al. (2004) re-analyse the Kamstra et al. (2000) data, agree with Pinegar (2002) about the test sensitivity to two outliers, and provide two additional placebo tests, comparing returns after two public holiday weekends (after which investors may experience sleep loss) and after other weekends in October.[118] First day of trading post-holiday weekends (including 4th July and St. Patrick's Day on the 18th March) returns are not significantly different to other days but October Monday returns are 3x more negative than other days at the 1% level. More confidence can be placed in the latter finding as contrary to a transition effect insofar as the holiday weekends do not include a change in clock time of sun phases but the October negative returns suggest an October dip that coincides with DST-Offset. October index Mondays were only different to non-index October Mondays for equally weighted but not value weighted indices, but this effect vanished after addition of two more years (1998 and 1999). Dowling & Lucey provide two analyses, first of the Irish market in their 2005 publication and then of several international markets in their 2007 publication.[119] [120] The Irish analyses used value weighted data from ISEQ (1988-2000) subtracted from the daily return on FTSE All world index to isolate a local component, and used OLS regression with White standard errors in addition to further least absolute deviation (LAD) and trimmed least squares (TLS) regression models with both all other days or all other Mondays as comparisons.[120] Compared to all other Mondays, estimates were significantly negative at the 10% level after adjustment for weather-related variables (deemed to possibly affect mood). A limitation is that DST-Offset index Mondays are often a public holiday in Ireland and the authors did not supplement with Tuesdays as first-trading day in this event. For the international analyses, they used 37 country indices (see article for full market listing) from Datastream from 1994-2004 and used GARCH models after specification based on Log Likelihood Ratio tests.[119] They included a World Index to adjust for global economic changes and a Monday variable to account for the Monday effect. Few countries presented with significant index Monday differences (at least at the 10% level) and those that indicated mostly positive returns. Regarding countries mentioned in other studies, no significant differences in returns were observed for the USA, UK, and Germany, but a significant effect for Ireland (positive for main index and negative for small capitalisation indices), Australia (negative for main index) and for Canada (positive for small capitalisation index).[119] Gregory-Allen et al. (2008) also analysed multiple market data using White standard errors.[121] They analysed markets from 22 countries (of the MSCI World Index with data from Global Financial Data as opposed to DataStream (see article for full market listing), from 1966-2005 – depending on country and DST paradigm in use). The authors found only a significant negative joint difference for the UK and negative DST-Offset difference for Luxembourg at the 5% level when comparing index Mondays against all trading days. Gerlach (2010) runs a series of placebo and control tests using the Kamstra et al. (2000) data and methods, and find, *inter alia*, significantly worse or no difference in returns for non-index Mondays in October compared to index Mondays in October.[102] Using data from other markets without a DST-Onset or DST-Offset date overlap with the USA but using USA DST-Onset/DST-Offset dates, few significant market differences are observed. Few significant differences using UK DST-Onset/DST-Offset dates or Germany DST-Onset/DST-Offset dates in non-UK or non-Germany markets are also observed. Using a regression model with a Monday covariate, the author also finds significant negative differences for DST-Offset index Mondays and other October Mondays (often greater in magnitude than for index Monday DST-Offset). Müller et al. (2009) analysed the German market (DAX30, MDAX, and REX General Bond Indices, 1980-2007) in addition to other central and Northern European markets comparing index Monday mean returns with other Mondays and other trading days using OLS and GARCH models.[122] Only the Rex General Bond regression was significant at the 10% level for DST-Offset index Mondays. In a stratified analysis using five-year intervals, they find an index Monday difference for the first five years only and no index Monday differences in Northern, Central, or Southern European country markets.

Overall, only the Irish market data by Dowling and Lucy (2005) supports a transition effect on returns in addition to Kamstra et al. (2000).[101 120] All other studies indicate a null effect with placebo tests suggesting the Kamstra et al. (2000) conclusion does not account for a general October dip in returns and outlier year omission and additional year inclusion tests highlighting an estimate sensitivity.

Next is a 10-year jump to publication of the most-recent article identified, which involves assessment of abnormal stock market returns of target US firms as part of completed public-to-public mergers and acquisitions announcements (data from SDC Thompson OneBanker, 1977-2017).[103] This focus was due to target firms exhibiting pronounced stock performances following merger announcements. Abnormal stock returns were defined as being in excess of corresponding stock market returns on the same day, with the top and bottom 1% winsorized. The authors find that index day deals experience stronger abnormal returns compared to other firms at the 1% level using OLS regressions with Petersen's clustered standard errors on each day of the weeks were estimated. Inclusion criteria were target size being at least 1% market value of the bidders, at least 50% acquisition, no minority stake purchases, acquisitions of remaining interest, spinoffs, recapitalisations, or repurchases. The timing of data collection was from 1 day before announcement (weekend announcements (< 2% total) were grouped with Mondays). Covariates in analyses included rumours, target price run-up (cumulative abnormal stock returns in the 30 days before merger), market capitalization, price, number of analysts following merger, and presence of block-holders activity. They also included merger deals with more than one bidder, tender/stock/cash deals, hostile mergers, pooling of interest, and when target and bidders do not share the first two SIC industry digits, and three-day lagged abnormal target stock returns. The authors include a series of robustness tests including testing against other Mondays only, testing a subset of only 100% merger acquisitions, testing a non-winsorized sample, and testing a subset of only positive abnormal trading volumes (in excess of the 100 to 31 days prior). They also tested with a firm-matching covariate, a GARCH model, and ran a placebo test of 106 countries that do not follow a DST paradigm. All robustness tests were significant apart from the latter placebo test.

Overall, there appears to be no effect of transitions on stock returns, with the few instances of an observed effect possibly being a statistical artefact and/or coincident with a general October dip. It remains to be seen if the merger study can also be explained by an October dip.[103] In conclusion, the evidence is in favour of a null effect on cognitive behaviour/performance that can be evidenced in stock market returns.

**Cognitive-Table 2: JBI Quality Indicators**

(Questions and possible answers are shown in detail in Table footnote)

| Author (Year) | Q1 | Q2 | Q3 | Q4 | Q5 | Q6 | Rating | Notes |
|---|---|---|---|---|---|---|---|---|
| Barnes (2015)[110] | Y | N/A | N+ | Y | U* | N | L | +Study includes states that did not always or currently practice DST, * no robustness tests used. |
| Ben Simon (2022)[98] | Y | N/A | Y/N+ | Y | Y | N | M/H | +Study includes a state that did not always practice DST. |
| Cho (2017)[97] | Y | Y | Y | Y | Y | N | H | |
| Doleac (2015)[94] | Y | Y | Y | Y | Y | N | H | |
| Gaski (2011)[100] | Y | U’ | U+ | Y | Y | N | M | '+School level data was used. |
| Goodwin (2023)[12] | Y | Y | Y | Y | Y | N | H | |
| Herber (2017)[108] | Y | U’ | U+ | Y | U* | N | L | 'Difficult to say across countries, some countries may be more prone to holding the tests pre DST-Onset and others to post DST-Onset, # unknown, * no robustness tests used. |
| Hicks (1980)[107] | Y | N/A | U+ | Y | U* | N | L | + cannot be determined, * no robustness checks, n=1 study. |
| Kountouris (2021)[96] | Y | N/A | Y | Y | Y | N | H | |
| Kountouris (2020)[95] | Y | N/A | Y | Y | Y | N | H | |
| Lindenberger (2019)[15] | U~ | Y | U+ | Y | U* | N | L | ~Possible unclear time of death, could be important with low sample size, + misclassifications cannot be ruled out. * there were limits as to what could be done, no robustness tests used. |
| Maier (2021)[104] | Y | Y | Y | U# | U* | N | L | #Potential issue with timing of testing and concurrent measurements, * no robustness checks. |
| Monk (1980)[109] | Y | Y | Y | U# | U* | N | L | #Self-reported abilities with no observer, * no robustness tests. |
| Orsini (2022)[93] | Y | Y | Y | Y | Y | N | M/H | |
| Schaffner (2018)[105] | Y | Y | Y | U# | U* | N | L | #Issue with timing of testing and concurrent measurements, * no robustness checks. |
| Umbach (2017)[99] | U~ | Y | Y | Y | Y | N | M | ~Only post transition times were used as controls. |
| Wagner (2012)[106] | Y | Y | N/A | U# | U* | N | L | #Not necessarily at work, * no robustness checks. |

**JBI Critical Appraisal Checklist for Quasi-Experimental Studies (Adapted)**

Answers can be either Y=Yes, N=No, U=Unsure, or N/A.

1. Is it evident which factor is considered the 'cause' and which one is the 'effect'? Is there no ambiguity regarding the sequence of variables under investigation?
2. Were the characteristics of the study participants involved in comparisons adequately matched?
3. Were the exposures/treatments (beyond that of interest in our study) of the study participants involved in comparisons adequately matched?
4. Were the methods used to measure the outcomes the same for participants who were part of different comparisons?
5. Were appropriate statistical methods employed to analyse the data?
6. Are there any irregularities in the data reported?

#### 7. All-Cause Mortality, Neurologic & Gastrointestinal Section Synthesis

Thomas C. Erren & Ursula Wild

##### *Introduction*

This section includes endpoints for which systematic screening returned too few studies to warrant their own sections; namely, (i) one study of a neurological-related endpoint, (ii) two studies of gastro-intestinal (GI)-related endpoints, and (iii) three studies of all-cause mortality; thus, six studies in total for this section. This synthesis is laid out as per (i)-(iii) above, with an additional overall Conclusion sub-section. For an overview of the included studies see Mortality-Table 1. JBI quality indicators are presented in Mortality-Table 2.

Briefly, the studies in this section were published comparatively recently; the oldest in 2019 and the most recent in 2022; albeit, two studies on mortality include data from as early as 1980 in Austria (Poteser et al. 2020) and as early as 1998 from 16 countries across Europe (Levy et al. 2022).[123-125] One neurological-endpoint study by Schneider et al. (2019) concerned counts of seizures in the USA.[126] The two gastro-intestinal-related endpoints were conducted by Föh et al. (2019) and Zhang et al. (2020) on sick days due to ulcerative colitis and Crohn's disease in Germany and on various GI-disorders in the USA/Sweden, respectively.[6 127] The three mortality studies were by Poteser et al. (2013) in Austria,[125] Levy et al. (2022) in 16 countries across Europe,[124] and by Cook et al. (2022) in the USA.[123]

Four of the six studies consider the effects of DST-Onset and DST-Offset.[6 124-126] The other two studies consider the effects of living with DST compared to Standard Time during the summer months; namely, the studies of sick days by Föh et al. (2019) and mortality by Cook et al. (2022).[123 127] In terms of quality rating (see Mortality-Table 2), we find one study of low quality (Föh et al. 2019), one study of high quality (Cook et al. 2022) and the other four studies of medium quality.[6 123-127]

##### *(i) Neurological-related endpoints*

Schneider et al. (2019) studied seizures in relation to transitions using data from a large cohort study in the USA (2008-2016) using three methods.[126] First, in absolute terms, a decrease in seizures was observed in the first week post DST-Onset compared to all other weeks. Second, using a scaled seizure count for each individual (in an attempt to statistically balance the impact of individuals who may have a much higher baseline seizure propensity than others and differences in reporting seizures and enrolment timing), a 6% lower relative rate of seizures was observed in the first week post DST-Onset compared to the week pre DST-Onset. There was no difference compared to the second week post DST-Onset, but an increased rate was observed compared to all other weeks. This latter finding is in contrast to that from the first method. No differences were observed across DST-Offset. Third, using a generalized linear mixed model approach to compare weekdays pre vs post transitions, no differences were observed. Despite one method resulting in a statistically significant decrease in seizure propensity in the week following DST-Onset, the authors themselves conclude no clear associations between transitions and seizure propensity. The possible timing and reporting bias (which led to their second method above) disallow conclusion.

##### (ii) GI-related endpoints

Föh et el. (2019) reports increased frequencies of sick days due to ulcerative colitis and Crohn's disease in the 30 days following DST-Onset compared to the 30 days pre DST-Onset in a large cohort of more than 10 million insurance holders of a German health insurance company in 2010-2013.[127] There was no significant difference between pre and post DST-Onset. However, the use of 30-day frequency measures analysed by one-way ANOVA means any conclusion is open to confounding by season-associated variables. Even if true, we do not know if there is clustering in the more immediate post transition time or whether the increase is stable over the 30 days (i.e., whether this an 'acute' effect of transition or a more 'chronic' effect of Standard Time). These issues lead to assigning the lowest quality rating to this study.

The study by Zhang et al. (2020) assessed the effects of transitions on, *inter alia*, various GI-related endpoints, including disorders of the appendix, liver/gallbladder, biliary tract/pancreas, stomach, and intestines.[6] They used electronic health records of more than 150 million patients in the USA and 9 million in Sweden and compared mean diagnosis rates on days in the week post transition with what would be expected based on the second week pre transition and second week post transition. They used both Bayesian and Frequentist methods with stratifications by sex, age, and in/out-patient where possible. For US inpatients post DST-Onset, higher rates of non-infective enteritis and colitis for males between 0 and 10 years (RR = 1.06, CI 1.00-1.13) and females over 60 years (RR = 1.03, CI 1.00-1.05) were observed. A higher rate of appendix disorders was also observed for males over 60 years (RR = 1.08, CI 1.00-1.18). Following DST-Offset, lower rates of liver diseases in females between 21 and 40 years (RR = 0.91, CI 0.84-0.97) and over 60 years (RR = 0.94, CI 0.90-0.98) and of gallbladder and pancreas diseases in females between 21-40 years (RR = 0.94, CI 0.91-0.97) were observed. Coding errors in the electronic health records and exclusion of data from uninsured patients may be sources of bias, but coding errors are estimated to be negligible in this dataset. Moreover, a "driver and passenger" effect – meaning that some of diseases may trigger others – cannot be ruled out. Chronic diseases may only be diagnosed after reaching a certain severity or an exacerbation; thus, they may have been present pre transition. Of course, the transition may be the stressor that triggers exacerbations.

##### (iii) All-cause mortality

Of the three studies included here, that by Poteser et al. (2020) is the smallest by geography (Vienna, Austria) but the largest by number of included transition events (1980-2018).[125] They compared the Sunday, Monday, and Tuesday-to-Friday pre and post DST-Onset and DST-Offset and found that mortality increased by about 3% for Tuesday-to-Friday following DST-Onset but not DST-Offset. There was no information about differences by sex and age, which would have been useful to allow for better comparison with the other studies of all-cause mortality. The one hour extra or one hour less on the Sundays of time transition may confound but this only applies to the Sundays while similar findings were observed comparing other weekdays.

Lévy et al. (2022) conclude that mortality decreases after the first two weeks post DST-Onset and increases post DST-Offset in Europe,[124] which contrasts with the study by Poteser et al. (2020).[125] For Austria specifically, Lévy et al. report no difference. The authors attempt to analyse complex data collected across 16 European countries over more than a decade, which is demanding as differences too small to be relevant can become statistically significant with such large numbers, and the results will vary depending on model structures.[124 128]

The study by Cook (2022) – which received the high quality rating – considers mortality in 92 counties in the state of Indiana in the USA from 2003-2008.[123] Cook investigated the medium- and long-term mortality effects associated with living with DST compared to Standard Time during summer months in detail. Risk reduction effects for DST compared to Standard Time (4.03-5.07%) were observed, especially among those aged 65+ years, and primarily in regard to cancer mortality. Methodologically, the author compared counties that implement a DST paradigm to those that do not, and conducted sensitivity analyses that include one-month latency periods and excluding the month following DST-Onset. Further robustness checks included whether comparable results could be obtained for counties that are specifically located along the border of DST-practicing counties (but do not practice DST) compared to DST-practicing counties and include temporal placebo exposures. The results were supported in every instance. A one-month latency test (i.e., excluding the month in which DST-Onset occurs) revealed slightly greater risk reductions (4.1-5.5%) compared to analyses with DST-Onset month included. Although effects of transitions were not reported, the difference with including latency is compatible with a potential detrimental effect of transition on mortality. The author concluded that the DST-associated extra hour of evening daylight may contribute to lower mortality patterns and pointed out that a solar vitamin D synthesis mechanism is worthy to be explored as a possible explanation.

Taken together, there are contrasting findings regarding transition-associated effects on mortality at DST-Onset. Reduced mortality when DST is in practice during the summer months compared to no DST was observed in one high quality study.

##### *Conclusion*

Possible effects on seizures and medical leave due to GI-disorders are inconclusive. There may be short term increases and decreases in different GI-associated disorders following transitions. There may also be increased mortality following DST-Onset in the short term. This contrasts with, but is not contrary to, observation of reduced mortality overall during summer with DST compared to no DST.

**Mortality-Table 1: Overview of studies in "Other" Section, grouped by subcategorizations used in synthesis.**

| Author (Year) | Population/Data | Exposure/Comparator | Statistics | Results | Rating |
|---|---|---|---|---|---|
| ***Neurological*** | | | | | |
| Schneider (2019)[126] | n=14,166 (51% female), 19.46 ± 16.13 years, USA | 1 week pre vs 1 week post transitions, 2008-16 | (i) Absolute comparisons, (ii) standardized relative rates, (iii) linear mixed models (see text for details) | ↑ Relative rate of seizures following DST-Onset by method (ii) only. No differences for DST-Offset. | M |
| ***GI-related disorders*** | | | | | |
| Zhang (2020)[6] | USA MarketScan dataset (n>150 million records), Swedish national inpatient register (n>9 million records) | 1 week post vs 2 weeks pre and 2nd week post transitions, 2003-14 (USA) & 1980-2011 (Sweden) | Bayesian and frequentist relative risks, including use of "sham"/negative controls | ↑ diagnoses of appendix and non-infective enteritis/colitis disorders following DST-Onset. ↑ diagnoses of appendix and ↓ diagnoses of liver/gallbladder and pancreas disorders following DST-Offset.* | M |
| Föh (2019)[127] | Data from > 10 million health insurance records, Germany, average of 56.4 deaths/d | 30 days pre vs 30 days post transitions, 2010-13 | One-Way ANOVA with Tukey's honest significant difference (HST) test | ↑ medical leave following DST-Offset but not DST-Onset | L |
| ***All-Cause Mortality*** | | | | | |
| Cook (2022)[123] | n=6107 deaths (2907 female) Indiana-USA | DST vs non-DST practicing counties/times, 2003-2008 | Poisson regression with placebo and sensitivity analyses. | ↓ 4-5.1% mortality from living with DST vs Standard Time | H |
| Lévy (2022)[124] | n=59,067,376 deaths, 16 European countries | 2 months pre vs 2 months post transitions, 1998-2012 | Multiple negative binominal regression models, results presented as incidence rate ratios with a 95% CI | ↓ 3.5% and 2.8% mortality for week 1; 2.8% mortality and week 2 following DST-Onset; ↑ 1.8% and 2.3% mortality for week 1 and week 2 following DST-Offset. | M |
| Poteser (2020)[125] | Daily total mortality rates from Statistics Institute, Vienna-Austria | 1 week pre vs 1 week post transitions, 1980-2018 | Poisson regression; sine-cosine function modelling | ↑ 3% mortality following DST-Onset (Tue-Fri). ↑ 4.9% mortality on Sunday and ↓ 3.9% on Monday following DST-Offset. | M |

L=low, M=medium, H=high. *too many non-significant differences to allow coherent tabulated overview; see text for specifics.

**Mortality-Table 2: JBI Quality Indicators of Studies**

(Questions and possible answers are shown in detail in Table footnote)

| Author (Year) | Q1 | Q2 | Q3 | Q4 | Q5 | Q6 | Rating | Notes |
|---|---|---|---|---|---|---|---|---|
| Cook (2022)[123] | Y | Y | Y | Y | U/Y | N | H | |
| Föh (2019)[127] | Y | U | U | Y | Y* | N | L | *30-day frequency measures analysed by one-way ANOVA means any conclusion is open to confounding by season-associated variables and we cannot account for possible clustering around the transition. |
| Lévy (2022)[124] | Y | Y | Y | Y | Y | N | M | |
| Poteser (2020)[125] | Y | U | U | Y | Y | N | M | |
| Schneider (2019)[126] | Y | U | U | Y | Y | N | M | |
| Zhang (2020)[6] | Y | U | U | U/Y | Y | N | M | |

**JBI Critical Appraisal Checklist for Quasi-Experimental Studies (Adapted)**

Answers can be either Y=Yes, N=No, U=Unsure, or N/A.

1. Is it evident which factor is considered the 'cause' and which one is the 'effect'? Is there no ambiguity regarding the sequence of variables under investigation?
2. Were the characteristics of the study participants involved in comparisons adequately matched?
3. Were the exposures/treatments (beyond that of interest in our study) of the study participants involved in comparisons adequately matched?
4. Were the methods used to measure the outcomes the same for participants who were part of different comparisons?
5. Were appropriate statistical methods employed to analyse the data?
6. Are there any irregularities in the data reported?

### 8. Update to June 2025

Philip Lewis & Jonas P. Wallraff

##### *Introduction*

Systematic screening identified 30 relevant articles published since our initial search to September 2024 to be included as an update. The articles for the update are categorized into subsections below (with some articles in multiple subsections). The subsections here correspond with Appendix Sections 1-7 (i.e., the subsection "Cardiovascular" below corresponds with "Cardiovascular Section" of the whole Appendix). An overview of the studies can be found in Update-Table 1, with individual JBI quality assessments presented in Update-Table 2. Briefly, eleven studies were rated as high quality, nine as medium quality, and ten as low quality. Key factors influencing ratings included robustness tests, reference period issues, and sample sizes. We provide a detailed synthesis of the studies in the same order as Appendix Sections 1-7. The conclusion thereafter also links to the conclusions in Appendix Sections 1-7.

##### *Cardiovascular*

Four studies are included here. Two of these studies report no clear differences in measures of vascular stiffness, blood pressure, or augmentation index following both transitions in Canada,[129] or on myocardial infarction and unstable angina incidence and periprocedural mortality during their interventions within the first three or first seven days following transitions compared to reference periods in Poland.[130] The studies in Canada and Poland were rated low quality. One study describes mild-to-moderate probabilities of low effect size increases in cardiovascular events following DST-Onset and negligible probability of change following DST-Offset in the USA.[131] This USA study is rated medium quality. The fourth study reports increased AMI for two weeks following DST-Onset and no changes following DST-Offset compared to reference periods in the USA and is rated high quality.[132]

More specifically regarding the two low quality studies, Al-Bakry et al. (2024) included a very small sample size and their reference-index period comparison is not meaningful in their study in Canada.[129] They considered 11 and ten healthy young adults on the Sunday previous to the Monday, Wednesday and Friday post DST-Onset in 2023 and DST-Offset in 2022, respectively. It is also difficult to interpret this finding as relevant regarding findings on acute MI given the healthy status of the sample. Kazirod-Wolski et al. (2023) compared MI and unstable angina from 874,031 patients in the Polish National Register of Interventional Cardiology Procedures (2014-2021) to what they define as "ordinary" incidence, but to what this refers is ambiguous.[130] Moreover, there are discrepancies between graphs, tables and text. There is also ambiguity regarding whether "Sunday after transition" refers to the transition day itself or to the 7$^{th}$ day post transition.

Satterfield et al. (2024) show, in their medium quality-rated study, mild-to-moderate probabilities of low effect size increases in cardiovascular events (including AMI, stroke, cardiogenic shock, cardiac arrest, and sudden death) in the first week post DST-Onset compared to the two weeks pre and the second and third weeks post.[131] They observe a negligible probability of change following DST-Offset in the USA in 2015-2019. The authors used a Bayesian approach with "weakly informative" priors so that inference is predominantly data driven. As the population studied was limited to those with appropriate medical insurance in the USA, we assign a medium quality rating. The findings could be biased by not including potentially more susceptible individuals who cannot obtain sufficient medical insurance.

Tanaka et al. (2025) identify in their high quality study a significant increase (~27%) in AMI following DST-Onset but not DST-Offset (2002-2012) using hospital discharge records in Indiana (USA).[132] This analysis does not account for in-hospital fatalities after AMI but there were no differences in AMI mortality overall. A regression discontinuity design was used including four weeks pre and four weeks post transitions, including adjustment for daily weather and temperature conditions. Stronger effects were observed in women compared to men and in over 50 years of age compared to under. In terms of time, the effect is highest in the first week and has diminished to prior levels by week 3. The findings were robust to using all days compared to only weekdays, different bandwidths, different polynomials, and difference-in-differences approaches using counties in the state that did not observe a DST transition in certain years. The also conducted a longer-term analysis comparing years with transitions to years without. A non-significantly higher count of AMIs was observed in the years with transitions. Of course, as this includes transitions. it is not a straightforward comparison of DST compared to Standard Time.

Overall, although the study by Satterfield et al. (2024) detracts from indications that DST-Onset is associated with increases in myocardial infacrtion,[131] these studies together do not change the conclusion regarding transitions and cardiovascular outcomes put forward in the Cardiovascular Section. Indeed, the study by Tanaka et al. (2024) adds further strength to the finding of increased AMI following the DST-Onset transition.[132]

##### *Psychiatry*

Three studies considered psychiatric outcomes; namely, for transition effects on depressive symptoms in Chile,[133] for effects of living at a different longitude in a given time zone on suicides in the USA,[134] and for transition effects on suicides in Austria, Switzerland, and Sweden.[135] The Chile and USA studies are rated low quality and the European study is rated high quality. Labarca et al. (2024) found no difference in depressive symptoms using the Beck Depression Inventory following DST-Onset or DST-Offset in 30 male healthy university students in Los Angeles, Chile.[133] It is unclear exactly when or how often questionnaire data was collected during the two weeks pre and post transitions study periods; thus, the study is rated low quality. Reis et al. (2023) report increased suicides in more western longitudes compared to more eastern longitudes in specific time zones in the USA.[134] They used county level population and suicide data from the CDC, but the study is limited by ecological design and by only partly consistent sensitivity analyses; thus, it is rated low quality. These studies do not change conclusions regarding DST and psychiatric outcomes, but the latter might be considered hypothesis generating in terms of observed potential differences by latitude. Plöderl et al. (2024) found no differences in suicides following transitions using national data from Austria, Switzerland, and Sweden for the years 1981-2022 and Poisson regression and Bayesian changepoint analyses.[135] The authors performed sensitivity and placebo analyses using years wherein Easter holidays did not occur during index or reference periods, assessing years without DST (pre-1980), starting assessment from the Monday post transitions, comparing Sunday-Tuesday or the 7 days or the 3 weeks pre and post transitions. There were no apparent differences by age or by country (latitude). The study is rated high quality.

Overall, the study by Plöderl et al. (2024) in particular is supportive of no transition effects on suicide.

##### *Traffic Accidents*

Three studies update the Traffic Accidents section with two rated high quality and one rated medium quality.[136]

In the high quality rated study from Greece considering the years 2006-2016, Laliotis et al. (2023) report a decrease in serious vehicle accidents alongside a slight increase in fatal accidents in the two weeks post DST-Onset compared to the two weeks pre, while the number of minor accidents and total (all severity) accidents remained unchanged. Following DST-Offset they report an increase in total accidents that appears attributable to an increase in minor accidents with no clear change in fatal or serious accidents. For both transitions, these changes were linked to evening hours and the transition effects on ambient light. No strong differences were observed in other time windows. The findings are mostly supported by various robustness checks, including tests with different bandwidths and placebo testing, except for stratification by periods before (pre May 2010) and during (post May 2010) economic crisis and austerity measures in Greece. The overall finding appears driven by the pre-austerity time period.

In the high quality rated study by Gilmore (2024) in Chile, no immediate significant impact of either DST-Onset or -Offset transitions on total accident rates are observed using data from the National Commission of Transit Security in a Regression Discontinuity Design.[137] These null effects hold across accident severities (minor, serious, fatal) and are robust to different bandwidth specifications. The author then applied a difference-in-differences analytical approach, which included transition and weeks of DST vs no transition and weeks of Standard Time (and vice versa for DST-Offset and Onset) – possible, due to differences in transition dates over the years in Chile. Two months of DST including the DST-Onset transition was associated with decreased accidents (~2.7%) compared to two months of Standard time without a transition during the same months of the year. The opposite (~1.1% increase, albeit not statistically significant) was observed for DST-Offset. When dividing into acute post-transition times (1-2 weeks) and remainder times, the effects were significant for the remainder times in both instances. No trends were observed for pre-transition times. Next the author assesses the times-of-day when light has changed (morning and evening, i.e., ±3 hours around sunrise and sunset) and when it hasn't (mid-day), accounting for differences in times of light change across the various latitudes of Chile. Mid-day hour accidents were increased by 3.9-6.1% immediately after DST-Onset and decreased by 2.8-4% after DST-Offset. Conversely, in the morning, a 7.5% decline in accidents is observed after DST-Onset and 5.4% increase after DST-Offset. No significant differences were detected in evening hours (±3 hours around sunset) for total accidents. When focusing exclusively on serious and fatal accidents, a reduction is observed post DST-Onset and an increase is observed post DST-Offset for evening accidents whilst morning hour accidents were no longer significantly different.

In the medium quality rated study by Woods et al. (2025) using data from FARS (Fatality Analysis Reporting System) in the USA (only DST-practicing states from 2010-2019, weekly fatal crashes were observed to increase by 3.3% following DST-Onset and decreased by 2.4% after DST-Offset.[138] The authors used a Posisson regression model and included the five weeks pre and post transitions. When focusing on different time windows during the day, darker conditions (post DST-Onset mornings and post DST-Offset afternoons) were associated with increased fatal crashes and lighter conditions (post DST-Offset mornings and post DST-Onset afternoons) were associated with decreased fatal crashes. Stronger adverse effects of light timing changes were observed for pedestrians and bicyclists. The medium quality rating was assigned because of no placebo tests.

These studies do not change conclusions regarding DST and traffic accident outcomes, rather they add to the argument regarding USA and non-USA differences, and to the conclusion regarding a role for change in daylight availability at specific times affecting accident risk directly. The study by Laliotis et al. (2023) also opens the door to other confounding factors. Added by the study from Gilmore et al. (2024) is that DST during summer months may reduce traffic accidents overall (at least in the initial months post DST-Onset). The opposite is observed for the post DST-Offset period, though not as strong.

##### *Non-traffic Accidents, Injuries, & All-Cause Hospital Admissions*

Two studies on work accidents are rated as high and low quality from Italy and Turkey, respectively,[139 140] and one study on medical malpractice severity in the USA is rated low quality.[141] In the high quality study from Italy, Depalo et al. (2023) find decreased work injuries and disabilities in the 5 days following DST-Offset but no clear effect following DST-Onset.[139] They used a regression discontinuity design analysis of data from the Italian National Institute of Insurance against Accidents at Work for the years 2013-2017. The effects are more apparent for older compared to younger workers and in the north of Italy compared to in the south. The author also applies a "long run" assessment after the 'acute' effects have returned to normal at one week by assessing a second week and conducting placebo tests using different times of year. No apparent "long run" effects are observed post transitions. The effects sizes are small but in favour of maintaining transitions.

In Turkey, Kocali et al. (2023) report that cancellation of transitions would prevent some work accidents, but there is a lot of ambiguity regarding the methods and results.[140] For instance, prevention at a rate of "935.1116%" is not comprehendible. Poisson regression is reported to have been used but the article contains no coefficients from Poisson models. There is a formula for what appears to be a log-linear regression that may have been used, but there is ambiguity regarding the variables split into morning and night and regarding the indices corresponding to the sum symbols. The study is rated low quality and cannot be considered when drawing conclusions.

Gao et al. (2024) report no differences in severity of medical malpractice claims (severity based on payment amounts) in the week post DST-Onset in USA for the years 1990-2018 using data from the National Practitioner Data Bank of the US.[141] Specifically, they include severity levels for total year, week pre DST-Onset, and week post DST Onset for states that use DST and states that do not in an ANOVA model. There is no indication for taking paired measurements into account (i.e., by year). The authors also compared DST months to Standard Time months in control and DST states using a similar ANOVA model and observe a significant interaction. They do not report control and DST state differences, which appear different for Standard Time months, which suggests that perhaps they should not be compared given potential residual confounding and potential regression to the mean. The study is rated low quality.

Overall, DST-Offset may be associated with decreased work accidents as observed from the one high quality study from Italy.

##### *Sleep & Circadian*

Ten studies consider sleep and circadian biology. Seven are rated as low quality and three rated as medium quality. The medium quality studies are synthesised first, followed by the studies rated low quality.

The first medium quality study by McHill et al. (2024) concerns only sleep midpoint timing.[142] Specifically, the authors found that participants on lower fat diets appear to shift their sleep midpoint time from pre to post DST-Onset faster than participants on a high fat diet. The study included 37 undergraduate students, of whom seven were grouped as high fat diet, ten as intermediate fat diet, and 20 as low-fat diet. The faster shift is described as faster entrainment to the new light-dark and social clock paradigm rather being a less robustly entrained rhythm. It is unclear whether sleep duration changed. The study had a low sample size but used repeated measures and added results that are compatible with their posited mechanism. We should consider the study as hypothesis generating and that some individuals may be more predisposed to potential effects of transitions compared to others. The second medium quality study by Costa-Font et al. (2024) report decreased satisfaction with sleep following DST-Onset in Germany for the years 2008-2018.[143] The authors used regression discontinuity with difference-in-differences (using last Sundays of surrounding months for placebo DST-Onset days) to assess sleep satisfaction based on Likert-scale answers to a question asking about sleep satisfaction on the day of questioning. The study sample were respondents to the German Socio-Economic Panel (age 15+ years and living in Germany). The study sample was large and methods robust, but this was not a repeated measures study so participants who answered pre and post and on different days pre and post were different; thus, the study is rated medium quality. The third medium quality study by Völker et al. (2023) reports lower sleep quality and shorter sleep duration (~15mins) on the Monday following DST-Onset compared to the previous Monday in the year 2019 in a sample of 155 employees in Berlin.[144] There was no apparent effect of chronotype although they were not strictly statistically compared. Sleep and chronotype were assessed by questionnaire. The analyses did not utilise repeated measures although some participants provided data for both Mondays. DST-Offset was not assessed.

Regarding the low quality studies, two studies and their limitations are described above already.[129 133] Specifically, Al-Bakry et al. (2024) found no clear effects on sleepiness, fatigue, sleep latency, duration, quality, or efficiency.[129] The study and its limitations are described in the 'Cardiovascular' subsection above. Labarca et al. (2024) found mostly no differences in sleep quality or sleepiness using various commonly used questionnaires (e.g., PSQI, SATED-Q, ESS, and ISI).[133] The only difference among eight tests was an increased ISI score following DST-Offset. Comparing 7-day wrist actigraphy results of the second weeks pre and post transitions, they found decreased time-in-bed (~65mins), total sleep duration (~56mins), sleep efficiency, and fewer awakenings following DST-Offset and no changes following DST-Onset. Among five rest activity parameters, only relative amplitude was decreased following DST-Onset.

The studies by Ferguson et al. (2024), Zolfaghari et al. (2023), Halfmann et al. (2025), de Lange et al. (2024), and Angelino et al. (2024) make up the other low quality studies.[145-149]

Ferguson et al. (2024) report a decrease in sleep duration of 7 min/day and an increase in sedentary duration of 8 min/day for the week post DST-Offset (n=333 participants) compared to the pre transition week.[145] No changes were observed for DST-Onset (n=291 participants). They assessed movement behaviour (sleep, sedentary behaviour, light-, and moderate to vigorous activity) of parents in Adelaide, South Australia using Fitbit activity tracker data (available from the Annual Rhythms In Adults' lifestyle and health [ARIA] prospective cohort study). It is unclear how capable the device might be able to distinguish between 7-8 mins of sleep vs sedentary activity, especially considering the averaging of non-wear time to a full day and that inclusion criteria included minimum 2 valid days where a valid day has at least 18 h of movement data and a period of sleep. Furthermore, it is unclear what days individual participants contributed to analyses, i.e., whether there are repeated measures in individuals pre and post transitions. Based on this, the study is rated low quality.

Zolfaghari et al. (2023) finds 9 mins shorter sleep duration in 45 to 85-year-old adults in the Canadian Longitudinal Study of Aging 2011-2015 in the week post DST-Onset compared to the week pre DST-Onset.[146] Higher odds of sleep dissatisfaction, sleep-onset insomnia, sleep maintenance insomnia, and hypersomnolence with adequate sleep were found following DST-Offset. No symptoms persisted into a second week. That different groups were assessed pre and post transitions (i.e., non-repeated measures) with some important differences in characteristics that could affect sleep, and that sleep questions and scores concerned the previous month, the study was rated as low quality.

Halfmann et al. (2025) recruited 496 participants on the Monday post DST-Onset and a different 490 participants on the Monday two weeks later in a first test of sleep.[147] The year is not provided. T-tests were used to assess between group differences. Participants were asked how sleepy/tired they felt immediately before testing (first tests at 06:00 and all tests finished by 08:30; sleepiness score on a 10-point Likert scale). They were also asked when they fell asleep the night before and when they woke that morning to determine sleep duration. There were no differences by sleep duration or by sleepiness. Sleepiness was significantly higher on the Monday post DST-Onset after 21 participants were excluded because of failing to answer attention check items and 16 participants were excluded because of incorrect sleep timing answers that disallowed calculation of sleep duration. Such exclusion might have been expected to remove any differences between groups. The second (repeat) study of a different 496 participants on the Monday post DST-Onset vs 490 participants on the Monday 2 weeks later (no year given; counts are after exclusions; participants are restricted to workers only after the observation that almost 25% of first test participants were either unemployed, retired, or students) found no differences in sleep duration, tiredness, sleep debt, or sleep quality, but sleepiness was significantly higher on the Monday post DST-Onset. One-tailed tests were used for the second study. Overall, no clear differences in sleep parameters are observed. The main limitation is the lack of repeated measures.

De Lange et al. (2024) compares accelerometer-derived mean daily sleep duration (midnight to midnight) in n=11,780 participants from the UK Biobank, aged 43 to 78 years on Sundays immediately before and after DST-Offset/Onset transitions to the respective transition days (adjusted, so that transition Sundays equal 24 hours), as well as comparing weekdays before and after transitions.[148] For the DST-Onset transition, data were available from 2014 and 2015, while DST-Offset transitions were covered using data from 2013, 2014, and 2015. There were no repeated measured (between participant comparisons only). Mean daily sleep duration on DST-Onset transition Sundays was ~60 mins shorter than on the Sundays before and after transition. On DST-Offset transition Sundays, sleep duration was ~35 mins longer than on the Sundays before and after. Mean differences in sleep duration between weekdays pre and post transitions ranged from 0.95 to 19.01 mins, but were only significant for Wednesdays and Fridays across the DST-Onset transition.

Lastly, Angelino et al. (2024) identified no significant changes in any sleep-related metric (including overall quality, duration, and PSQI) following either transition (DST-Offset in 2022 or DST-Onset in 2023) n=62 diabetes patients (median age=26) in Naples, Italy.[149] Although paired analyses were used (Wilcoxon signed-rank test), it is unclear who was assessed and when exactly (e.g., were days comparable?). Due to this and the single transition event, the study is assigned low quality. In addition to only comparing group differences, another major limitation is using midnight to midnight data. This does not allow using the time of onset of the nightly sleep period on the

Saturday immediately pre-transitions to determine duration, which could bias the Sunday sleep estimates.

Taken together, there is minimal evidence of an effect of transitions on sleep duration and quality, no clear evidence of an effect on circadian rhythm, although the medium quality studies may be considered indicative of some individuals being at higher risk of disturbed sleep than others.

***Cognitive***

Similar to Appendix Section 7, this subsection includes various outcomes that can be related to cognition and cognitive performance. Two studies concern driving simulator fatigue,[150] [151] one study assessed criminal activity,[152] one study assessed worker engagement,[144] one study assessed healthy consumer choices,[153] one study uses the psychomotor vigilance test,[133] one study assessed memory by means of the Mnemonic Similarities Task,[129] one study examined GoogleTrends search timings,[154] one study assessed whether perceptions of social norm violations changed,[147] one study assessed high school test performances,[155] and two studies consider the stock market, specifically investor sentiment and investor reaction.[156] [157]

Orsini et al. (2023, 2024) report increased driving simulator fatigue in the week post DST-Onset compared to pre DST-Onset in Italy in 337 young adults in the years 2022-2023.[150] [151] They use repeated measures, conducted placebo tests, and confirmed their findings in two different samples. Therefore, the studies together are rated high quality.

For Santiago de Chile in the years 2005-2010, Domínguez et al. (2023) find an increase and decrease in overall criminal activity (apparently driven by robbery) during sunset hours by 20% following DST-Offset and DST-Onset, respectively.[152] They find no significant differences when considering sunrise hours and whole day (albeit the estimates are in the opposite direction and same direction to sunset hours, respectively) and find no evidence of crime displacement although this cannot be completely ruled out. They analyse data for the city of Santiago specifically by regression discontinuity design. The data stem from an administrative database supported by the Chilean government and is collected by national police. The findings are supported with various robustness tests, including using two different sources of variation caused by delays to enactment of DST by drought and earthquake in 2008 and 2010. The study is rated high quality.

Völker et al. (2023) report lower work engagement on the first and second Monday following DST-Onset compared to the previous Monday in the year 2019 in a sample of 155 employees in Berlin.[144] Part of the effect appeared to be mediated by sleep quality and was more pronounced in later chronotypes. The analyses did not utilise repeated measures although some participants provided data for both Mondays. DST-Offset was not assessed. The study is rated medium quality. Work engagement was assessed by a survey but specifics are not described.

Janakiraman et al. (2024) find poorer consumer choices with DST-Onset but not DST-Offset.[153] Specifically, they compared differences in packaged snack consumption and fitness centre visits between the day pre and the day post transition in Arizona in the USA (which does not observe DST) with neighbouring states that do implement DST. Adverse effects of DST-Onset were identified for both outcomes. The findings persisted when comparing the difference in differences for five days pre and post transition and placebo tests were negative. The data came from a marketing research company and the authors consider it representative of the US population. However, in the first part of the study, the control group contains only 50 participants, which is difficult to reconcile as representative; thus, the study is rated medium quality.

Analysing data from GoogleTrends in Italy from 2015-2020, Domenie et al. (2023) found a phase shift in the peak of relative search volume (adjusted to universal standard time) for terms specific to sleep, sleep/health-related medication, and several unrelated random terms when comparing one week pre and one week post transitions.[154] The phase shifts were in line with what might be expected as individuals shift their behaviour and social times (relative to universal standard time) following transitions. The study can be rated as medium quality as only days wherein a diurnal rhythm could be fit to the data were used. Overall, though, there is little to be concluded other than individuals appear to shift their cognitive behaviour in terms of timing of Google searches across transitions in line with what would be expected.

Regarding high school test performance, Coury (2025) assessed the impact of DST-Onset transitions among 10th grade students in Ohio using a fixed-effects difference-in-differences design.[155] The neighbouring state of Indiana served as a comparison group; graphical trend analysis confirmed trends in test-outcomes to be parallel. The analysis included data from n=609 school districts in Ohio (2005–2014) and n=289 school districts in Indiana (2002–2008). All regressions were weighted by the number of students taking the test, and standard errors were clustered at the school district level to account for within-district correlation. In Ohio, a 2007 policy change shifted the DST-Onset transition to occur immediately before the Ohio Graduation Test week, with the reading test administered on Monday and the math test on Tuesday. In contrast, Indiana's standardized exams (ISTEP+) were administered in September, well outside the DST transition period, and thus assumed to be unaffected, as would 2002-2004 in Ohio. Data from the year 2009 was excluded because the test week deviated from the usual schedule. The author found that the DST-Onset transition led to statistically significant declines in test pass rates: -2.51% for reading and -1.9% for math. Sensitivity checks – including restricting the sample to border counties and to districts with high minority student populations – were consistent with the main results. The main limitation is that the control tests in Indiana did not involve the method of measurement and were at a different time of year that is quite far from the Ohio group; thus, the study is rated low quality.

The study designs and limitations for those considering memory, psychomotor vigilance, medical malpractice claim payment amounts are described already above and are all rated low quality. Specifically, Al-Bakry et al. (2024) report no clear effects on memory in healthy young adults in Canada following transitions.[129] Labarca et al. (2024) report that psychomotor vigilance parameter scores were deteriorated following DST-Offset and DST-Offset.[133] Gao et al. (2024) report higher than expected payment decisions for medical malpractice claims in the week post DST-Onset.[141] Regarding the latter study and in addition to the limitations described above, that no difference-in-differences for the weeks pre and post DST-Onset are observed, that whole year payments appear appreciably lower in the DST-utilising state than in control states, and that there is no indication for taking paired measurements into account (i.e., by year), the study quality is rated low.[141]

The study by Halfmann et al. (2025), which is also described under Sleep and Circadian Rhythms, also assessed perceptions of violations of social norms. There were no differences between the Monday post DST-Onset and the Monday two weeks later for ratings of real-life social norm violations (e.g., cycling on the footpath, applauding at a funeral) or for a monetary distribution game (third-person Dictator game). There were also no differences when assessing less severe violations or more severe violations.

Kim et al. (2023) support what was observed in studies of stock market investor sentiment in Appendix Section 7, insofar as there does not appear to be a transition effect. They used the same dataset as the original study that identified an effect and that sparked the accumulation of studies on the topic (described in Appendix Section 7) and included an extension of the dataset to 2023 (no differences observed with extension). The authors used an extreme bounds analysis (EBA), which indicated a consistent negative tendency of a potential transition anomaly, but results were not statistically significant at the 0.1% level, Bayes factor values were low, and even the best models had low explanatory power. The study quality is rated medium. Different from investor sentiment, Kleppe et al. (2024) report reduced investor reaction (conceivably cognitive impairment) in the USA – proxied by the Earnings Response Coefficients in the week following DST-Onset compared to other earnings announcements within the surrounding months (February, March, April).[157] The finding was more pronounced among firms with higher levels of retail trading activity and transient institutional ownership and robust in terms of various models and placebo tests. The findings are consistent regardless of whether earnings news is "good" or "bad", which is more indicative of cognitive impairment rather than a susceptibility toward pessimism following bad earning news. No significant effects following DST-Offset are reported. The study quality is rated high.

***All-Cause Mortality, Neurologic, & Gastrointestinal Section Synthesis***

There is only one study on mortality in this subsection and is rated high quality. Zhao et al. (2023) report no change following DST-Onset but decreased mortality following 0-7 weeks DST-Offset compared to what is expected from seasonal trends, with consistency in all-cause and leading cause-specific outcomes and some heterogeneity by time zone and demographic.[158] The study was conducted in 48 states across the contiguous US from 2014-2020. Higher risk individuals may be offered more protection with DST-Offset. The biggest difference was observed in the eastern time zone, which also has the highest population. Placebo tests in Hawaii and Arizona presented no difference. This finding lends support in favour of maintaining transitions.

There is only one study on neurological endpoints in this subsection; namely, the study by Göbel et al. (2025) on migraines in Germany.[159] They studied n=258 patients treated at a German tertiary headache centre from 2020-2022 and observed a significant increase (~6%) in migraine frequency on the Sunday or (presumably the authors mean and) Monday post DST-Onset compare to corresponding days in the week after but not the week prior. The opposite (~1-5% decrease for days on the week pre and first week post compared to the second week post) was observed across DST-Offset. That no differences between the Monday immediately post-transition compared to the Monday pre-transition (for both transitions) are observed but are compared to the Monday one week later is unexpected. Placebo tests would have been beneficial to rule secular trends. It is also unclear why pooled data for e.g., weeks or work days was not assessed. The study is categorised as medium quality but the unexpected nature of the finding, i.e., that it might take some time to develop, warrants caution.

***Conclusion***

Given the number and specificity of the subsections, the conclusions are presented here in corresponding bullet points.

- The cardiovascular subsection synthesis of this update strengthens the conclusions from the Cardiovascular Section of the Appendix (increased AMI following DST-Onset).

- The psychiatry subsection synthesis of this update strengthens the conclusions in the Psychiatry Section of the Appendix (little-to-no effect, especially for suicides).

- The traffic subsection synthesis of this update strengthens the conclusions from the Traffic Accidents Section of the Appendix, adding to the argument regarding USA and non-USA differences, to the conclusion regarding a role for change in daylight availability at specific times affecting accident risk directly, and it opens the door to other confounding factors regarding the associations between transitions and traffic accidents. There is also indication that DST may be preferred to Standard Time during the initial months that would follow the DST-Onset transition and initial months that would follow the DST-Offset transition.

- The finding from the Non-traffic Accidents, Injuries, & All-Cause Hospital Admissions Section of the Appendix is that, it appears unlikely that transitions increase risk of non-traffic injury, but the evidence is not conclusive. The finding that DST-Offset may be associated with decreased work accidents in the non-traffic accidents subsection synthesis of this update is in line with no clear effect following DST-Onset and lends more support for a possible benefit of DST-Offset.

- The sleep and circadian subsection synthesis of this update does not change the conclusions from the Sleep & Circadian Section of the Appendix of little-to-no effects, although there is more support for individual susceptibilities.

- The findings from the cognitive subsection synthesis of the update support the conclusions from the Cognitive Section of the Appendix insofar as adverse effects of DST-Onset on driving simulator performance are observed, that DST-Onset and DST-Offset may decrease and increase crime that causes physical harm but that ambient light for vision appears to be driving this, and that investor sentiment does not appear affected by transitions. That workplace engagement may be affected is supported by a slightly stronger study in this update compared to those on work-place attention in the Cognitive Section of the Appendix. A potential new finding is reduced investor reaction (conceivable proxy for cognitive impairment) following DST-Onset.

- The one high quality update study on all-cause mortality does not support an increase in mortality following DST-Onset but adds that mortality may be decreased following DST-Offset. Regarding neurological outcomes there was one medium quality study in this update that suggests a delayed effect of DST-Onset on increased migraines.

**Update-Table 1: Overview of studies in "Update" Section, grouped by subcategorizations used in synthesis, then alphabetically ordered.**

| Author (Year) | Population/Data | Exposure/Comparator | Statistics | Results | Rating |
|---|---|---|---|---|---|
| ***Cardiovascular*** | | | | | |
| Al-Bakry (2024)[129] | N=10 (DST-Offset) and 11 (DST-Onset) healthy adults, aged 18-35 years, Ontario-Canada | 7th day pre vs 1st, 3rd, and 5th days post transitions, 2022-2023 | RM-ANOVA, Student's t-test, and Bonferroni-Holm adjustment | No clear effects on measures of vascular stiffness, blood pressures, or augmentation index. | L |
| Kazirod-Wolski (2023)[130] | n=874,031 patients from Polish Register of Interventional Cardiology Procedures | First 3 days and 7 days post transitions, unclear reference periods, 2014-2021 | $\chi^2$/Fisher Exact Test, Student's t-test, Multiple Poisson regression | Ambiguous due to discrepancies between figures and tables and text. | L |
| Satterfield (2024)[131] | US Medical and Pharmacy Claims Database, OptumLabs Data Warehouse, ≥ 18 years with medical insurance coverage | 1 week post vs 2 weeks pre & weeks 2-3 post transitions, 2015-2019 | Bayesian hierarchical Poisson regressions. | Mild-to-moderate probabilities of low ↑ cardiovascular events following DST-Onset. No change following DST-Offset. | M |
| Tanaka (2024)[132] | Indiana Hospital Discharge Data, USA | 4 weeks post vs 4 weeks pre transitions, 2002-2012 | Regression discontinuity | ↑AMI following DST-Onset, returning to normal by week 3. No change following DST-Offset.<br>↑AMI in years with transitions compared to years without. | H |
| ***Psychiatry*** | | | | | |
| Labarca (2024)[133] | N=30 healthy male university students, Los Angeles-Chile | 2 weeks pre vs post transitions, 2022 | Paired Wilcoxon Signed-Rank Test | No effects on depressive symptoms following transitions. | L |
| Reis (2023)[134] | County-level populations and suicides from CDC Wonder database, for Mountain, Central, and Eastern time zones, USA | 5° longitude categories within time zones, 2010-2018 | Left-centred Poisson regressions using integrated nested Laplace approximation | ↑Suicides in western partition of time zones. | L |
| Plöderl (2024)[135] | Suicide reports from national statistics offices of Austria, Switzerland, and Sweden | 2 weeks pre vs post transitions, 1981-2022 | Poisson regression and Bayesian Changepoint analyses | No differences in suicides. | H |
| ***Traffic Accidents*** | | | | | |
| Gillmore (2025)[137] | Automobile accident reports from National Commission of Transit Security (CONASET), Chile | Pre vs post transitions with different bandwidths,<br>&<br>Transition to- + 2 months of living with- DST vs Standard Time and vice versa, 2002-2018 | Regression discontinuity and difference-in-differences | ↓Accidents in mornings following DST-Onset but ↑following DST-Offset, the opposite observed in afternoons, and no differences in evenings.<br>↓Accidents living with DST vs Standard Time in late spring (~3.5%) and ↑ living with Standard Time vs DST in late autumn (~1.5%). | H |
| Laliotis (2023)[136] | Road Accident Reports by Hellenic Statistical Authority (ELSTAT), Greece | ~2 weeks pre vs post transitions, 2006-2016 | Regression discontinuity | ↓Serious vehicle accidents, ↑fatal vehicle accidents following DST-Onset. ↑Total and minor vehicle accidents following DST-Offset | H |
| Woods (2024)[138] | Fatality Reporting System, USA | 5 weeks pre- and post transitions, 2010-2019 | | ↑fatal crashes following DST-Onset (+3.3%), ↓after DST-Offset (−2.4%).<br>↑motor vehicle occupant deaths (+12%) & ↓ pedestrian/bicyclist deaths (−24%) after DST-Onset; reversed pattern after DST-Offset | M |

| | | | | | |
|---|---|---|---|---|---|
| ***Non-traffic Accidents, Injuries, & All-Cause Hospital Admissions*** | | | | | |
| Depalo (2023)[139] | Recorded work accidents among 16-70 years old, National Institute of Insurance against Accidents at Work, Italy | Averaging effects over increasing bandwidth (up to 5 days pre and post transitions) 2013-2017 | Regression discontinuity, averaging effects over increasing bandwidths | No clear effect on injuries, disabilities or deaths following DST-Onset. ↓Injuries and disabilities but not deaths following DST-Offset. Effects more apparent in older vs younger workers and in the north vs south of Italy. | H |
| Gao (2024)[141] | N=288,432 malpractice claims from National Practitioner Data Bank, United States | 1 week pre vs post DST-Onset vs rest of year, 1990-2018 | ANOVA, logistic regression, with state*time period interaction term. | No effect on incident severity. | L |
| Kocali (2023)[140] | Work accident claims submitted to Republic of Turkey Social Security Institution. | Unclear | Unclear | Unclear | L |
| ***Sleep & Circadian*** | | | | | |
| Al-Bakry (2024)[129] | As above | As above | As above | No clear effects on sleepiness, fatigue, sleep quality, quantity, latency, or efficiency. | L |
| Angelino (2024)[149] | N=62 adult participants (58% female), median 26 years, with type 1 diabetes and in stable treatment. Naples, Italy | 2 weeks pre vs post DST-Onset (2023) and DST-Offset (2022) | Wilcoxon signed-rank test | No clear effects on sleep quality or duration. | L |
| Costa-Font (2023)[143] | Adult 15+ years, German Socio-Economic Panel | 4-weeks pre and post DST-Onset, 2008-2018 | Regression discontinuity | ↓ sleep-satisfaction following DST-Onset | M |
| de Lange (2024)[148] | n=11,780 UK Biobank participants aged 43-78 years. | 2 weeks pre vs post transitions, 2013-2015 | Independent one and two-tailed t-tests | ↓sleep duration on DST-Onset Sundays, ↑sleep duration on DST-Offset Sundays | L |
| Ferguson (2024)[145] | Fitbit activity tracker data for n=291/333 (DST-Onset/Offset) parents aged 18-65 | 7 days pre vs post transitions, 2019-2021 | Multilevel Mixed-Effects Linear Regression with Random Intercepts | ↓Sleep duration and ↑sedentary behaviour following DST-Offset. | L |
| Halfmann (2025)[147] | N=724 and 986 participants in the first study and in the repeat, respectively. The repeat study included only employed persons (mean 41.19±12 years; 46.6% female). The first study included unemployed, retired, and students too (mean 44.75±13.4 years; 58.4% female). UK | Monday post DST-Onset vs Monday two weeks later | One and two-tailed t-tests | No clear differences in sleep duration, sleep quality, or sleepiness. | L |
| Labarca (2024)[133] | As above | As above | As above | No clear effects on questionnaire-assessed sleep quality, insomnia, or sleepiness. ↓Sleep duration, time-in-bed, efficiency & ↑awakenings following DST-Offset only using wrist actigraphy. | L |
| McHill (2024)[142] | N=37 undergraduate students aged 18-22. | 1 week pre vs post DST-Onset | ANOVA, paired t-test, multiple linear regression | Lower-fat diet associated with faster sleep midpoint shift following DST-Onset. | M |
| Völker (2023)[144] | N=155, mean 39 (SD 13) years, full time non-shiftwork employees in Berlin, Germany | 1st Monday pre vs post DST-Onset. 2019 | Non-linear mixed effects regression | ↓Sleep duration and quality following DST-Onset. | M |
| Zofaghari (2023)[146] | N=2594, 45-85 years, Canadian Longitudinal Study of Aging | 1 week pre vs post transitions, 2011-2015 | Linear and logistic regression | ↓ Questionnaire-based sleep duration following DST-Onset. ↑Sleep dissatisfaction, sleep-onset | L |

| | | | | | |
|---|---|---|---|---|---|
| | | | | and maintenance insomnia, and hypersomnolence with adequate sleep following DST-Offset. | |
| ***Cognitive*** | | | | | |
| Al-Bakry (2024)[129] | As above | As above | As above | No clear effects on memory performance. | L |
| Coury (2025)[155] | Pass rates for 10th graders first attempt of the Ohio Graduation Test (2005-2014) and Indiana's standardized exams (ISTEP+) (2002-2008) from Dept of Education website, USA | DST-Onset and living with DST- vs living with Standard Time, 2002-2014 (not incl. 2009) | Difference-in-differences | ↓Pass rates following DST-Onset transition after introduction of DST practice (2.51% for reading, 1.9% for math) | L |
| Domenie (2023)[154] | GoogleTrends relative search volume (hourly data), Italy | 15 days pre vs post transitions, 2015-2020 | Unpaired student t-test, RM-ANOVA | Phase advance/delay in peak of diurnal rhythm of relative search volumes following DST-Onset and Offset, respectively. | M |
| Domínguez (2023)[152] | Administrative database on crime reports by Chilean government, Santiago-Chile | 3 weeks pre vs post transitions, 2005-2010 | Regression discontinuity | ↑/↓ robbery incidents (~30%) and overall crime activity (~20%) during sunset hours following DST-Offset and Onset, respectively. | H |
| Gao (2024)[141] | As above | As above | As above | ↑malpractice payments above expected | L |
| Halfmann (2025)[147] | As above | As above | As above | No clear differences in perceptions of violations of social norms | L |
| Janakiraman (2024)[153] | Marketing research agency, "representative of US population" | 1 day pre vs 1 day post transitions, no years provided. | Differences-in-differences across transition dates using states with/without DST | ↑poorer consumer behaviour following DST-Onset.<br>No change following DST-Offset. | M |
| Kim (2023)[156] | Mean daily returns, US | transitions weekends vs other weekends, 1965-2023 | Extreme bounds analysis | No clear evidence of a transition effect on market returns | M |
| Kleppe (2024)[157] | N=37,544 quarterly earnings announcements, USA | 1 week post DST-Onset vs surrounding month announcements | Probit regression, Mills ratio, OLS regression | ↓earnings response coefficients in the week following DST-Onset | H |
| Labarca (2024)[133] | As above | As above | As above | ↓PVT scores following both transitions. | L |
| Orsini (2023)[151] | N=18 males, 21-30 years, with driving experience, Italy | 1 week pre vs post DST-Onset, 2022 | Linear mixed effect models for repeated measures design | ↑Driving simulator fatigue following DST-Onset in Italy | H |
| Orsini (2024)*[150] | N=37 adults, 19-30 years, with driving experience, Italy | 1 week pre vs post transitions, 2022-2023 | Linear mixed effect models for repeated measures design | ↑Driving simulator fatigue following DST-Onset in Italy | H |
| ***All-Cause Mortality, Neurologic, & Gastrointestinal*** | | | | | |
| Göbel (2025)[159] | n=258 patients (mean age 51.54 ± 11.78; 86.8% female) treated in tertiary headache centre, Germany | Sunday/Monday pre vs Sunday/Monday post DST Onset/Offset, 2020-2022 | Two-sided Fisher's exact test | ↑migraine frequency at 2 weeks post DST-Onset and ↓ t 2 weeks post DST-Offset. | M |
| Zhao (2023)[158] | Nationwide study of state level mortality data, National Center for Health Statistics, USA | 0-8 weeks post transitions vs rest of year, 2014-2020 | Standardised rate ratios from negative binomial log linear regressions. | No effects following DST-Onset. ↓Mortality following DST-Offset. | H |

L=low, M=medium, H=high. *The study included that pilot sample used in the corresponding earlier study by the same author.

#### Update-Table 2: JBI Quality Indicators of Studies

(Questions and possible answers are shown in detail in Table footnote)

| Author (Year) | Q1 | Q2 | Q3 | Q4 | Q5 | Q6 | Rating | Notes |
|---|---|---|---|---|---|---|---|---|
| Al-Bakry (2024)[129] | Y | Y | N* | Y | Y* | N* | L* | *Comparison days are not corresponding pre and post transitions. The sample size is very low. |
| Angelino (2024)[149] | Y | Y | N* | Y | Y | N | L | *Unclear whether repeated measures were on corresponding days. |
| Costa-Font (2023)[143] | Y | U* | Y | Y | Y | N | M | *No repeated measures. |
| Coury (2025)[155] | Y | Y | N* | N* | Y | N | M | *Some different methods for measures of outcomes at different times were used for difference-in-differences estimates. Even with adjustment for the covariates that account for differences, there may still be measurement bias. |
| de Lange (2024)[148] | Y | N* | N* | Y | Y | N | L | *No repeated measures and sleep duration determined from mid-night to mid-night rather than e.g., mid-day to mid-day, which could bias results. |
| Depalo (2023)[139] | Y | Y | Y | Y | Y | N | H | |
| Domenie (2023)[154] | Y | U* | U* | N/A* | Y | N | H | *Analysis of GoogleTrends data rather than individuals. |
| Domínguez (2023)[152] | Y | Y | Y | Y | Y | N | H | |
| Ferguson (2024)[145] | Y | U* | U* | N/A | U* | N | L* | *The contribution of lack of wear time, whether participants contributed data pre and post transitions, and well the algorithm can distinguish potentially small differences correctly is questionable. |
| Gao (2024)[141] | N* | U* | U* | N | N* | N | L | *Control states 'control' values not similar to DST states suggesting comparisons may not be appropriate due to residual confounding and regression to the mean. Paired measurements by year were not considered. |
| Gillmore (2025)[137] | Y | Y | Y | Y | Y | N | H | |
| Göbel (2025)[159] | N* | N/A* | N/A* | Y | Y | N | M | *There was no matching in the study design. |
| Halfmann (2025)[147] | Y | U* | U* | Y | Y | N | L | *No participant details or comparisons between groups; no repeated measures. |
| Janakiraman (2024)[153] | Y | U | Y | Y | Y | N | M* | *In part 1 of the study, it is hard to envisage 50 participants being representative of the US population. |
| Kazirod-Wolski (2023)[130] | Y | Y | U | U | U | N | L* | *Several issues including discrepancies between graphs and text and ambiguity surrounding the reference periods. |
| Kim (2023)[156] | Y | N/A | N/A | N/A | Y* | N | M* | *Stock market returns as a proxy of investor sentiment. Better analytical methods have been reported. |
| Kleppe (2024)[157] | Y | N/A | N/A | N/A | Y | N | H* | *Investor reaction (conceivably cognitive impairment) proxied by earnings responses. |
| Kocali (2023)[140] | N* | Y | Y | Y | U* | Y* | L | *What was assessed, how, and presentation of results is too ambiguous and contains irregularities. |
| Labarca (2024)[133] | Y | Y | Y | U* | Y | N | L* | *It is unclear exactly when or how often questionnaire data was collected during the two weeks pre and post transitions study periods. It is unclear when PVT data was collected. Only one transition year was assessed. |
| Laliotis (2023)[136] | Y | N/A | U | N/A | Y | N | H | |
| McHill (2024)[142] | Y | Y | Y | N/A | Y | N | M* | *The sample size is low but repeated measures are used. |
| Orsini (2023)[151] | Y | Y | Y | N/A | Y | N | H* | *See next row. |
| Orsini (2024)[150] | Y | Y | Y | N/A | Y | N | H* | *Orsini et al. 2023 and 2024 are complementary in terms of repeated investigation and placebo tests. |
| Reis (2023)[134] | U* | U* | U* | N/A | Y | N | L | *The study conclusions can be affected by ecological fallacy. |
| Plöderl (2024)[135] | Y | Y | Y | Y | Y | H | H* | *Placebo and sensitivity analyses, size of study, and analytical methods warrant high quality rating. |
| Satterfield (2024)[131] | Y | Y | U | N/A | Y | N | M* | *Participants were limited to only those with sufficient medical insurance, which may bias outcomes. |
| Tanaka (2024)[132] | Y | Y | N/A | Y | Y | N | H | |
| Völker (2023)[144] | Y | U* | U* | Y | Y* | N | M* | *Some participants only provided data pre or post transition. The study would have benefited from analysis using repeated measures. There is no indication that participants were unaware the study was about DST, which could bias findings. Only one transition period was assessed. |
| Woods (2024)[138] | Y | Y | Y | Y | Y | N | M* | *No placebo tests. |
| Zhao (2023)[158] | Y | Y | Y | Y | Y | N | H* | *Placebo tests presented negative findings. |

| | | | | | | | | |
|---|---|---|---|---|---|---|---|---|
| Zolfaghari (2023)[146] | N* | N* | U* | Y | Y | N | L | *The questionnaire regarding sleep enquired about the previous month, there were no repeated measured, there were differences in group characteristics that may affect sleep. |

**JBI Critical Appraisal Checklist for Quasi-Experimental Studies (Adapted)**
Answers can be either Y=Yes, N=No, U=Unsure, or N/A.

1. Is it evident which factor is considered the 'cause' and which one is the 'effect'? Is there no ambiguity regarding the sequence of variables under investigation?
2. Were the characteristics of the study participants involved in comparisons adequately matched?
3. Were the exposures/treatments (beyond that of interest in our study) of the study participants involved in comparisons adequately matched?
4. Were the methods used to measure the outcomes the same for participants who were part of different comparisons?
5. Were appropriate statistical methods employed to analyse the data?
6. Are there any irregularities in the data reported?

#### Conclusion Robustness to Quality Ratings

Philip Lewis & Jonas P. Wallraff

Many of the included studies report multiple outcomes; thus, some studies were assigned to more than one of Appendix Sections 1-7. Ten of these studies were assigned different quality ratings in the different sections (Conclusion Robustness to Quality Ratings-Table 1). In the subsections below (which are aligned with Appendix Sections 1-7), we consider whether changes to the quality ratings of studies (based on differences in how they were rated in the different sections) might affect conclusions.

**Conclusion Robustness to Quality Ratings-Table 1: Differences in Quality Ratings Across Sections**

| | Cardiovascular | Psychiatry | Traffic Accidents | Non-traffic Accidents, Injuries, & All-Cause Hospital Admissions | Sleep & Circadian | All-Cause Mortality, Neurologic & Gastrointestinal | Cognitive |
|---|---|---|---|---|---|---|---|
| Barnes (2009)[56] | | | | M | L | | |
| Borisenkov (2017)[32] | | M | | | | | L |
| Fetter (2014)[24] | | M | | | H | | |
| Goodwin (2023)[12] | M | | M | | | | H |
| Jankowski (2014)[33] | | L | | | H | | |
| Jin (2020)[11] | M | | | H | L | | |
| Lahti (2008b)[25] | | M | | | L | | |
| Lindenberger (2019)[15] | M | M | L | M | | | L |
| Nohl (2021)[53] | | | H | M | | | |
| Zhang (2020)[6] | M | L | | M-H | | M | |

L=Low quality, M=Medium Quality, H=High Quality

##### *Cardiovascular*

Four studies in this section contributed outcomes to other sections and were assigned different quality ratings. Changing the medium ratings for Goodwin et al. (2023), Jin et al. (2020), Lindenberger et al. (2019) and Zhang et al. (2020) to high or low as per the Table above only serves to strengthen the conclusions from this section.[6 11 12 15] Moreover, the same conclusions are reached for MI here as in a recent meta-analysis despite some differences in study quality ratings.[1]

##### *Psychiatry*

Of the twelve studies included in this section of the Appendix, six that contributed outcomes to other sections were assigned different quality ratings. The studies by Fetter et al. (2014) and Lahti et al. (2008b) in the 'Null Associations' subsection of the Psychiatry Section were medium quality.[24 79] Changing the quality ratings to high and low, respectively, does not affect conclusions for this section. Four studies in the 'Weak or Inconsistent Associations' subsection were rated as low[6 33] and medium quality.[15 32] Similarly, changing these quality ratings to those assigned in other sections of the Appendix does not impact conclusions.

##### *Traffic accidents*

Of the 23 studies included in this section of the Appendix, three that contributed outcomes to other sections were assigned different quality ratings. The studies by Lindenberger et al. (2019) and Nohl et al. (2021) in the 'Traffic Accident-Linked Hospital Admissions' subsection that were rated high and low quality, respectively, were rated medium quality elsewhere.[15 53] Changing both to medium strengthens the report of no change in hospital admissions/autopsies post transitions and weakens any suggestion of increased admissions. Changing the quality rating for Goodwin et al. (2023) from medium to high would strengthen the finding of increased traffic accident fatalities following both DST-Onset and DST-Offset.[12] The original overall conclusion, i.e., that both transitions may increase fatal accidents, at least on some of the initial days post transition and in some places, remains.

##### *Non-traffic Accidents, Injuries, & All-Cause Hospital Admissions*

Of 15 studies included in this section of Appendix, five that contributed outcomes to other sections were assigned different quality ratings. If the study by Barnes et al. (2009) in the 'Workplace Accidents' subsection with medium quality was changed to low, this would not change the original conclusion that there is little-to-no evidence of DST-Onset affecting workplace accidents.[56] In the 'Non-occupational Accidents/Traumas' subsection, changing the study by Nohl et al. (2021) from medium to high quality and the study by Zhang et al. (2020) from high-medium to low quality would strengthen the conclusion of a null effect of transitions on these outcomes.[6 53] This is because Zhang et al. (2020) is the only study to suggest a transition effect and greater weight to the study by Nohl et al. (2021) would result in a total of two high quality studies and one medium quality study supporting a null effect. Changing the quality rating from high to low for Jin et al. (2020) strengthens that no clear conclusion can be drawn from the current literature regarding all-cause hospital admissions changes following transitions.[11] Changing from medium to a low quality rating for Lindenberger et al. (2019) in the 'Accidental Deaths' subsection would slightly attenuate the strength of the conclusion that increased accidental deaths are observed following DST-Onset, but would not change it.[15]

##### *Sleep & Circadian*

Four studies included in the 'Acute Transition Effects' subsections of this section of the Appendix contributed outcomes to other sections and were assigned different quality ratings. Overall, changing Lahti et al. (2008b) and Barnes et al. (2009) from low to medium,[56 79] changing Fetter et al. (2014) from high to medium,[24] and changing Jin et al. (2020) from low to high would strengthen support for DST-Onset and DST-Offset decreasing and increasing sleep duration, respectively.[11] However, the strengthening would not be by much.

Two studies in the 'DST vs Standard Time (and longitude) contributed outcomes to other sections and were assigned different quality ratings. Jankowski et al. (2014) was rated high quality here but low quality elsewhere and Borisenkov et al. (2017) was rated low quality here but medium elsewhere.[32 33] Changing the quality ratings would not change the conclusions that eastern populations within the same time zone appear more likely to be more morning chronotypes but would slightly enhance the evidence base that perennial DST may be more detrimental than perennial Standard Time regarding sleep duration and social jet lag.

##### *Cognitive*

Two of the four studies in the 'Criminal Activity' subsection of this Appendix section contributed outcomes to other sections and were assigned different quality ratings. Lindenberger et al. (2019) was rated low quality and Goodwin et al. (2023) as high quality; both were rated medium quality elsewhere.[12 15] Applying the medium rating to both articles would not change the conclusions of the subsection that there is no sufficient evidence to abandon DST practices based on these cognitive outcomes.

##### *All-Cause Mortality, Neurologic & Gastrointestinal*

Zhang et al. (2020) was rated as medium quality in 'Gastrointestinal' subsection.[6] Changing this to either low or high quality (as is assigned elsewhere) would not change the conclusion of the subsection as the results of this study are inconclusive and the only other study by Föh et al. (2019) is rated low quality.[127]

##### *Conclusion*

Overall, changing the quality ratings of studies to be in line with other Appendix Sections has negligible effects on conclusions from the respective sections.
